# Altered kynurenine and indole tryptophan pathways in calcific aortic stenosis: cross-sectional evidence for a gut-host metabolic imbalance (GUT-CAS)

**DOI:** 10.64898/2026.05.22.26353844

**Authors:** Caroline Chong-Nguyen, Sarah Atighetchi, Cyril Ferro, Bahtiyar Yilmaz, Harry Sokol, Matthias Siepe, David Reineke, Selim Mosbahi, Daijiro Tomii, Masaaki Nakase, Christoph Wingert, Liam Tanner, Camille Dupuy, Lydie Nadal-Desbarats, Yara Banz, Tereza Losmanovà, Pamela Nicholson, Aparna Pandey, Yvonne Döring, Thomas Pilgrim

## Abstract

**Background:** Calcific aortic stenosis (CAS) is a progressive valvular disease characterized by lipid accumulation and osteogenic remodeling. The contribution of gut microbiota–derived metabolites to CAS remains unclear.

**Objective:** To investigate gut microbiome–associated metabolomic signatures in calcific aortic stenosis and to assess genetic evidence for causal contributions of selected metabolic and inflammatory markers using Mendelian randomization.

**Methods:** In a prospective cohort of 54 CAS patients and 41 BMI-matched controls, we performed integrated 16S rRNA gene sequencing and targeted metabolomic profiling. MR analyses were conducted to assess genetically predicted effects of selected metabolic and inflammatory markers on CAS.

**Results:** The principal finding was a selective activation of tryptophan metabolism toward the kynurenine pathway. The kynurenine-to-indole-3-sulfate (KI) ratio was the only tryptophan metabolite finding to survive BH-FDR correction (p = 0.0031, q = 0.013). Its inverse association with indexed aortic valve area (Spearman ρ = −0.310, p = 0.036, n = 46) was attenuated after adjustment for age and BMI (p = 0.111), suggesting a relationship that may be partly confounded by these variables. In contrast, global microbiome diversity and overall community structure were preserved. MR analyses identified lipoprotein(a) as a genetically supported causal risk factor for CAS (Wald ratio OR = 1.116, p < 0.001), independent of the gut-related metabolic findings. Exploratory sex-stratified analyses suggested stronger kynurenine-pathway alterations in male patients, but require replication.

**Conclusion:** In this exploratory study, calcific aortic stenosis was associated with a selective shift in tryptophan metabolism toward the kynurenine pathway, observable in the absence of global gut microbial differences. These findings are exploratory and hypothesis-generating, suggesting that altered tryptophan partitioning at the gut-host interface may be a feature of CAS biology warranting prospective validation.

**HIGHLIGHTS:**

- Calcific aortic stenosis is associated with a selective shift in tryptophan metabolism toward the kynurenine pathway, without global gut microbial differences.
- This metabolic imbalance reflects a gut–host immunometabolic signal linked to structural disease severity.
- The kynurenine–indole axis may represent a biologically relevant pathway in valvular heart disease.
- Lipoprotein(a) emerges as an independent genetically supported causal risk factor for calcific aortic stenosis.

**CLINICAL PERSPECTIVES:** *Competency in Medical Knowledge:* Calcific aortic stenosis is associated with a selective shift in host tryptophan metabolism toward the kynurenine pathway, representing an immunometabolic signature linked to structural disease severity. This signal occurs in the absence of global gut microbial differences, indicating that pathway-specific metabolic alterations, rather than generalized microbial disruption, characterize the disease-associated metabolic profile.

*Translational Outlook:* The kynurenine–indole axis represents a promising immunometabolic signal associated with calcific aortic stenosis that warrants further investigation in longitudinal and independent cohorts. Future studies should determine whether this pathway tracks disease progression and whether it may provide additive value beyond established clinical and imaging markers. In parallel, mechanistic studies are needed to clarify whether modulation of tryptophan metabolism could inform novel therapeutic strategies in valvular heart disease.

## INTRODUCTION

Calcific aortic stenosis (CAS) is the leading cause of death from valvular heart disease in high-income countries, with prevalence increasing sharply with age(1,2). It is a progressive disorder driven by endothelial injury, inflammation, lipid infiltration, and osteogenic transformation of valvular interstitial cells (VICs). These processes converge to drive extracellular matrix remodeling and progressive valve stiffening, ultimately leading to hemodynamic obstruction and heart failure.

In parallel, growing evidence implicates the gut microbiota as a central regulator of host metabolic and inflammatory homeostasis. Microbial communities shape systemic physiology through the production of bioactive metabolites that influence immune signaling, redox balance, and lipid handling. These metabolites include trimethylamine N-oxide (TMAO) (3), bile acids (BAs) (4), short-chain fatty acids (SCFA) (5), and tryptophan derivatives(6). TMAO promotes inflammatory and osteogenic signaling in human aortic VICs, while tryptophan-derived metabolites, such as indoxyl sulfate and serotonin induce endothelial-to-mesenchymal transition and trigger osteogenic differentiation.(7,8) Preclinical studies suggest that these microbial metabolites directly affect valve cell biology, promoting calcific and fibrotic remodeling, thus supporting the existence of a gut-valve axis in the pathogenesis of CAS(9).

Despite these advances, the specific contribution of the gut microbiota-metabolite axis to CAS remains poorly defined. In particular, it is unclear whether CAS is associated with global microbiota disruption or with selective alterations in defined metabolic pathways, and how these changes relate to host factors such as adiposity and sex. Prior Mendelian randomization studies have investigated the causal contribution of conventional lipid fractions, lipoprotein(a), and inflammatory markers to CAS risk, and a proteomics-based MR study has identified circulating proteins associated with CAS susceptibility.(10–12) However, no study has integrated gut microbial composition, targeted metabolomic profiling, and Mendelian randomization within a single, phenotypically characterized cohort, a multi-modal approach that could link microbial ecology, downstream metabolite production, and potential causal pathways in a unified biological framework.

In this study, we performed an integrated analysis of gut microbiota composition and circulating microbial metabolites in patients with and without CAS. By combining 16S rRNA microbiome profiling with targeted multi-panel metabolomics and Mendelian randomization, we aimed to identify disease-associated gut microbial and metabolic signatures, to assess the directionality of key metabolite– disease associations, and to generate testable hypotheses regarding gut–host crosstalk in valvular calcification.

## MATERIALS AND METHODS

### Methods

#### Study Design and Participants

The GUT-CAS study was a prospective observational cohort study conducted at Bern University Hospital in Switzerland. Adult patients (≥18 years) with CAS and control participants without CAS were recruited. CAS severity was defined according to current echocardiographic guidelines(13). Controls had no evidence of aortic stenosis on echocardiography or cardiac CT. Study participants were matched for age, sex, and body mass index. Key exclusion criteria included presence of rheumatic heart disease or infective endocarditis, intake of antibiotics, corticosteroids, bile acid sequestrants, selective serotonin reuptake inhibitors, or antiretroviral therapy, a history of cholecystectomy or chronic liver disease, and inflammatory bowel disease.

All participants underwent standardized clinical evaluation, laboratory testing (including cardiac biomarkers, lipid profile, glycemic markers, renal and hepatic function), electrocardiography, and echocardiography. When available, CT calcium scoring (N=21 in CAS group, 3 in non CAS group) was used to quantify valvular calcification. The study was approved by the local ethics committee (KEK Bern, ID 2023-01173) and registered at ClinicalTrials.gov (ID NCT06021535). All participants provided written informed consent.

#### Biospecimen collection Stool Samples

Stool samples were collected at baseline in CAS patients and controls. Stool sampling was repeated in CAS patients annually. Participants followed standardized dietary instructions for 48–72 hours prior to sampling, avoiding probiotics, prebiotics, fermented foods, and excessive fiber intake. Samples were collected using OMNIgene•GUT | OM-200 (for microbiome) and OMNImet•Gut (for metabolomics) kits from DNAGenotek, according to the manufacturer’s instructions, transported at ambient temperature, and stored at –80 °C until analysis.

#### Blood Samples

Fasting blood samples were collected at baseline and follow-up visits. Serum was isolated by centrifugation and stored at −80 °C until analysis. Metabolomic analysis included BAs, tryptophan metabolites, TMAO, and SCFAs. As serotonin was measured in serum, which contains predominantly platelet-derived serotonin released during coagulation rather than exclusively circulating serotonin — platelet count was included as a covariate in sensitivity analyses for serotonin group comparisons.

### Microbiota Analysis Sequencing

Total DNA was extracted from stool samples using a magnetic bead–based method (ZymoBIOMICS DNA MagBead Kit, Zymo Research) on an automated platform (KingFisher Apex, Thermo Fisher Scientific), following manufacturer instructions. Samples were mechanically lysed using bead-beating to ensure efficient disruption of microbial cells. DNA quantity and quality were assessed using fluorometric and electrophoretic methods prior to library preparation.

Full-length 16S rRNA gene libraries (V1–V9 regions) were generated using the PacBio Kinnex platform and sequenced on the PacBio Revio system. This pipeline performs end-to-end analysis of circular consensus (HiFi) reads, including quality control, denoising, and taxonomic assignment. Briefly, high-fidelity reads were filtered based on quality and length thresholds, followed by amplicon sequence variant (ASV) inference using the DADA2 algorithm within the QIIME2 framework.(14,15)

Taxonomic classification was performed against Silva reference databases using QIIME2-compatible classifiers. The pipeline generates feature tables, taxonomic profiles, and summary visualizations at near species-level due to the use of full-length 16S sequences.(15)

Microbial community analyses were performed using the *phyloseq, vegan*, and *microbiome* packages. Alpha diversity metrics (Shannon, Simpson) and beta diversity (Bray-Curtis dissimilarity) were calculated, with group differences assessed using non-parametric tests and permutational multivariate analysis of variance (PERMANOVA), respectively. Ordination was visualized using principal coordinate analysis (PCoA).(16,17)

Differential abundance analysis was performed using MaAsLin3, applying multivariable linear models to identify taxa associated with clinical variables, including CAS status, BMI, and sex, while adjusting for relevant covariates. Both relative abundance and prevalence-based models were evaluated. False discovery rate (FDR) correction was applied using the Benjamini-Hochberg method, with q-values <0.05 (18)

The gut microbiome analysis was performed prior to the BMI-matching procedure on all participants with available 16S data (n = 50 CAS, n = 49 controls); BMI, age, and sex were included as covariates in the MaAsLin3 linear model to account for their independent effects on microbial composition. Because no significant differences in overall microbial community composition were observed between the matched and pre-matching cohorts, the microbiome analysis was not repeated in the BMI-matched subset.

To assess the impact of metabolic status, additional stratified analyses were performed by BMI category (normal weight vs. overweight/obese), including both binary classification models and continuous BMI analyses. Associations between microbial features and circulating metabolites were evaluated using correlation analyses and integrated statistical modeling.

### Metabolomic Analysis

Serum and fecal metabolites were quantified using high-performance liquid chromatography coupled to mass spectrometry (HPLC–MS). The following metabolite classes were analyzed:

• **Short-chain fatty acids:** acetate, propionate, butyrate, valerate, isobutyrate and isovalerate.

• **Bile acids:** primary and secondary bile acids (free and conjugated).

• **Tryptophan pathway metabolites:** tryptophan, kynurenine derivatives, serotonin derivatives, indole derivatives.

• **Choline pathway metabolites:** L carnitine, choline, TMA, TMAO.

All metabolite panels (bile acids, SCFAs, choline pathway metabolites, and tryptophan metabolites) were quantified using targeted LC-MS/MS with stable isotope-labeled or structurally analogous internal standards. Concentrations are reported as absolute values (nM or µM) derived from validated external calibration curves across the established LLOQ–ULOQ range. Batch accuracy and precision were monitored with quality control samples at three concentration levels. Detailed extraction protocols and MRM transitions for each analyte are provided in the Supplementary Methods. Serotonin was quantified as part of the tryptophan panel using this method. Because serotonin in serum reflects predominantly platelet-derived release during coagulation rather than circulating free serotonin, platelet count was included as a covariate in sensitivity analyses for serotonin group comparisons. The results of this sensitivity analysis are reported in the main text Limitations section. MRM transitions (precursor ion m/z, product ion m/z, collision energy, and chromatographic retention time) for all quantified analytes across all four metabolite panels (bile acids, SCFAs, choline pathway metabolites, tryptophan pathway metabolites) are listed in Supplementary Table 1 — MRM Transitions.

#### Follow-up and Clinical Data

Patients with mild, moderate, or asymptomatic severe CAS who did not undergo aortic valve replacement were followed annually for 5 years, with clinical assessment, echocardiography, and repeated stool/blood sampling. Patients who underwent valve replacement were followed longitudinally via telephone to monitor adverse outcomes. Controls were not followed over time, as gut microbiota composition is generally stable in the absence of disease.

Control subjects were enrolled in a cross-sectional design and were not followed longitudinally. The stability of the gut microbiome over the study period in controls cannot be assumed, as dietary changes, antibiotic exposures, and intercurrent illnesses can alter microbial composition; this is acknowledged as a limitation. Longitudinal stool data from CAS patients are being collected as part of a planned extension and will be reported separately.

### Statistical Analysis

In this exploratory study, we estimated that enrolling 50 participants per group (CAS and controls) would provide sufficient power to detect differences in microbiota and metabolite. Based on previously published studies with comparable designs and methodologies, we estimated that enrolling 50 participants per group (CAS and controls) would provide adequate power to detect significant differences in microbiota and metabolite profiles for moderate or strong effects(19–21). We therefore aimed to recruit slightly more than 50 CAS patients to account for potential exclusions or incomplete data, ultimately including all 54 eligible CAS patients. Controls were subsequently selected using BMI matching, resulting in a smaller matched non-CAS group (N = 41). This pragmatic approach prioritized inclusion of all eligible CAS patients and preservation of BMI comparability between groups, consistent with the exploratory nature of the study.

### Statistical tests

All analyses were performed using R (v4.5.1 for maaslin3 analysis and v4.2.1 for other analysis) and GraphPad Prism (v10). Continuous variables are expressed as mean ± standard deviation (SD) or median and interquartile range (IQR); categorical variables are reported as counts and percentages. Normality was assessed using the Shapiro–Wilk test. Between-group comparisons (CAS vs. controls) was performed using Student’s t test or Mann–Whitney U test. A two-sided P value <0.05 was considered statistically significant after correction for multiple testing with the Benjamini–Hochberg false discovery rate method.

For circulating and fecal metabolites, including bile acids, SCFAs, tryptophan derivatives, and choline/TMAO pathway compounds, concentrations were log-transformed when necessary to approximate normality. Group comparisons were performed using t tests or Mann–Whitney U tests.

Correlation analyses were used to link metabolite concentrations with specific bacterial taxa or clinical parameters. Tryptophan pathway ratios (kynurenine-to-indole, kynurenine-to-tryptophan, kynurenine-to-serotonin) were calculated from absolute concentrations measured within the same LC-MS/MS analytical run, using metabolite-specific stable isotope-labeled internal standards.

The continuous relationship between the kynurenine-to-indole ratio and indexed aortic valve area was assessed in CAS patients using Spearman rank correlation. Ninety-five percent confidence intervals were derived by 1000 bootstrap resamples. To assess robustness of the KI ratio–indexed AVA correlation to potential confounders, single-covariate sensitivity analyses were performed by computing partial Spearman correlations adjusting separately for: age, sex, eGFR, atrial fibrillation, diabetes, BMI, statin use, and anticoagulant therapy; partial correlations were estimated via ordinary least-squares residualisation of ranks. The robustness of the primary KI ratio finding to individual patient influence was assessed by leave-one-out (LOO) analysis, repeating the Mann–Whitney U test after removing each patient in turn.

### Matching procedure

Controls were matched to CAS patients according to age and sex at the cohort design stage. Matching according to BMI was applied algorithmically to limit confounding related to adiposity, which is a major determinant of gut microbiota composition and circulating BAs, SCFAs, and tryptophan-derived metabolites.

A caliper matching approach was applied, whereby for each CAS patient, control subjects with a BMI within ±1 kg/m² were considered eligible. Matching was performed without replacement, allowing each control to be selected only once, but not enforcing a fixed 1:1 matching ratio. As a result, the final matched cohort consisted of 54 CAS patients and 41 control subjects, reflecting a variable matching ratio while maintaining close BMI comparability between groups.

When multiple potential controls were available within the BMI window for a given CAS patient, one was randomly selected. This strategy optimized balance in BMI distribution between groups while preserving statistical power in this exploratory metabolomic analysis.

Age and sex were matched at the cohort design stage, and BMI matching was applied algorithmically as described above.

### Subgroup and exploratory analyses

Predefined exploratory subgroup analyses were conducted according to:

i. disease severity (severe vs. non-severe CAS),
ii. sex (female vs. male).

These analyses were performed to investigate potential effect modification of gut-derived and tryptophan-related metabolites by biological sex, metabolic status, and hemodynamic severity of aortic stenosis. Given the limited sample size, these subgroup analyses were considered hypothesis-generating.

### Multiple testing and interpretation framework

Metabolite concentrations were compared between groups using non-parametric tests. FDR correction for multiple comparisons was applied using the Benjamini–Hochberg procedure. In addition to the conventional significance threshold (q < 0.05), metabolites with FDR-adjusted q values < 0.25 were considered suggestive of potential biological relevance, consistent with exploratory metabolomics studies. Emphasis was placed on consistency across related metabolic pathways rather than on isolated nominal p values.

### Bi-directional Mendelian Randomization

No serum proteins were directly measured in the GUT-CAS cohort; all protein exposures were investigated exclusively via two-sample MR using publicly available GWAS summary statistics as genetic instruments. We conducted a bi-directional, two-sample Mendelian Randomization (MR) study to investigate the causal influence of 18 molecular exposures on the risk of CAS.(22,23) Genetic instrumental variables (IVs) for the molecular exposures were sourced from large-scale Genome-Wide Association Study (GWAS) summary statistics. The protein instruments included IDO1 (prot-a-1410), IL-1beta (GCST004448), IL-6 (GCST90012005)(24), IL-6 Subunit alpha (GCST90012025)(24), IFN-gamma (prot-a-1428)(25), IFN-gamma R1 (prot-a-1430)(25), IL-18 (GCST90012024)(26), IL-18R (prot- a-1491)(26), ALPL (GCST90025947)(27), BMPR1A (prot-a-259)(28), and Osteopontin (GCST90010244)(29). The metabolite and lipid instruments comprised Indole propionate (met-a-475), Indoleacetate (met-a-446), Kynurenine (met-a-375), Tryptophan (met-a-584), Serotonin (met-a-358), LDL cholesterol (GCST90002412), and Lipoprotein(a) (GCST90025993)(30). The outcome data for Aortic Stenosis were obtained from the Kany et al. (2025) harmonized GWAS (Accession: GCST90651074), encompassing 45,352 European cases and 1,938,877 controls. The methodology followed the STROBE-MR guidelines.(31)

### Instrumental Variable Selection and Quality Control

To ensure the validity of the MR assumptions, we selected SNPs associated with each exposure at a genome-wide significance threshold of P < 5 x 10^-8^.(32) To mitigate weak-instrument bias, we calculated the F-statistic for each variant based on the specific sample sizes of the discovery cohorts, retaining only robust instruments with an F-statistic > 10.(33) Genetic independence was ensured through a clumping procedure (r^2^ < 0.001)(34), and all SNPs were cross-referenced with the Kany et al. outcome data using the *hm_rsid* identifiers. Directional consistency was maintained through a harmonization step, wherein effect sizes were aligned or flipped according to the effect alleles of the outcome dataset.

### Statistical Estimation and Sensitivity Analysis

The primary causal effect was estimated using the Inverse Variance Weighted (IVW) method. For exposures with multiple instruments, we assessed heterogeneity using Cochran’s Q statistic; a significant Q-statistic (P < 0.05) prompted the transition from a Fixed-Effects to a Multiplicative Random-Effects IVW model to provide more conservative confidence intervals. Potential horizontal pleiotropy was evaluated via the MR-Egger regression intercept test adding weighted median, and the stability of the causal estimates was validated using a Leave-One-Out (LOO) analysis to identify influential outliers. To confirm the assumed causal direction and prevent bias from reverse causality, Steiger filtering was applied by comparing the variance explained (R^2^) by the SNPs in both the exposure and the outcome.

### Reverse Mendelian Randomization

To assess the possibility of reverse causality—wherein the pathological state of Aortic Stenosis might drive changes in molecular concentrations—a reverse MR analysis was performed. In this phase, significant genetic variants from the Kany et al. (2025) Aortic Stenosis GWAS were utilized as instruments to estimate the causal effect of disease liability on the 18 previously identified proteins and metabolites.(35) All analyses were implemented using a custom Python pipeline incorporating the SciPy and Statsmodels libraries for high-precision statistical computation.

## RESULTS

Participant enrolment and exclusion are summarised in Figure 1.

**Figure 1.**
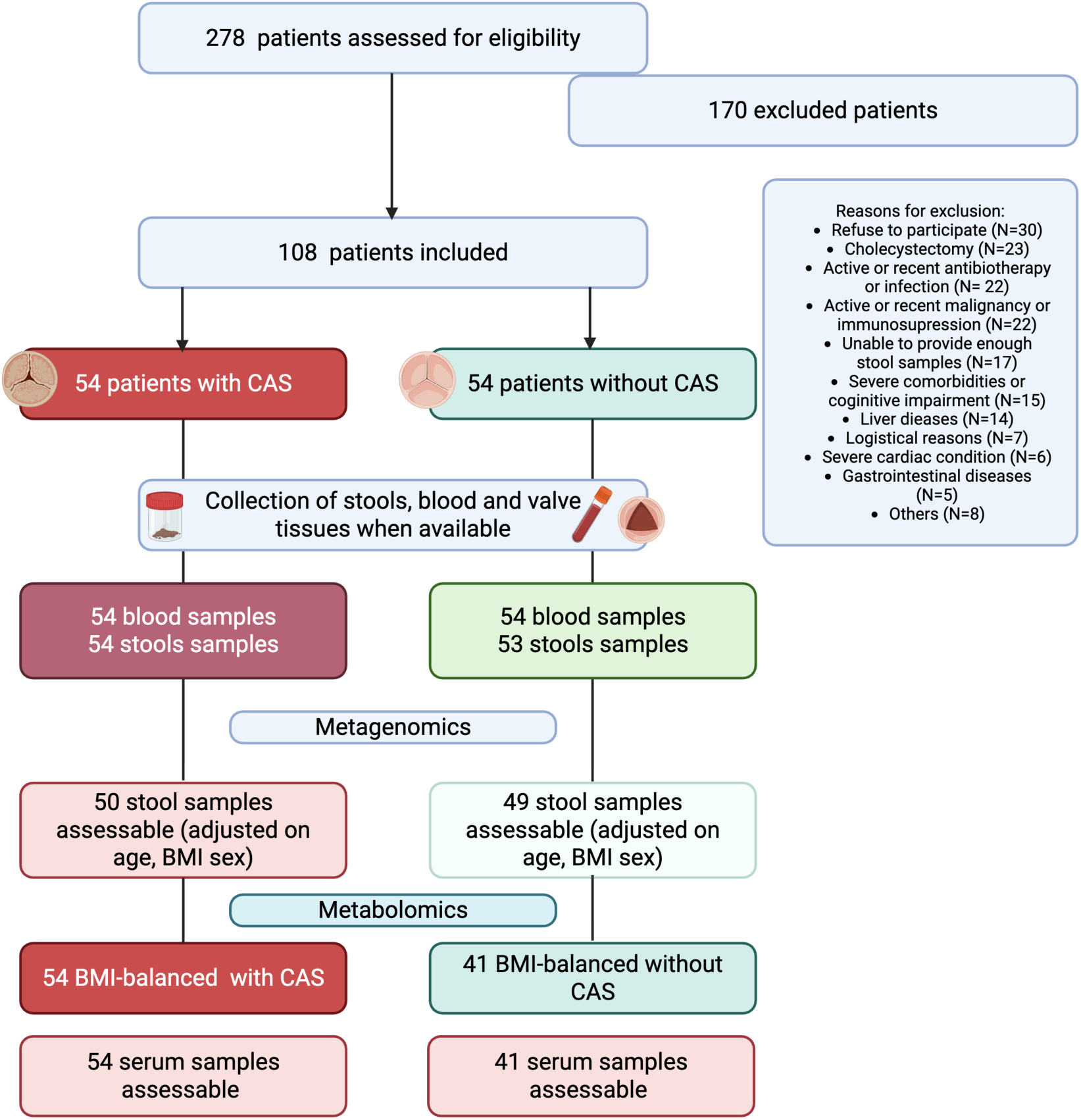
Study flowchart. Of 278 patients assessed for eligibility, 108 were enrolled (54 with calcific aortic stenosis [CAS] and 54 controls without CAS). After BMI matching, 41 controls were retained for the metabolomic analysis. Gut microbiome analysis by 16S rRNA sequencing was performed prior to BMI matching in all participants with available stool samples (n = 50 CAS, n = 49 controls), with BMI, age, and sex included as covariates in the linear model. Serum tryptophan metabolomics was performed in the BMI-matched cohort (n = 54 CAS, n = 41 controls).

### Clinical and biological characteristics of the study cohort

We prospectively enrolled 54 patients with CAS (median age 66.0 years, interquartile range [IQR] 60.0–75.5) and 41 controls without CAS (median age 69.0 years, IQR 60.25–76.0), with no significant difference in age between groups (p = 0.482). The two groups were comparable for sex, body mass index, prevalent coronary artery disease, and conventional cardiovascular risk factors, as well as for inflammatory markers, markers of myocardial injury, and metabolic profiles. C-reactive protein concentrations were low and similar between groups. Some clinically relevant differences were nonetheless present, including the prevalence of atrial fibrillation, anticoagulant use, and diabetes, as well as blood pressure **(Table 1).**

### Gut-derived metabolomic profile in BMI-matched CAS and non-CAS patients

We next examined whether CAS was associated with alterations in gut-derived metabolite profiles. Given the exploratory nature of targeted metabolomics in a cohort of this size, all comparisons are reported with both nominal p-values and Benjamini–Hochberg false-discovery-rate (FDR) q-values, and findings are interpreted as hypothesis-generating. In BMI-matched CAS and non-CAS patients, targeted fasting serum metabolomics suggested a selective shift within tryptophan metabolism, with directionally higher concentrations of several kynurenine-pathway metabolites in CAS. Compared with controls, CAS patients showed nominally higher circulating concentrations of kynurenine (p = 0.078), 3-hydroxyanthranilic acid (p = 0.030), and xanthurenic acid (p = 0.045) (Figure 2A–2D), with quinolinic acid and 3-hydroxykynurenine numerically higher and upstream tryptophan concentrations comparable between groups. After FDR correction, none of the individual metabolites reached the conventional threshold (q < 0.05); 3-hydroxyanthranilic acid, xanthurenic acid, and indole-3-sulfate remained in the suggestive range (q = 0.227 each; q < 0.25) commonly used as a discovery threshold in exploratory metabolomic studies, while kynurenine fell marginally above this threshold (q = 0.254). The full tryptophan-metabolite panel, including substrates unaffected in CAS and all p- and q-values, is summarized in Supplementary Table 2A. In sensitivity analyses in which atrial fibrillation, anticoagulant use, and diabetes were each added individually to a base model adjusted for age and BMI, the CAS effect estimates were stable in direction and magnitude for all kynurenine-pathway metabolites (for example, kynurenine β ≈ 0.11–0.13 and indole-3-sulfate β ≈ −0.24 to −0.27 across models), and no single comorbidity attenuated the associations (Supplementary Table 3; Supplementary Figure 1). Indole-3-sulfate remained nominally significant in every model (p = 0.023– 0.035). These analyses indicate that the tryptophan-kynurenine signal is not explained by the measured differences in comorbidity or medication, while the individual associations remain exploratory and do not survive FDR correction.

**Figure 2.**
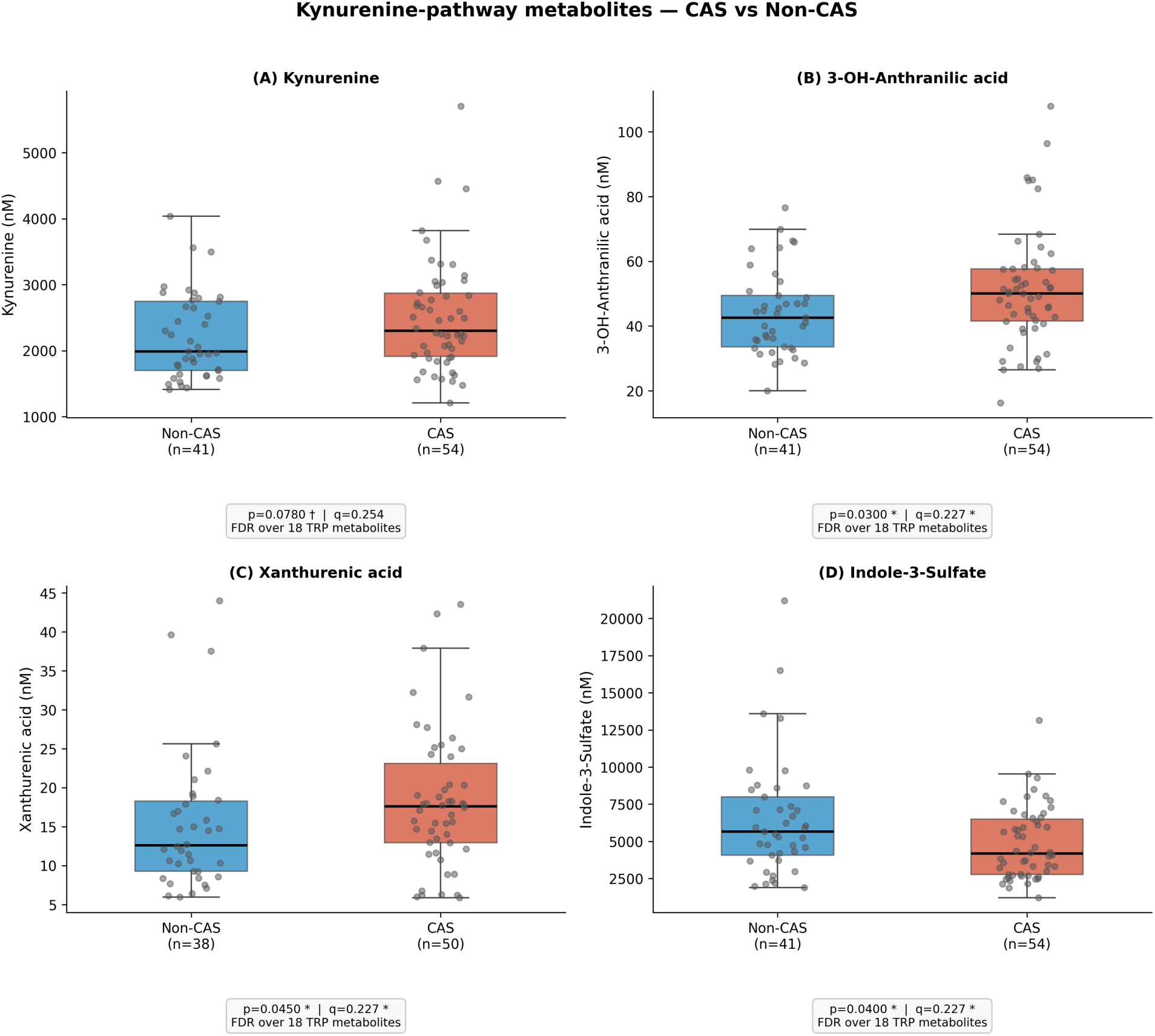
Tryptophan-pathway metabolites in BMI-matched CAS and Non-CAS patients. Boxplots show circulating kynurenine (A), 3-OH-anthranilic acid (B), xanthurenic acid (C), and indole-3-sulfate (D) concentrations in CAS (n = 54) and Non-CAS (n = 41) patients. Differences between groups were assessed using Mann-Whitney U tests with Benjamini-Hochberg FDR correction. *q < 0.05, **q < 0.01. Box: IQR; whiskers: 1.5× IQR.

Non-parametric effect sizes were directionally consistent with these observations. The largest positive Cliff’s δ values were observed for indole-3-acetamide (δ = 0.282), 3-hydroxyanthranilic acid (δ = 0.242), and xanthurenic acid (δ = 0.229), with 5-hydroxyindoleacetic acid showing a small negative effect (δ = −0.198); these values correspond to small-to-moderate effects and reinforce the directional coherence of the kynurenine-pathway signal without substituting for statistical significance. Within the indole pathway, only indole-3-sulfate, a microbiota-derived uremic solute that undergoes renal clearance, was nominally lower in CAS (p = 0.04, Figure 2D), whereas indole-3- propionic acid, indole-3-acetic acid, indole-3-lactic acid, indole-3-aldehyde, and indole-3-acetamide did not differ between groups. Because indole-3-sulfate is renally cleared, we tested whether the observed reduction could reflect differences in renal function rather than microbiota-driven indole production. In a sensitivity model adding eGFR to the base model (CAS + age + BMI), the CAS association with indole-3-sulfate was not attenuated; the effect estimate strengthened (β = −0.278, p = 0.013 vs. β = −0.236, p = 0.035 in the base model), while eGFR itself was independently and inversely associated with indole-3-sulfate (β = −0.176, p = 0.022), consistent with its role as a marker of renal solute handling. Notably, renal function did not differ significantly between groups (eGFR: CAS 91.3 vs. controls 95.0 ml/min/1.73m², p = 0.46), and CAS patients had modestly lower creatinine concentrations (78 vs. 85 µmol/L, p = 0.015), indicating that if anything, the better renal function in CAS would bias toward higher rather than lower indole-3-sulfate clearance. Renal confounding therefore does not explain the observed indole-3-sulfate reduction; the finding is best attributed to reduced microbiota-dependent indole production (Supplementary Table 4A; Supplementary Figure 2). Importantly, indole-3-sulfate levels were not significantly correlated with eGFR in CAS patients (Spearman ρ = −0.18, p = 0.19), in controls (ρ = −0.05, p = 0.77), or in the full cohort (ρ = −0.14, p = 0.18), indicating that the KI ratio elevation was independent of renal function.

Serotonin-pathway metabolites, including serotonin and 5-hydroxytryptophan, were numerically higher in CAS without reaching significance. Because serum serotonin is largely derived from platelet release, we examined whether platelet count confounded any apparent group difference; platelet count was a significant predictor of serum serotonin (β = +0.231, p = 0.003, r = 0.33) but did not differ between CAS and controls (217 vs. 209 × 10⁹/L, p = 0.57), and after adjustment for platelet count the CAS effect on serotonin remained null (β = −0.007, p = 0.96). These analyses confirm that the absence of a significant serotonin difference between groups is not materially influenced by platelet-mediated confounding (Supplementary Table 4B).

Because absolute quantification was performed using stable-isotope dilution, metabolite ratios were analytically interpretable. The kynurenine-to-indole ratio was higher in CAS than in controls (p = 0.0031, q = 0.013 after FDR correction over the four prespecified tryptophan-pathway ratios; **Figure 3A**), the only FDR-significant result among the ratio analyses and consistent with a relative shift toward host kynurenine metabolism at the expense of microbiota-dependent indole production. Leave-one-out sensitivity analysis confirmed that this result was not driven by any single patient: removing any one of the 95 patients left the p-value in the range 0.0015–0.0051, with Cliff’s δ stable at 0.339–0.384 across all removals and no removal pushing p above 0.005 (Supplementary Figure 3). The indole-to-tryptophan ratio was lower in CAS (p = 0.027, q = 0.054; **Figure 3C**), approaching but not reaching the FDR threshold, and corroborating the directional shift away from the indole branch. In contrast, the kynurenine-to-tryptophan ratio did not differ between groups (**Figure 3B**), indicating that upstream tryptophan supply was preserved and that the signal reflected downstream pathway routing rather than global IDO upregulation. The serotonin-to-tryptophan ratio was similarly unchanged (**Figure 3D**), confirming selectivity of the kynurenine-pathway shift. Pathway-level cumulative metabolite scores showed a non-significant tendency toward higher kynurenine-pathway concentrations in CAS (p = 0.144), with no difference in total indole or serotonin scores; because summed concentrations conflate metabolites spanning different concentration ranges, these scores are reported as exploratory in Supplementary Figure 4 rather than as a primary analysis. To further confirm that the kynurenine-pathway activation reflected CAS status rather than adiposity, a direct comparison of CAS-specific and BMI-specific effect sizes across all tryptophan pathway metabolites demonstrated that xanthurenic acid and indole-3-sulfate were significantly associated with CAS but not with BMI, while 3-OH-anthranilic acid showed associations with both, confirming CAS-specificity of the dominant kynurenine-pathway signal (Figure 4). Finally, to determine whether CAS was associated with broader gut-derived metabolic alterations, we measured short-chain fatty acids, bile acids, and TMAO-related metabolites; none differed significantly between CAS and non-CAS patients (Supplementary Table 2B–D). The absence of changes across these canonical classes supports the selectivity of the tryptophan-kynurenine signal rather than a generalized metabolic disturbance.

**Figure 3.**
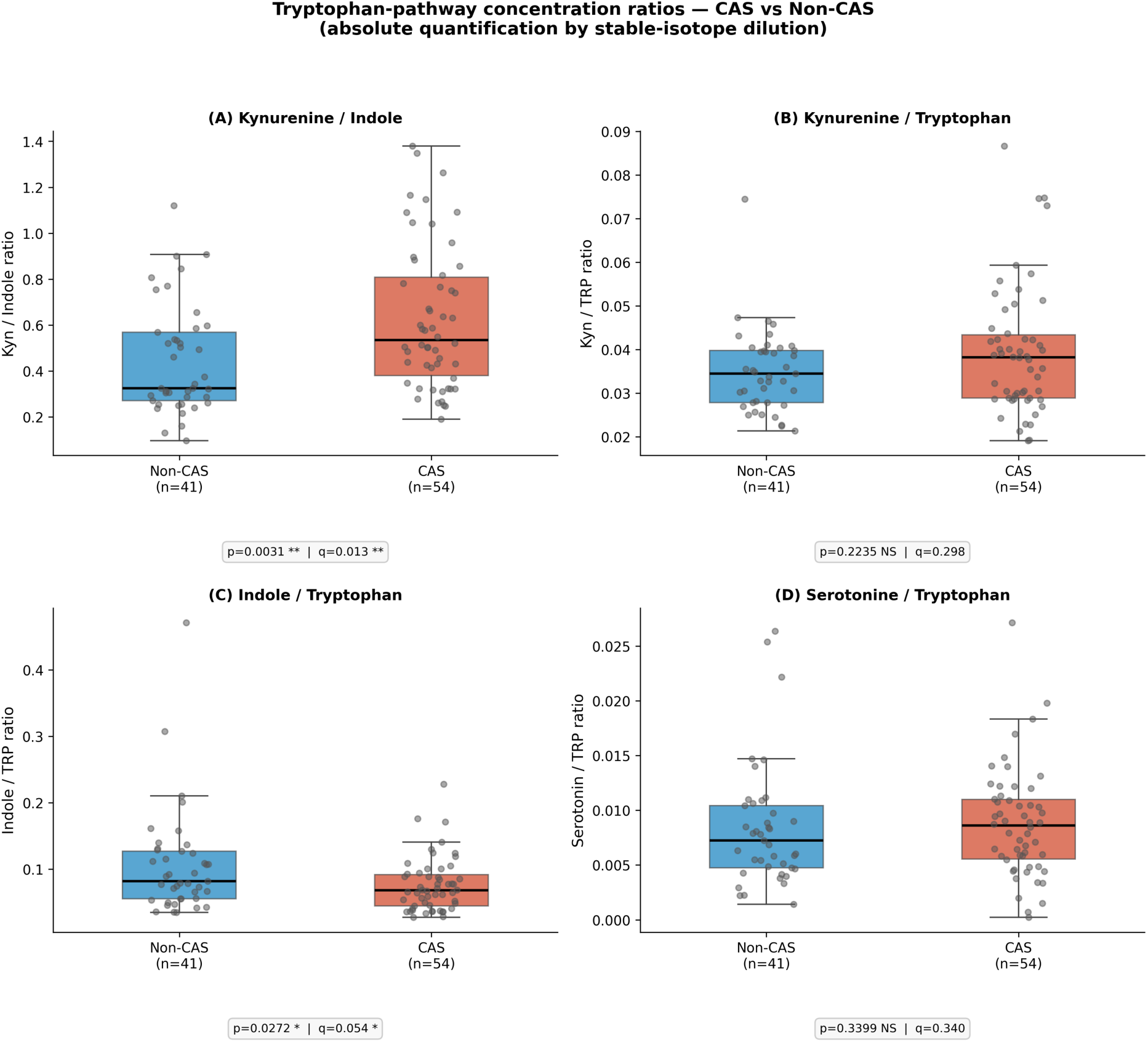
Tryptophan-pathway concentration ratios in BMI-matched CAS and Non-CAS patients. Scatter and violin plots show the kynurenine-to-indole-3-sulfate ratio (KI ratio) and related kynurenine-pathway/indole-pathway ratios in CAS (n = 54) versus Non-CAS (n = 41) patients. Higher KI ratio reflects preferential kynurenine-pathway activation relative to indole-pathway metabolism. Mann-Whitney U test with BH-FDR correction. *q < 0.05, **q < 0.01.

**Figure 4.**
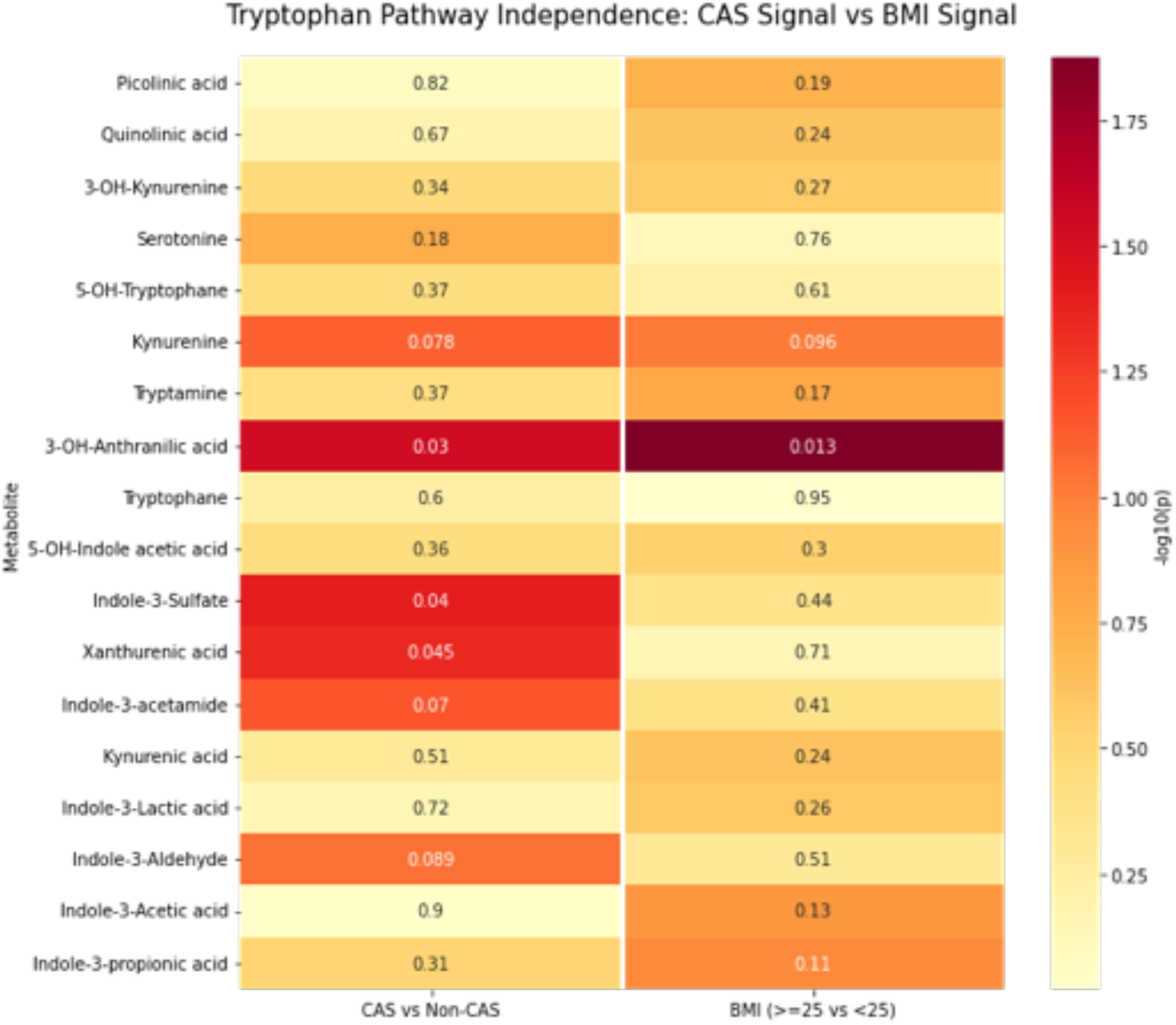
Tryptophan pathway metabolites show CAS-specific signals independent of BMI. Heatmap of Mann-Whitney U test p-values comparing CAS vs. non-CAS (left column) and overweight (BMI ≥ 25) vs. normal-weight (BMI < 25) patients (right column) for each tryptophan pathway metabolite. Color intensity reflects −log₁₀(p); annotated values are raw p-values. Kynurenine-pathway metabolites — including 3-OH-anthranilic acid, xanthurenic acid, and indole-3-sulfate — show nominally significant CAS associations; while 3-OH-anthranilic acid also shows a BMI association (p = 0.013), xanthurenic acid (p = 0.71) and indole-3-sulfate (p = 0.44) remain CAS-specific, confirming that the tryptophan–kynurenine signal is not driven by adiposity. Matched cohort, n = 95.

In a pre-specified sensitivity analysis pooling the KI ratio with all 18 individually quantified tryptophan metabolites (19 tests total under BH-FDR), the KI ratio q-value was 0.059, marginally above the 0.05 threshold. FDR significance for the KI ratio is therefore conditional on the prespecified four-ratio analytical family, consistent with standard practice for targeted metabolomics studies with a priori metabolite groupings. Sex-stratified analyses are detailed in the sex-specific section below; the CAS main effect on kynurenine was independent of sex in adjusted models. To confirm independence from demographic confounding, multivariable logistic regression adjusting for sex, age, and BMI was performed (n = 95). The KI ratio remained independently associated with CAS status after adjustment (OR = 2.49, 95% CI 1.34–4.62, p = 0.004). Indole-3-sulfate also showed an independent inverse association with CAS (OR = 0.56, 95% CI 0.32–0.99, p = 0.046), consistent with the directional shift away from indole-branch production observed in the univariable analyses. Although IDO activity has been linked to lipid status, LDL cholesterol and total cholesterol were essentially identical between groups in the current study (CAS 2.24 vs. controls 2.26 mmol/L), arguing against lipid-driven confounding of the kynurenine finding.(36) Formal correlation analysis within CAS patients confirmed the absence of significant associations between the KI ratio and LDL cholesterol, total cholesterol, or triglycerides (all |ρ| < 0.23, all p > 0.09; Supplementary Figure 7), providing quantitative corroboration of this observation.

### Gut microbiota composition in CAS patients compared to controls

To determine whether these metabolomic alterations were accompanied by structural changes in the gut microbiota, we analyzed microbial composition by 16S rRNA sequencing. Alpha diversity, assessed by the Shannon and Chao1 indices, did not differ between CAS and non-CAS patients (all p > 0.05, **Figure 5A–B**), and beta diversity showed no separation according to CAS status (PERMANOVA, **Figure 5C**). CAS was therefore not associated with global gut microbial differences.

**Figure 5.**
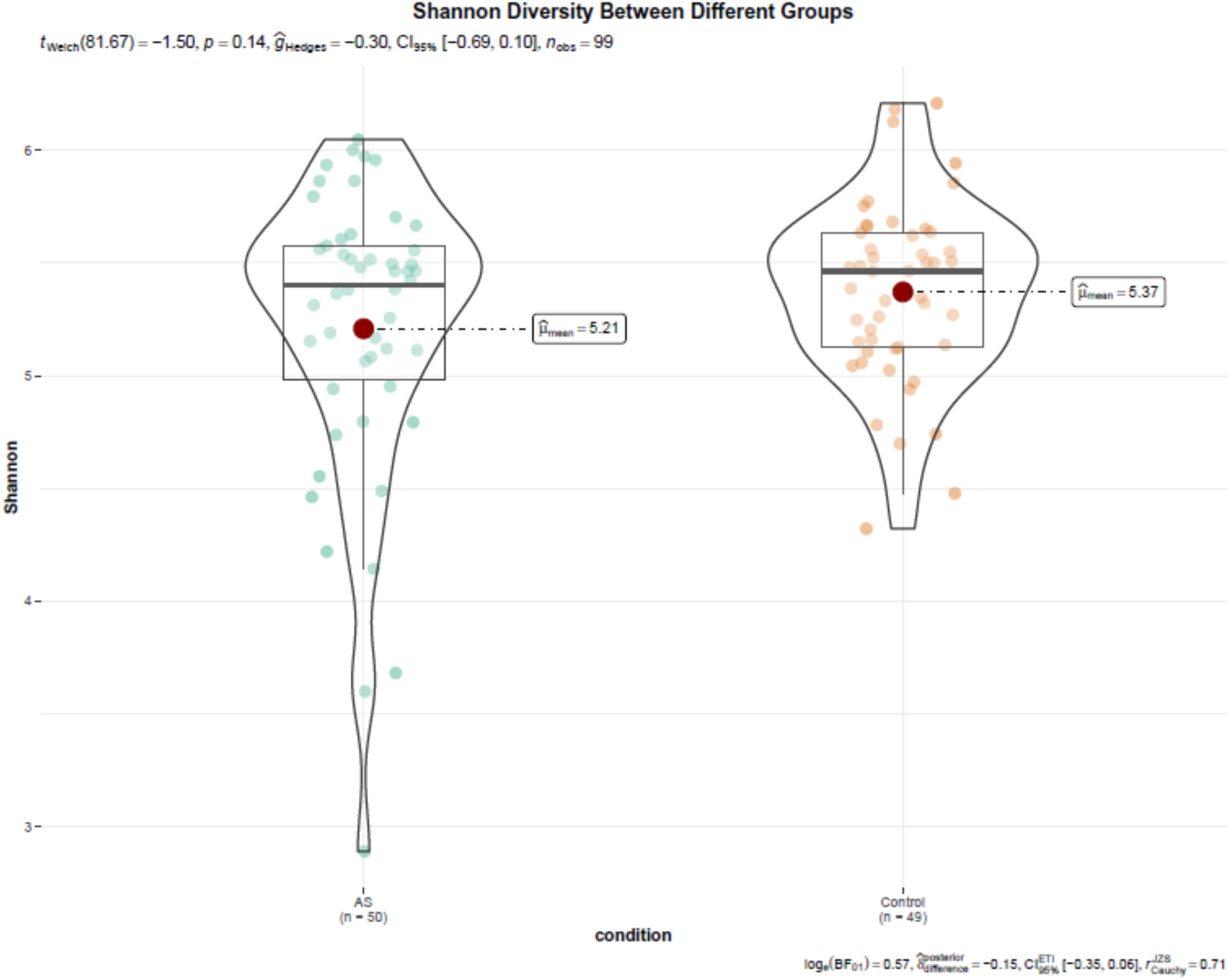
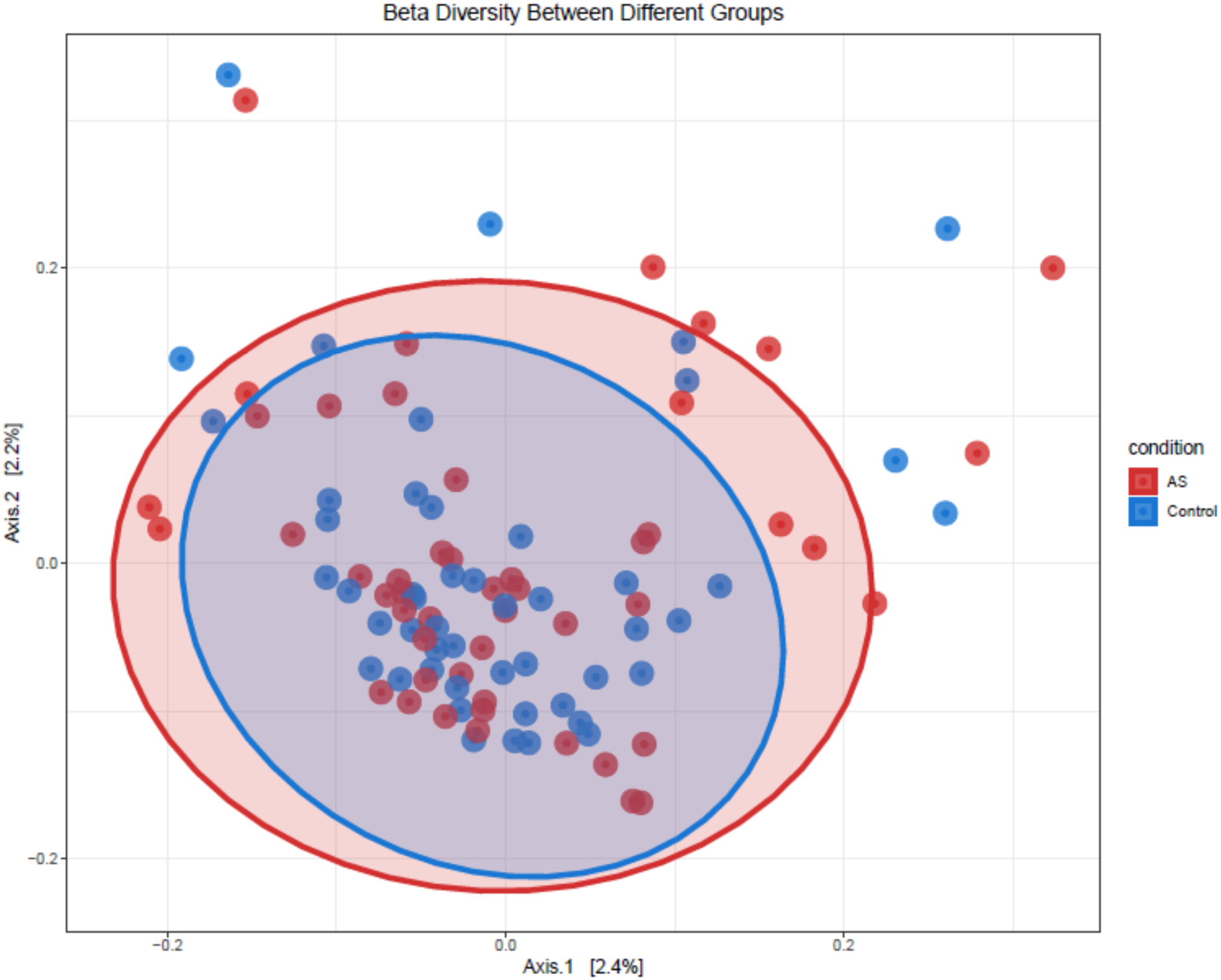
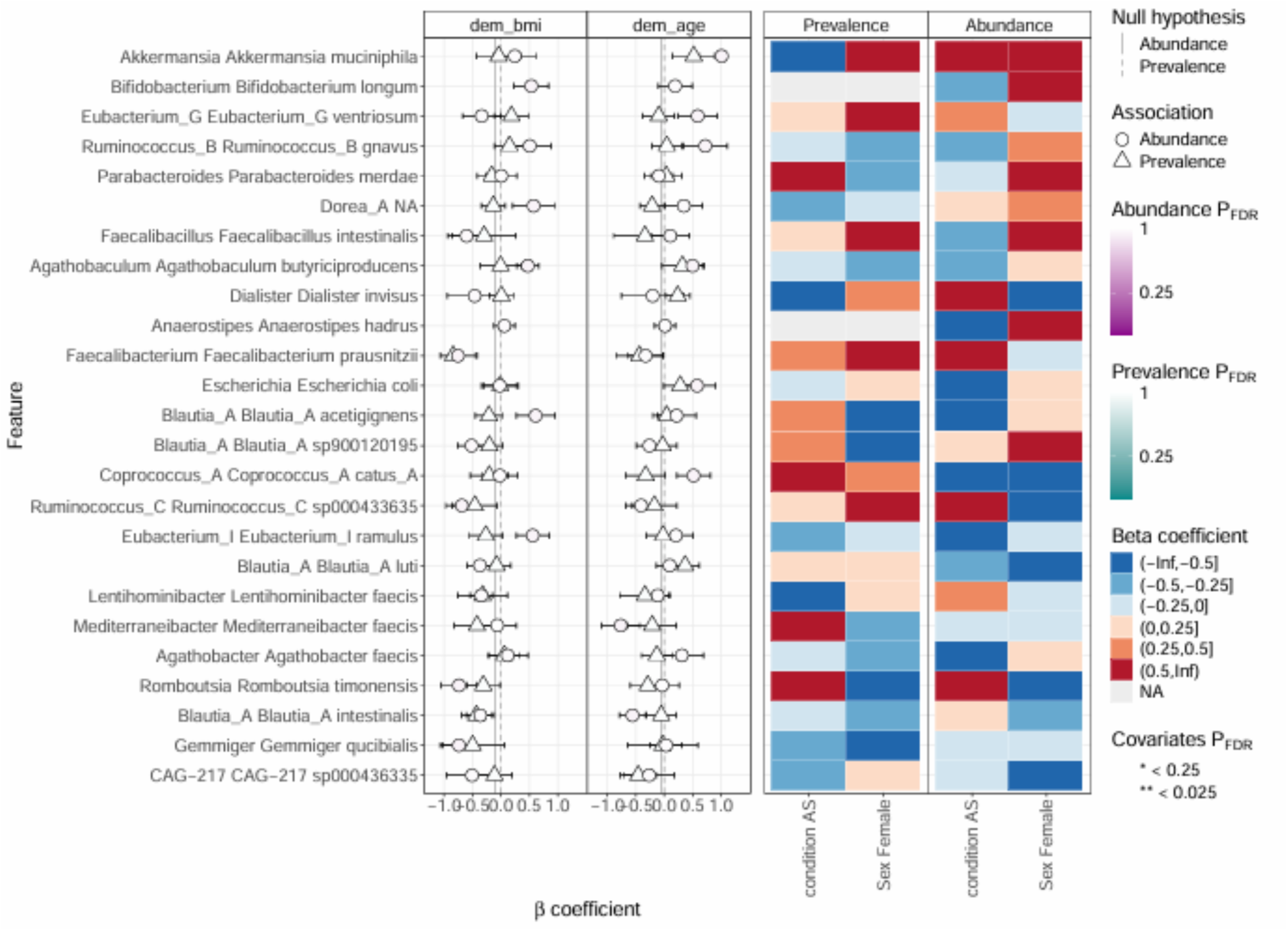
Shannon alpha diversity in CAS and Non-CAS patients. Violin and box plots show comparable within-sample microbiome diversity (Shannon index) between CAS and Non-CAS patients, indicating no global reduction in alpha diversity. The second panel shows beta-diversity (Bray-Curtis dissimilarity) ordination. Statistical differences were assessed by Mann-Whitney U (alpha diversity) and PERMANOVA (beta diversity). The last panel shows the taxonomic associations comparing CAS versus non CAS patients.

At the taxonomic level, multivariable differential abundance analysis adjusted for age and BMI identified a focused pattern of nominally reduced abundance among specific taxa within the Bacillota (formerly Firmicutes) and Actinomycetota phyla. The most consistently depleted genera were *Blautia* and *Holdemanella*, together with Hominilimicola fabiformis, and several additional members of the Lachnospiraceae family, including *Agathobacter* and *Anaerostipes*, were nominally reduced in CAS. None of these associations remained significant after Benjamini–Hochberg correction (Figure 5D), and the coherent depletion of multiple Lachnospiraceae members is therefore best interpreted as a selective, hypothesis-generating signal rather than a study-wide significant microbial signature. Nominally, *Blautia* and *Agathobacter* were less abundant in CAS patients; however, these differences did not survive BH-FDR correction and should be interpreted as hypothesis-generating signals only.

### Metabolite profiles according to CAS severity

After simultaneous adjustment for age and BMI, prespecified as the primary confounding model given that BMI was the primary matching criterion, the KI ratio was not significantly associated with indexed aortic valve area (partial Spearman, p = 0.111, n = 46). In the unadjusted analysis, the KI ratio was inversely correlated with indexed AVA (Figure 6; Spearman ρ = −0.310, 95% CI [−0.540, −0.022], p = 0.036, n = 46), and this association was directionally consistent across individual single-covariate sensitivity models but was attenuated by BMI adjustment. In contrast, the KI ratio did not correlate significantly with mean transvalvular gradient (ρ = +0.047, p = 0.74), which is expected given the greater sensitivity of indexed AVA to structural valve severity. Individual kynurenine-pathway metabolites (kynurenine, 3-OH-anthranilic acid, xanthurenic acid) were not significantly correlated with indexed AVA when examined alone, indicating that the severity signal is captured by the ratio rather than any single metabolite. The inverse correlation between the KI ratio and indexed AVA remained significant after adjusting individually for age, sex, eGFR, atrial fibrillation, diabetes, statin use, and anticoagulant therapy (all partial ρ −0.29 to −0.31, p < 0.05; Supplementary Figure 5), with only BMI showing marginal attenuation (partial ρ = −0.244, p = 0.107), confirming robustness to the main clinical confounders (Supplementary Table 5).

**Figure 6.**
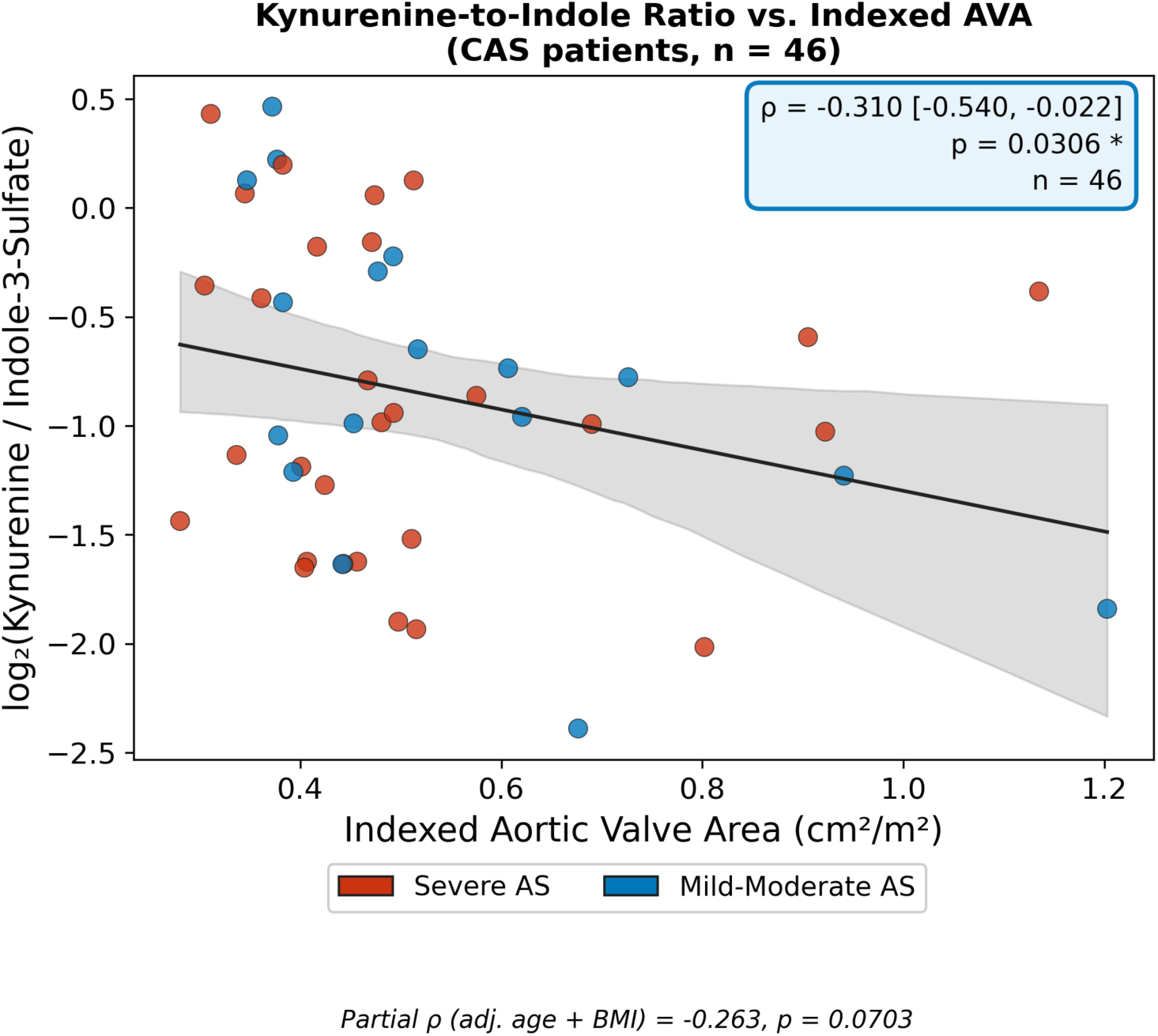
Inverse association between the kynurenine-to-indole ratio and indexed aortic valve area in CAS patients. Scatter plot showing the Spearman correlation between the log₂ kynurenine-to-indole-3-sulfate ratio (KI ratio) and indexed aortic valve area (cm²/m²) in CAS patients with available echocardiographic data (n = 46). Each point represents one patient. The regression line is shown with 95% confidence interval. Spearman ρ = −0.31, 95% CI [−0.54, −0.02], p = 0.031. A lower indexed AVA reflects more severe structural stenosis, suggesting that higher KI ratio is associated with greater valve disease severity.

In the categorical severity comparison, patients with severe CAS (N = 32) had nominally higher kynurenine (p = 0.031, q = 0.464) and quinolinic acid (p = 0.045, q = 0.464) compared with mild-to-moderate disease (N = 16), and propionic acid tended to be lower (p = 0.056); none survived FDR correction (Supplementary Table 6). Total and LDL cholesterol were higher in the severe group, a potential confounder not adjusted for in this comparison, while age, BMI, blood pressure, and comorbidity prevalence were comparable (Supplementary Table 7). Bile acids, short-chain fatty acids, and TMA/TMAO-related metabolites were similar across severity groups. This categorical directional pattern is consistent with the continuous correlation above and with the overall CAS-versus-control direction, but both analyses are exploratory and require confirmation in larger cohorts.

In an exploratory comparison of non-severe CAS (N = 16) with age-, sex-, and BMI-matched controls (N = 13), tryptophan metabolism was largely preserved, with a nominal increase in indole-3- acetamide (p = 0.029) and a consistent but non-significant rise in kynurenine-pathway metabolites (Supplementary Table 6). Given the small sample, this observation is hypothesis-generating only, but it is compatible with kynurenine-pathway activation being a feature of more advanced disease rather than an early event.

### Sex-specific metabolic and microbial signatures in CAS

Baseline characteristics were largely comparable between male (N = 41) and female (N = 13) CAS patients. Women had lower hemoglobin (p = 0.038) and higher total and HDL cholesterol (p = 0.041 and p = 0.025), with a smaller absolute aortic valve area (p = 0.009) but similar indexed valve area, indicating comparable disease severity (Supplementary Table 8). The small number of female patients limits all sex-stratified analyses, which are reported as exploratory.

Sex-stratified comparisons with sex-matched controls confirmed that the kynurenine-pathway signal is a CAS-associated finding independent of sex: the CAS main effect on kynurenine persisted after adjustment for sex, age, and BMI (β = 0.158, p = 0.019), and a formal CAS × sex interaction term did not reach significance for any metabolite (best, kynurenine p = 0.083). In the sex-stratified comparisons, male CAS patients showed nominally higher 3-hydroxyanthranilic acid (p = 0.009, q = 0.185), kynurenine (p = 0.022, q = 0.185), and xanthurenic acid (p = 0.026, q = 0.185) versus male controls, with 3-hydroxyanthranilic acid representing the strongest single-metabolite signal in the study. Among female CAS patients, no kynurenine-pathway metabolite reached nominal significance versus female controls; the CAS × sex interaction analysis indicates this reflects the limited power of the female subgroup (n = 13) rather than a true sex-specific absence of the signal.

Within CAS patients, women and men differed in their tryptophan profiles. Women exhibited higher indole-3-acetamide (p = 0.014, q = 0.135) and lower kynurenine-pathway metabolites, including kynurenic acid (p = 0.015, q = 0.135), along with reduced picolinic acid (p = 0.035, q = 0.202) and indole-3-lactic acid (p = 0.045, q = 0.202) (Figure 7A–D). No metabolite survived FDR correction at q < 0.05, and these intra-CAS differences should be interpreted alongside the small female sample. A summary of all four sex-by-disease contrasts across the relevant tryptophan metabolites is shown in Figure 7E. Full metabolite-level p- and q-values across all sex-stratified CAS vs. control comparisons are provided in Supplementary Table 9.

**Figure 7.**
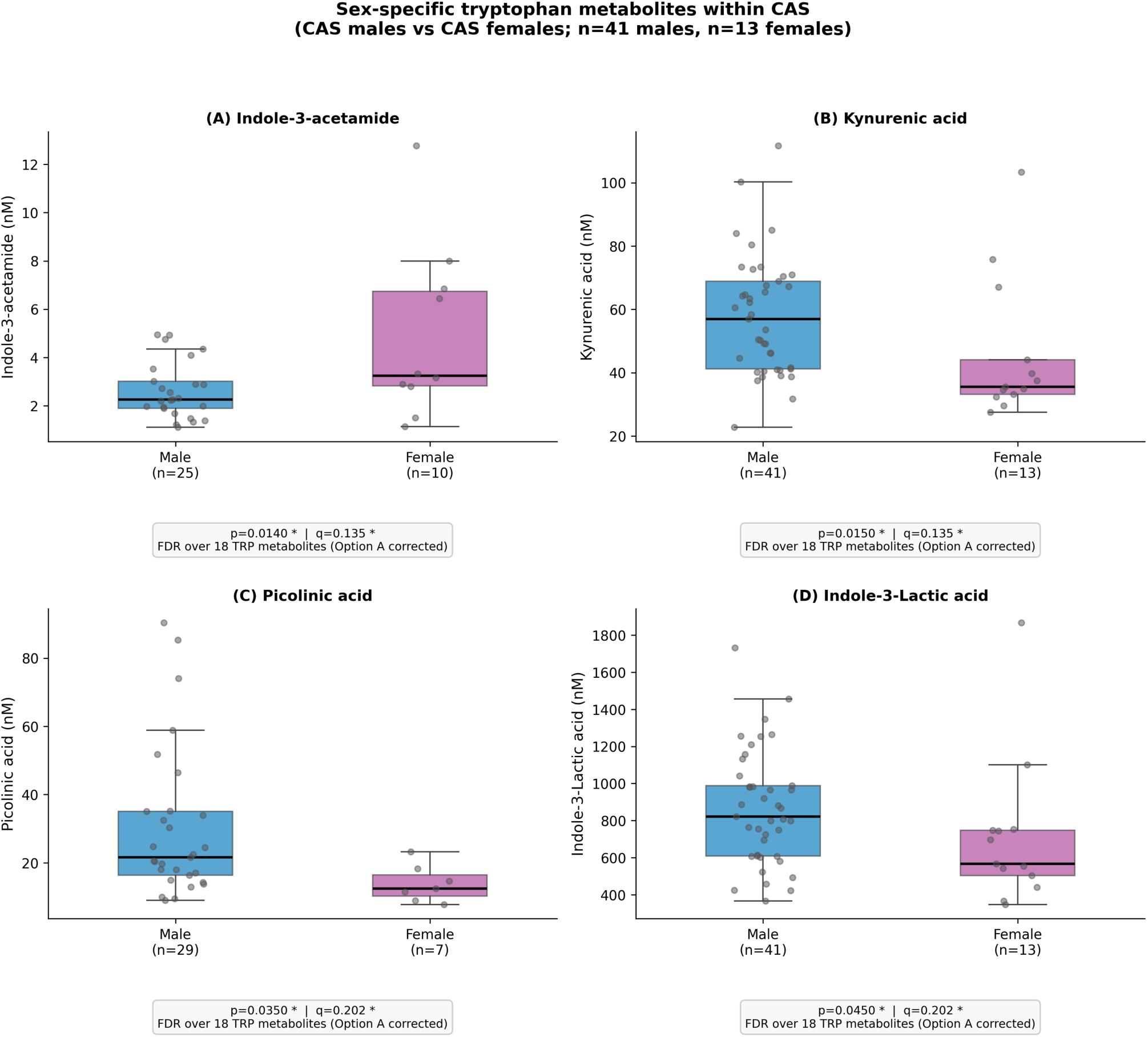
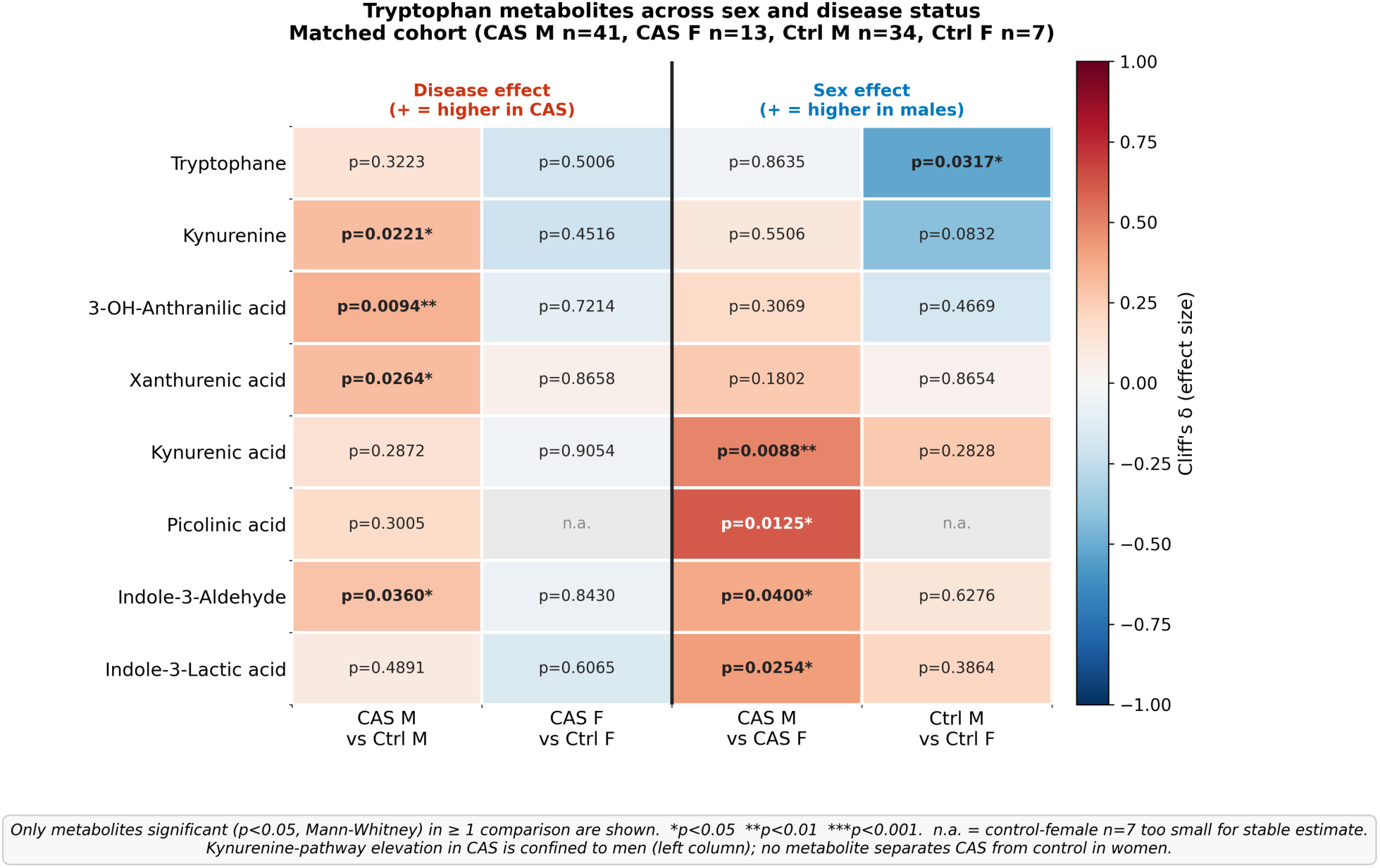
Sex-specific tryptophan-metabolite patterns in CAS and matched controls. (A–D) Selected tryptophan-pathway metabolites showing significant differences between CAS and Non-CAS patients in sex-stratified analyses. Boxplots compare female CAS vs. female controls and male CAS vs. male controls. (E) Heatmap summarizing all tryptophan metabolite comparisons by sex, with color indicating statistical significance (−log₁₀ q-value). BH-FDR correction applied within each sex stratum. *q < 0.05.

Microbial diversity did not differ between male and female CAS patients (Supplementary Figure 6 (panels A–C)). Differential abundance analysis showed higher Bifidobacterium and Lactobacillus in women and enrichment of taxa linked to cardiometabolic risk in men. Finally, a modest difference in microbial community structure was observed in women according to CAS status (female CAS vs. female controls; PERMANOVA R² = 0.051, p = 0.046), indicating that CAS status explained only about 5% of community variance; this marginal result is reported as exploratory.

### Mendelian Randomization

We finally performed MR to assess the potential causal role of inflammatory and metabolic pathways in CAS, using genetic variants as proxies for lifelong exposure to circulating biomarkers (**Figure 8A–B; GWAS sources in Table 2**).

**Figure 8.**
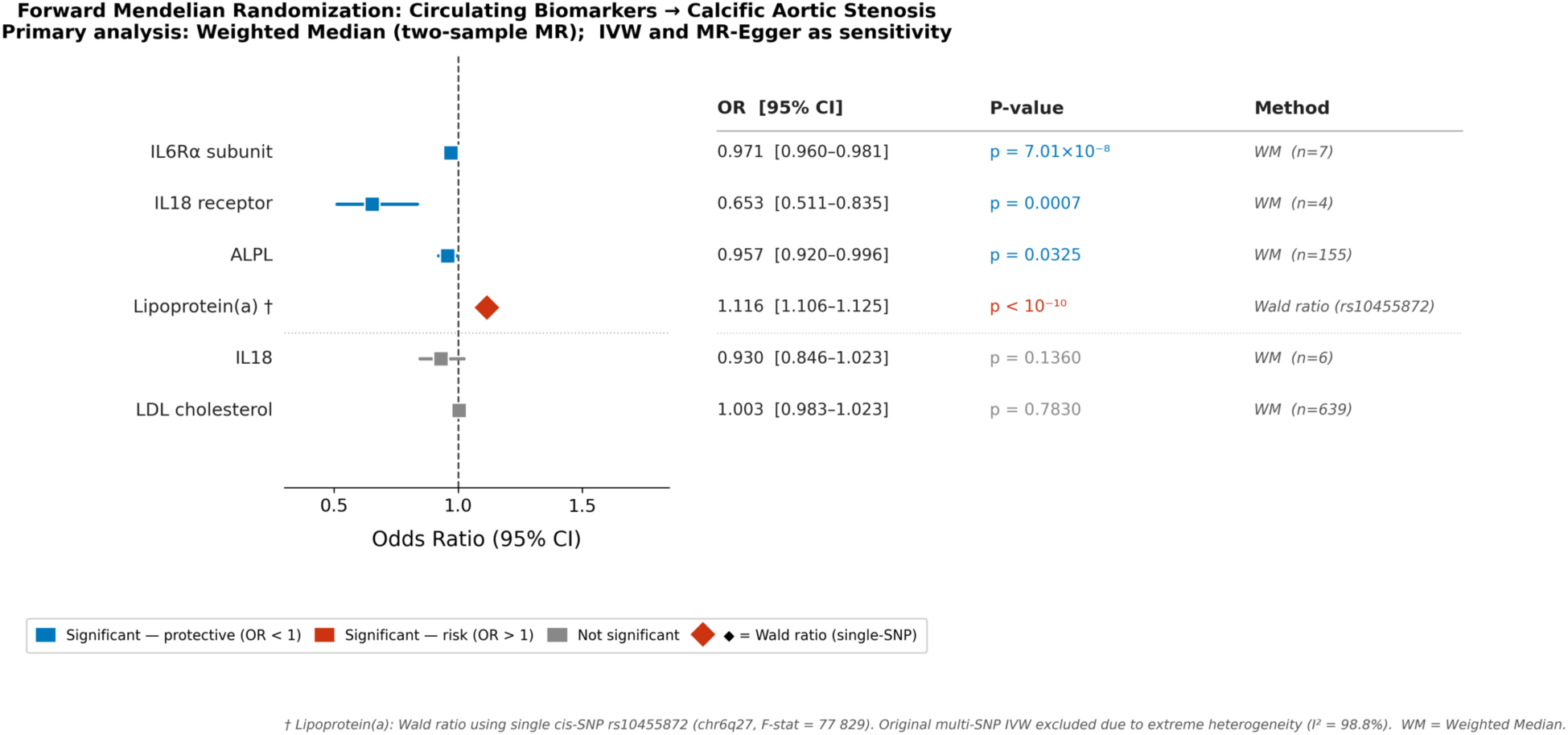
Direct Mendelian randomization estimates for circulating biomarkers and calcific aortic stenosis risk. Forest plot of two-sample Mendelian randomization estimates for the effect of six circulating biomarkers — IL-6Rα subunit, IL-18 receptor, ALPL, lipoprotein(a), IL-18, and LDL cholesterol — on CAS risk (odds ratio per 1-SD genetically instrumented increase in each biomarker). Weighted median was the primary method, with inverse-variance weighted (IVW) and MR-Egger regression as sensitivity analyses; for lipoprotein(a), a single-SNP Wald ratio (rs10455872) was used because the multi-SNP instrument showed extreme heterogeneity (I² = 98.8%). Point estimates with 95% confidence intervals and p-values are shown. MR analyses used summary statistics from the largest available GWAS for CAS. Genetically higher IL-6Rα subunit (OR 0.97), IL-18 receptor (OR 0.65), and ALPL (OR 0.96) were associated with lower CAS risk, whereas higher lipoprotein(a) (OR 1.12) was associated with higher CAS risk; IL-18 and LDL cholesterol were not significant.

In the forward direction (exposure → CAS; Supplementary Table 11), lipoprotein(a) was positively associated with CAS risk (Wald ratio ORi=i1.116 [1.106–1.125], pi<i0.001; single-SNP instrument rs10455872 used due to harmonization failure of the multi-SNP instrument; Supplementary Table 10), consistent with prior observational evidence. In contrast, LDL cholesterol showed no significant causal effect on CAS. Genetically predicted IL6 receptor α subunit (IL6Rα), IL18 receptor, and ALPL were each inversely associated with CAS risk (effect estimates below unity). Notably, receptor-level markers (IL6Rα and IL18R), rather than circulating cytokines, were associated with CAS.

In the reverse direction (CAS → biomarkers; Supplementary Table 12), genetically predicted CAS was associated with downstream inflammatory pathways, including interferon signaling (IFN and IFNγ receptor 1) and IL18, and with multiple tryptophan metabolites; kynurenine and indole derivatives were significantly associated with genetic liability to CAS. Kynurenine showed a negative genetic effect estimate in reverse MR despite being elevated in the circulation of CAS patients. Rather than a contradiction, this discordance is consistent with a model in which circulating kynurenine reflects downstream, disease-driven inflammatory amplification rather than a primary causal driver, with the genetic estimate capturing a distinct regulatory layer. Robustness analyses supported the stability of these findings: all instruments showed strong F-statistics (> 10), and IVW, weighted-median, MR-Egger, and leave-one-out analyses were concordant for both directions (Supplementary Tables 13– 20). Associations were not driven by single variants, with the strongest robustness for ALPL and IFN-related pathways, while inflammatory and tryptophan-related markers showed greater variability consistent with their downstream nature. Steiger filtering (Supplementary Tables 21–22) supported correct directionality in forward MR and showed mixed directionality for inflammatory pathways in reverse MR, particularly IL6 and IL18 signaling, consistent with a bidirectional relationship between systemic inflammation and CAS. Taken together, tryptophan metabolites showed no evidence of forward causal effects but significant reverse-MR associations, supporting their role as downstream markers of CAS-related inflammatory activation rather than primary causal drivers.

## DISCUSSION

In this integrated microbiome and metabolomic study of patients with calcific aortic stenosis, we report three principal findings. First, CAS was associated with a selective alteration of tryptophan metabolism characterized by elevated kynurenine-to-indole-3-sulfate ratio and a relative shift away from indole-derived metabolites. Second, these metabolic changes occurred in the context of only modest alterations in specific gut microbial taxa and in the absence of broad perturbations in other major gut-derived metabolite classes, including short-chain fatty acids, bile acids, and TMAO-related metabolites, arguing against generalized gut microbial differences. Third, the kynurenine-to-indole ratio was associated with structural disease severity, as reflected by aortic valve area, linking this immunometabolic signature to the burden of valvular disease. Collectively, these findings are consistent with the concept of a focused gut–host immunometabolic remodeling in CAS and identify the tryptophan–kynurenine axis as a potential pathway of interest in valvular disease biology.

### Kynurenine pathway metabolites elevated in calcific aortic stenosis

The present study reports evidence of a shift of tryptophan metabolism toward the kynurenine pathway across increasing CAS severity, with higher circulating kynurenine and quinolinic acid, supporting activation of IDO 1, a key enzyme induced by interferon-γ and other pro-inflammatory cytokines.(37) This pattern is consistent with chronic low-grade inflammation and immune activation and has been extensively described in atherosclerosis, heart failure, and valvular heart disease.(38–40) Experimental and clinical data indicate that IDO 1 activity is increased in inflammatory cardiovascular conditions and promotes endothelial dysfunction, oxidative stress, and vascular remodeling.(41) In the aortic valve, inflammatory cell infiltration precedes and accompanies calcification, with macrophage-derived cytokines driving osteogenic differentiation of VICs.(42) Kynurenine and its downstream metabolites, particularly quinolinic acid, have been shown to enhance reactive oxygen species production and activate NMDA-receptor–dependent pathways, further amplifying local inflammatory and calcific signaling.(43) Our findings of a potential elevated kynurenine-to-indole-3-sulfate ratio in patients with CAS, particularly in association with sex and BMI, fit well with the current concept of CAS as an inflammation-driven fibro-calcific disease. Transcriptomic profiling of human aortic valves across the spectrum of disease severity has demonstrated that immune and inflammatory pathways are among the earliest and most strongly up- regulated biological programs in CAS, with a progressive enrichment of adaptive immune responses, B-cell signaling, and pro-inflammatory cytokine networks as stenosis severity increases.(44) In this study, genes involved in interleukin signaling, including IL6 and IL1B, and cytokine such as IFNγ, were markedly overexpressed in severe and early-onset CAS, highlighting a chronic low-grade inflammatory milieu within the valve.(45,46) Such cytokines are known inducers of IDO 1 activity,(47)

Oxidative stress is a central driver of calcification through activation of the NOTCH1–Wnt/β-catenin– BMP2–Runx2 axis and mitochondrial dysfunction processes that are themselves amplified by inflammatory cytokines and kynurenine pathway metabolites.(48,49) In parallel, systemic inflammatory profiling in severe AS has revealed sex-specific cytokine signatures associated with fibrocalcific burden, including IL-8, TGF-α, GM-CSF, IP-10, and IL-10, supporting the concept that chronic immune activation and immune-metabolic crosstalk differ between men and women.(30) In contrast, kynurenine has been shown to exert a protective role in arterial calcification models, highlighting that the functional consequences of kynurenine elevation may be tissue- and context-specific: the predominantly fibro-calcific osteoblastic process driving valvular calcification may respond differently from the atherosclerosis-driven mechanisms underlying arterial calcification.(50)

In our study, we observed a trend toward higher levels of kynurenine pathway metabolites even in CAS patients with early-stage disease, although these differences did not reach statistical significance. This finding may reflect the limited sample size and reduced statistical power, but it aligns conceptually with the notion of early low-grade inflammation preceding overt valve calcification. Similar to observations from 68Ga-DOTATATE PET/CT studies, which demonstrated that increased valvular uptake, a surrogate of macrophage and pericyte activation, is associated with baseline aortic valve calcification and predicts disease progression, our results suggest that inflammatory processes are active even before substantial calcific burden is present.(51)

### Immune-tryptophan metabolism reprogramming in calcific aortic stenosis

Mendelian randomization (MR) suggests a potential involvement of the kynurenine pathway in a broader disease process, potentially driven by IFN-γ–mediated activation of IDO. This could favor the degradation of tryptophan toward kynurenine at the expense of alternative pathways, including the production of protective indole derivatives by the gut microbiota.^9^/16/2026 3:43:00 AM

Consistently, while the classical kynurenine/tryptophan ratio was not significantly altered in our cohort, the kynurenine/indole ratio was markedly increased, suggesting a shift in tryptophan partitioning rather than isolated host pathway activation. This observation is consistent with the concept of “interkingdom” competition for tryptophan, whereby gut bacteria and the host directly compete for substrate availability.(52) In this context, the elevated kynurenine/indole may reflect a dual process involving increased host-driven kynurenine production and impaired microbial utilization of tryptophan.

A potential microbial contribution is suggested by the depletion of key indole-producing taxa, particularly within the *Lachnospiraceae* family,(53) including *Blautia* and *Agathobacter*. Notably, IS was the only indole-derived metabolite significantly reduced, whereas other derivatives showed neutral or compensatory trends. This selective depletion may suggest a disruption within microbial tryptophan metabolism, with reduced flux through the indole-to-IS pathway and consequent loss of ligands for the aryl hydrocarbon receptor.^8^ Although IS is considered a pro-calcific uremic toxin at high concentrations, physiological levels in non-renal settings are increasingly recognized as essential for maintaining gut barrier integrity and limiting systemic inflammation^53^. Its depletion may therefore reflect a loss of key anti-inflammatory regulatory signals. Furthermore, kynurenine has been classified as a uremic toxin with endothelial cytotoxic properties; while severe uremia was excluded in the current cohort (eGFR 94.5 vs. 87.3 ml/min/1.73m² in CAS and controls respectively), this endothelial toxicity pathway represents a plausible mechanistic link between elevated circulating kynurenine and valve endothelial dysfunction.(55)

At the tissue level, ongoing valvular inflammation, immune cell infiltration, and calcific remodeling may increase local metabolic demand, while systemic inflammatory signaling continues to drive kynurenine-to-indole-3-sulfate ratio elevation.(56) This imbalance may be consistent with a state of metabolic disequilibrium or “metabolic steal”, in which sustained pathway activation exceeds local regulatory capacity, promoting accumulation of kynurenine metabolites and depletion of protective indole compounds.(57) These processes may contribute to a more pro-inflammatory, pro-oxidative, and pro-calcific environment.

Bidirectional MR analyses were consistent with shared inflammatory pathways influencing CAS risk (forward MR) and CAS genetic liability associating with altered circulating tryptophan metabolite levels (reverse MR), suggesting a complex bidirectional or reactive relationship rather than simple causal directionality. The negative genetic effect estimate for kynurenine in the reverse direction is consistent with a model in which circulating kynurenine reflects disease-driven inflammatory amplification rather than constituting a primary upstream driver; the strong reverse-MR associations with IFN signaling further support CAS-related immune activation as a driver of kynurenine pathway upregulation.

These findings are compatible with a hypothesis in which CAS evolves from an initial lipid-driven valvular trigger into a state characterized by persistent crosstalk between inflammatory activation and metabolic reprogramming. Within this framework, the kynurenine pathway may represent a potential interface linking immune activation, microbial ecology, and tryptophan metabolism, and its sustained activation could contribute to the pro-inflammatory and pro-oxidative environment of the stenotic valve. The inverse correlation between the KI ratio and indexed aortic valve area (ρ = −0.310, p = 0.031), robust to individual adjustment for age, sex, eGFR, atrial fibrillation, diabetes, statin use, and anticoagulant therapy, is consistent with a graded relationship between tryptophan pathway routing and structural severity, though causality cannot be inferred from these cross-sectional data.

These interpretations remain associative and hypothesis-generating; no causal relationship was demonstrated in Mendelian randomization analyses, where kynurenine appeared as a downstream marker of CAS-related inflammation rather than an upstream driver. Prospective and mechanistic studies in adequately powered cohorts are needed to determine whether targeting this pathway could modify valvular disease progression.

### Sex dimorphism in tryptophan metabolism

We observed exploratory signals suggesting potential sex-related differences in tryptophan metabolism, with male CAS patients exhibiting higher circulating kynurenine and downstream metabolites than females. This observation is broadly consistent with previous reports indicating that men and women display distinct immune–metabolic profiles and valvular remodeling patterns in calcific aortic stenosis, with men generally developing greater calcific burden and women exhibiting a more fibrotic phenotype. (58,59) Sex-associated differences in gut microbial composition and inflammatory signaling have also been described and may influence tryptophan metabolism and kynurenine-pathway activation. (60–62) In this context, the higher kynurenine pathway activity observed in male CAS patients may reflect differences in systemic immune activation and host– microbiome interactions. However, the CAS-by-sex interaction did not reach statistical significance, and the study was not powered for sex-stratified analyses. These findings should therefore be considered hypothesis-generating and require confirmation in larger, independent cohorts.

### Limitations

Several limitations should be acknowledged. First, this was an exploratory single-center study with a modest sample size and no independent replication cohort. Consequently, subgroup analyses should be considered hypothesis-generating, and confirmation in larger, independent populations is required.

Second, microbiome profiling was performed using full-length 16S rRNA gene sequencing rather than shotgun metagenomics, limiting strain-level resolution and direct functional pathway characterization.

Third, despite matching for age, sex, and body mass index, residual confounding cannot be excluded. Differences in clinical characteristics, comorbidities, medication use, lifestyle factors, and other unmeasured variables may have influenced both gut microbial composition and circulating metabolite levels. Of note, antidiabetic medications — in particular metformin — independently alter gut microbial composition and tryptophan metabolism although sensitivity analyses adjusting individually for diabetes status did not substantially attenuate the primary associations, a residual effect of antidiabetic therapy cannot be fully excluded.(63)

Fourth, serotonin was measured in serum rather than plasma. Because serum serotonin is substantially influenced by platelet release during coagulation, future studies should employ plasma-based measurements to more accurately assess circulating serotonin concentrations.

Fifth, the cross-sectional design precludes causal inference. The temporal relationship between gut microbial alterations, tryptophan metabolism, and calcific aortic stenosis progression cannot be determined from the present data.

Sixth, renal function was estimated using creatinine-based equations (CKD-EPI), which may underestimate true GFR in elderly patients with reduced muscle mass. Although indole-3-sulfate levels were not significantly correlated with eGFR in either group (see Results), cystatin C-based estimation would be preferable in future studies to improve the accuracy of renal function assessment.

Finally, although Mendelian randomization analyses provided complementary evidence supporting selected pathways, some genetic instruments were limited in strength or availability, and horizontal pleiotropy cannot be completely excluded. In addition, genetic instruments reflect systemic exposures and may not fully capture local biological processes within the aortic valve. Experimental validation was beyond the scope of this study, and mechanistic studies will be required to establish causal links between the gut microbiome, tryptophan metabolism, and valvular calcification.

## CONCLUSION

This exploratory multi-omics study identifies a selective shift of tryptophan metabolism toward the kynurenine pathway in CAS patients — observed in the absence of global gut microbial differences and significant after adjustment for sex, though sex-stratified analyses indicate this elevation is present primarily in male patients. Mendelian randomization analyses positioned kynurenine and indole derivatives as downstream markers of CAS-related inflammatory activation rather than primary drivers, while receptor-level inflammatory signals (IL6Rα, IL18R) and ALPL showed evidence of forward causal associations with CAS risk.

Together, these findings are consistent with a focused gut–host metabolic remodeling in CAS, characterized by preferential inflammatory tryptophan catabolism. They generate a specific, testable hypothesis: that IFN-γ–driven altered kynurenine-indole balance may contribute to kynurenine accumulation in a sex-dependent manner. Confirming this hypothesis will require prospective, adequately powered, multi-cohort studies incorporating functional microbiome characterization and experimental validation.

## DECLARATIONS

### Ethics approval and consent to participate

The study was approved by the local ethics committee (BASEC). All participants provided written informed consent prior to inclusion.

### Clinical Trial number

This study is registered at ClinicalTrials.gov (NCT06021535).

## Supporting information

Main tables

STROBE

## Data Availability

All data produced in the present study are available upon reasonable request to the authors

## Acknowledgement

The authors gratefully acknowledge financial support from the Peter Bockhoff Foundation, the Sana Foundation, and the Swiss Life Foundation for the metabolomics and metagenomics analyses.

The summary Figure was created with BioRender.com

## Data Availability Statement

The datasets generated and/or analyzed during the current study are not publicly available due to patient privacy considerations but are available from the corresponding author on reasonable request.

## Conflicts of interest

David Reineke reports travel expenses from Abbott, Edwards Lifesciences and Medtronic and has proctor and consulting contracts with Abbott and Medtronic.

Caroline Chong-Nguyen reports research grants from the French Society of Cardiology and the MIDHOS group, research funding from the Swiss Life Foundation, the Sana Foundation and the Peter Bockhoff Foundation, travel and educational grants from Abbott as well as consulting and proctoring for Abbott without personal remuneration.

Cyril Ferro reports travel expenses to the institution from Boston Scientific without personal remuneration.

Thomas Pilgrim reports research grants from the Swiss National Science Foundation, the Swiss Heart Foundation, the Swiss Polar Institute, the Bangerter-Rhyner Foundation, the Mach-Gaensslen Foundation, and the Monsol Foundation. Research, travel or educational grants to the institution without personal remuneration from Biotronik, Boston Scientific, Edwards Lifesciences, and ATSens; speaker fees and consultancy fees to the institution from Biotronik, Boston Scientific, Edwards Lifesciences, Abbott, Medtronic, Biosensors, and Highlife.

All other co authors declared no conflict of interest for this manuscript

## Author contributions

Caroline Chong-Nguyen conceptualized and designed the study, supervised the project, performed data analysis and interpretation, and drafted the manuscript.

Sarah Atighetchi contributed to patient recruitment and clinical data collection.

Cyril Ferro contributed to patient recruitment, clinical data collection, and manuscript revision. Bahtiyar Yilmaz contributed to data interpretation, particularly in microbiome analyses.

Harry Sokol contributed to the interpretation of metabolomics and integrated analysis of the results. Matthias Siepe contributed to surgical procedures and aortic valve tissue collection

David Reineke contributed to surgical procedures and aortic valve tissue collection Selim Mosbahi contributed to surgical procedures and aortic valve tissue collection. Daijiro Tomii contributed to biological sample collection and biobanking

Masaaki Nakase contributed to biological sample collection and biobanking. Christoph Wingert contributed to patient recruitment and clinical data collection. Liam Tanner contributed to data collection and project support.

Camille Dupuy developed and performed metabolomics analyses.

Lydie Nadal-Desbarats developed and performed metabolomics analyses.

Yara Banz performed tissue processing and preparation for storage and downstream analyses.

Tereza Losmanovà performed tissue processing and preparation for storage and downstream analyses.

Pamela Nicholson performed metagenomics experiments.

Aparna Pandey performed statistical and bioinformatic analyses, particularly for metabolomics data.

Yvonne Döring contributed to study design, critically revised the manuscript, and provided senior supervision.

Thomas Pilgrim contributed to study design, critically revised the manuscript, and provided senior supervision.

All authors critically revised the manuscript and approved the final version.

## ABBREVIATIONS

AhR: Aryl hydrocarbon receptor
BA / BAs: Bile acid / bile acids
BMI: Body mass index
CA: Cholic acid
CAS: Calcific aortic stenosis
CDCA: Chenodeoxycholic acid
CT: Computed tomography
CVD: Cardiovascular disease
DNA: Deoxyribonucleic acid
HbA1c: Hemoglobin A1c
HDL: High-density lipoprotein
IDO: Indoleamine 2,3-dioxygenase
IS: Indoxyl sulfate
LDL: Low-density lipoprotein
LV: Left ventricle
NMDA: N-methyl-D-aspartate
NOTCH1: Notch receptor 1
PERK: Protein kinase R-like endoplasmic reticulum kinase
RAAS: Renin–angiotensin–aldosterone system
Runx2: Runt-related transcription factor 2
SCFA / SCFAs: Short-chain fatty acid / short-chain fatty acids
SSRI / SSRIs: Selective serotonin reuptake inhibitor(s)
TAVI: Transcatheter aortic valve implantation
THCA: Taurocholic acid
TMA: Trimethylamine
TMAO: Trimethylamine N-oxide
TRP: Tryptophan
UDCA: Ursodeoxycholic acid
VECs: Valvular endothelial cells
VICs: Valvular interstitial cells
Wnt: Wnt signaling pathway
XBP-1: X-box binding protein 1

---

**Summary Figure.**
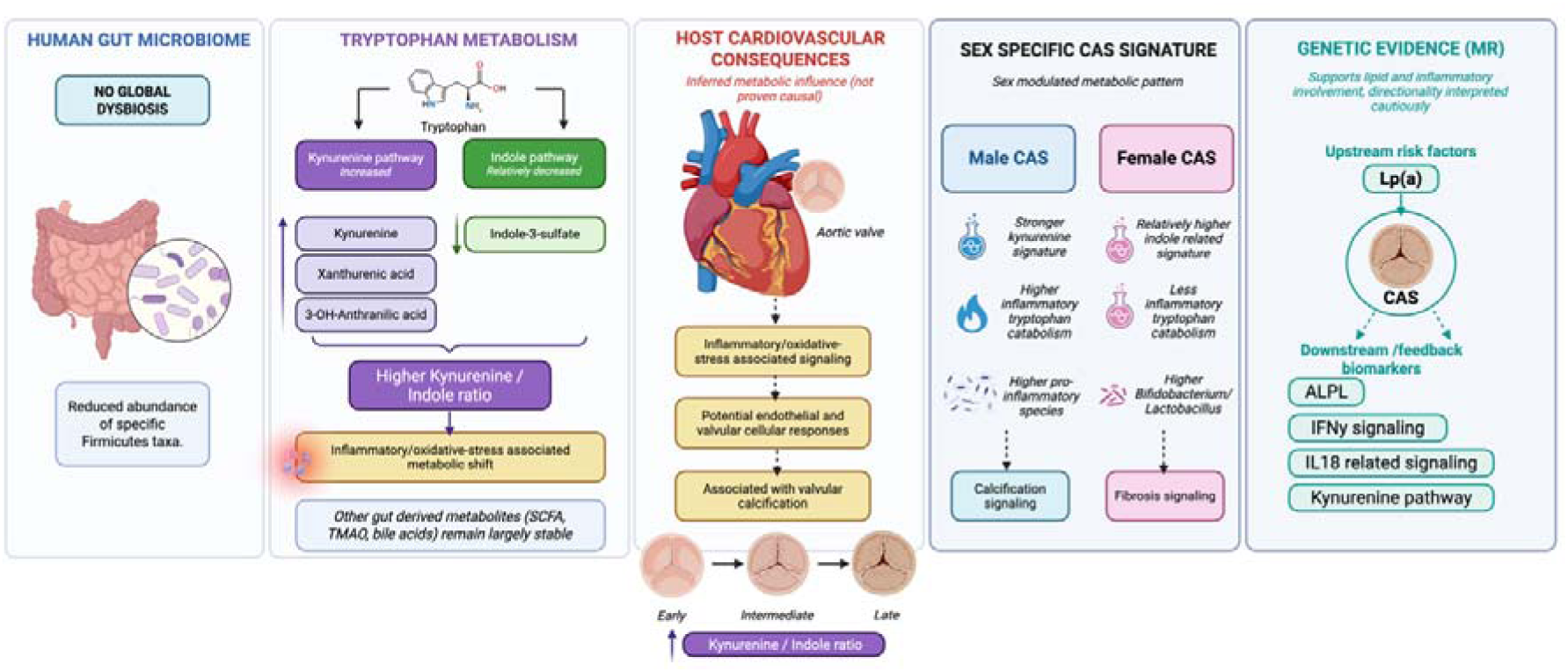
Proposed gut–host metabolic axis in calcific aortic stenosis (CAS). In this study, CAS was associated with a selective gut–host metabolic signature rather than major global disruption of the gut microbiome. (1) **Human gut microbiome**: CAS was associated with depletion of selected *Firmicutes/Bacillota* taxa, including *Eubacterium coprostanoligenes, Intestinibacter, Clostridium sensu stricto 1, and Christensenellaceae R-7* group, together with reduced cholesterol-to-coprostanol conversion, while global alpha and beta diversity were largely preserved. (2) **Tryptophan metabolism**: targeted metabolomics showed a relative shift toward the kynurenine pathway, with higher kynurenine, 3-hydroxyanthranilic acid, and xanthurenic acid, lower indole-3-sulfate, and a higher kynurenine/indole ratio in CAS. In contrast, short-chain fatty acids, bile acids, and TMAO-related metabolites were largely unchanged overall. (3) **Proposed host cardiovascular consequences**: these findings support an inferred metabolic model linking altered gut-derived metabolite balance to inflammation/oxidative-stress-associated signaling, potential endothelial and valvular cellular responses, and association with valvular calcification; notably, a higher kynurenine/indole ratio was progressively associated with increased severity of the disease (from early to intermediate and late stages). These links are biologically plausible but were not directly proven as causal in the cohort. (4) **Sex-specific CAS signatures**: male CAS patients showed a stronger kynurenine signature, higher inflammatory tryptophan catabolism, and a higher abundance of pro-inflammatory species linked to calcification signaling; whereas female CAS patients showed relatively higher indole-related signatures, less inflammatory tryptophan catabolism, higher *Bifidobacterium/Lactobacillus*, and an association with fibrosis signaling. (5) **Genetic evidence (Mendelian randomization)**: genetic analyses supported involvement of lipid and inflammatory pathways in CAS, with lipoprotein(a) [Lp(a)] represented as the upstream risk factor, and ALPL, IFNγ-related signaling, IL18-related signaling, and kynurenine-pathway alterations represented as downstream or feedback biomarkers.

