## Supplementary Tables and Figures for "Altered kynurenine and indole tryptophan pathways in calcific aortic stenosis: cross-sectional evidence for a gut-host metabolic imbalance (GUT-CAS)"

**Supplementary Material**

### Table of Contents

**Supplementary Figures**

Figure 1. Robustness of kynurenine-pathway metabolite signals to clinical confounder adjustment

Figure 2. Indole-3-sulfate concentrations and eGFR in CAS patients

Figure 3. Leave-one-out sensitivity analysis: KI ratio vs. indexed AVA

Figure 4. Cumulative pathway scores of tryptophan metabolites in CAS and Non-CAS patients

Figure 5. Single-covariate sensitivity analysis: KI ratio vs. indexed AVA

Figure 6. Sex-stratified gut microbiome diversity and taxonomic associations in CAS

Figure 7. Spearman correlations between the kynurenine-to-indole-3-sulfate (KI) ratio and circulating lipid parameters, stratified by group.

**Supplementary Tables**

Table 1. MRM transitions for all quantified metabolite analytes Table 1A. Tryptophan derivatives metabolomics panel — quantification values

Table 2B. Short-chain fatty acids (SCFA) panel — quantification values

Table 2C. Bile acids (BA) panel — quantification values

Table 2D. TMA/TMAO panel — quantification values

Table 3. Comorbidity-adjusted sensitivity analysis for kynurenine-pathway metabolites

Table 4. Sensitivity analyses: eGFR adjustment (I3S) and platelet adjustment (serotonin)

Table 5. KI Ratio vs. Indexed AVA: Single-Covariate Sensitivity Analysis

Table 6. BH-FDR corrected tryptophan metabolite comparisons by CAS severity

Table 7. Clinical characteristics by CAS severity (Severe vs. Mild-Moderate)

Table 8. Clinical characteristics by sex (Male CAS vs. Female CAS)

Table 9. BH-FDR corrected tryptophan metabolite comparisons by sex

Table 10. Forward MR Lipoprotein(a) Wald Ratio: Confirmed Values

Table 11. Weighted median MR: biomarkers on CAS

Table 12. Weighted median MR: CAS on biomarkers

Table 13. Sensitivity analyses: direct MR

Table 14. Sensitivity analyses: reverse MR

Table 15. Cochran’s Q heterogeneity: direct MR

Table 16. Cochran’s Q heterogeneity: reverse MR

Table 17. Fixed- vs. random-effects IVW: direct MR

Table 18. Fixed- vs. random-effects IVW: reverse MR

Table 19. Leave-one-out sensitivity: direct MR

Table 20. Leave-one-out sensitivity: reverse MR

Table 21. Steiger directionality test: direct MR

Table 22. Steiger directionality test: reverse MR

### Supplementary Figure 1

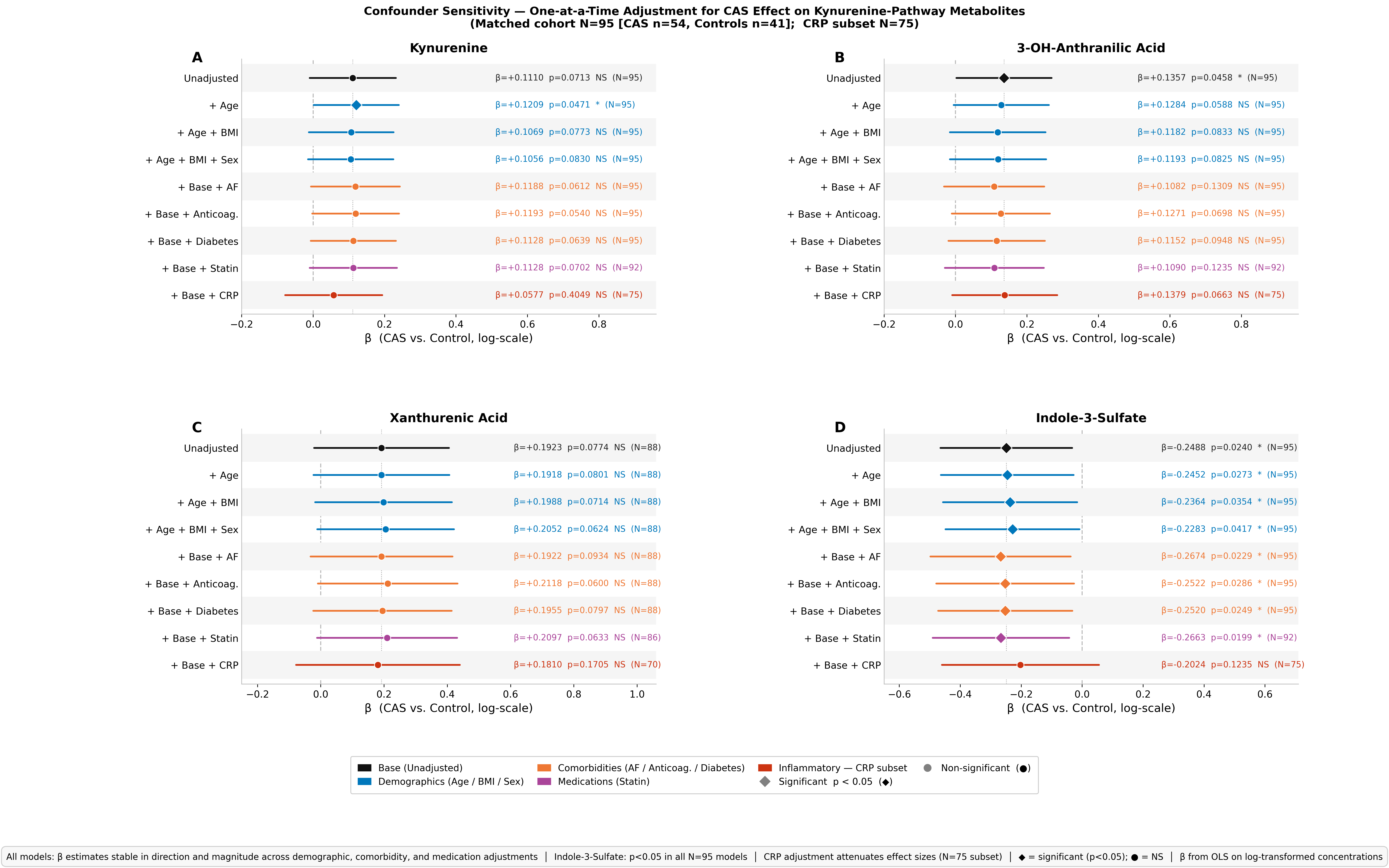

Supplementary Figure 1. Robustness of kynurenine-pathway metabolite signals to adjustment for clinical confounders. Forest plot showing standardized CAS effect sizes (β; log-transformed metabolite concentrations) for key kynurenine-pathway metabolites before and after sequential adjustment for age, BMI, atrial fibrillation, and anticoagulant use. Effect sizes remain directionally consistent and nominally significant across all confounder-adjusted models, supporting the independence of the tryptophan–kynurenine signal from common cardiovascular comorbidities. Linear regression, β per unit CAS status.

### Supplementary Figure 2

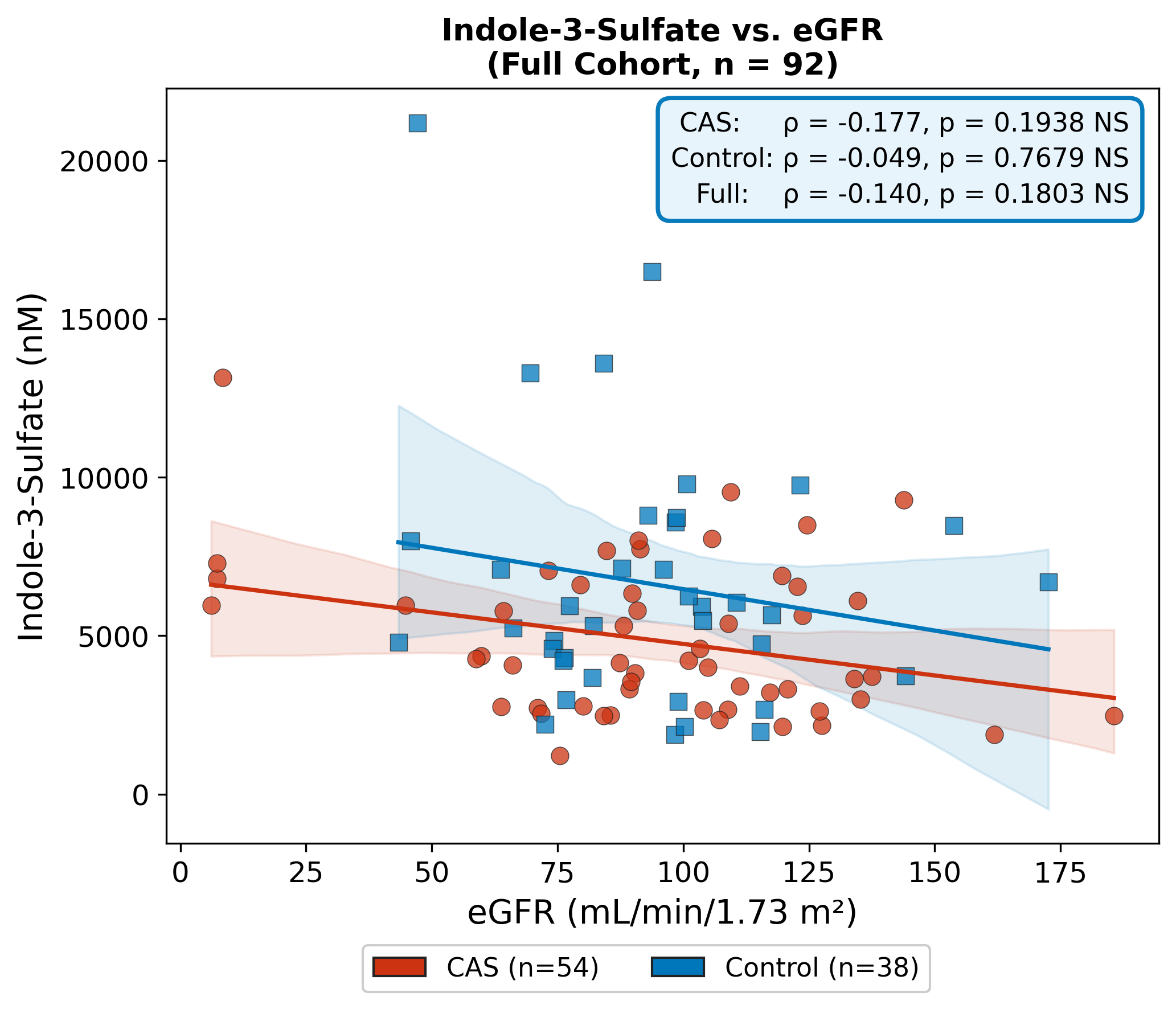

Supplementary Figure 2. Indole-3-sulfate concentrations and estimated glomerular filtration rate (eGFR) (Sensitivity Analysis 5). Scatter plot of indole-3-sulfate concentration vs. eGFR (mL/min/1.73 m²) in the full cohort (n = 92), shown separately for CAS (n = 54) and Non-CAS control (n = 38) patients. No significant correlation was observed in CAS patients (Spearman ρ = −0.18, p = 0.19), in controls (ρ = −0.05, p = 0.77), or in the full cohort (ρ = −0.14, p = 0.18). The absence of a significant correlation indicates that lower indole-3-sulfate concentrations observed in CAS are not attributable to renal clearance confounding, supporting the gut microbial origin of the indole-pathway depletion. Spearman ρ; two-tailed.

### Supplementary Figure 3

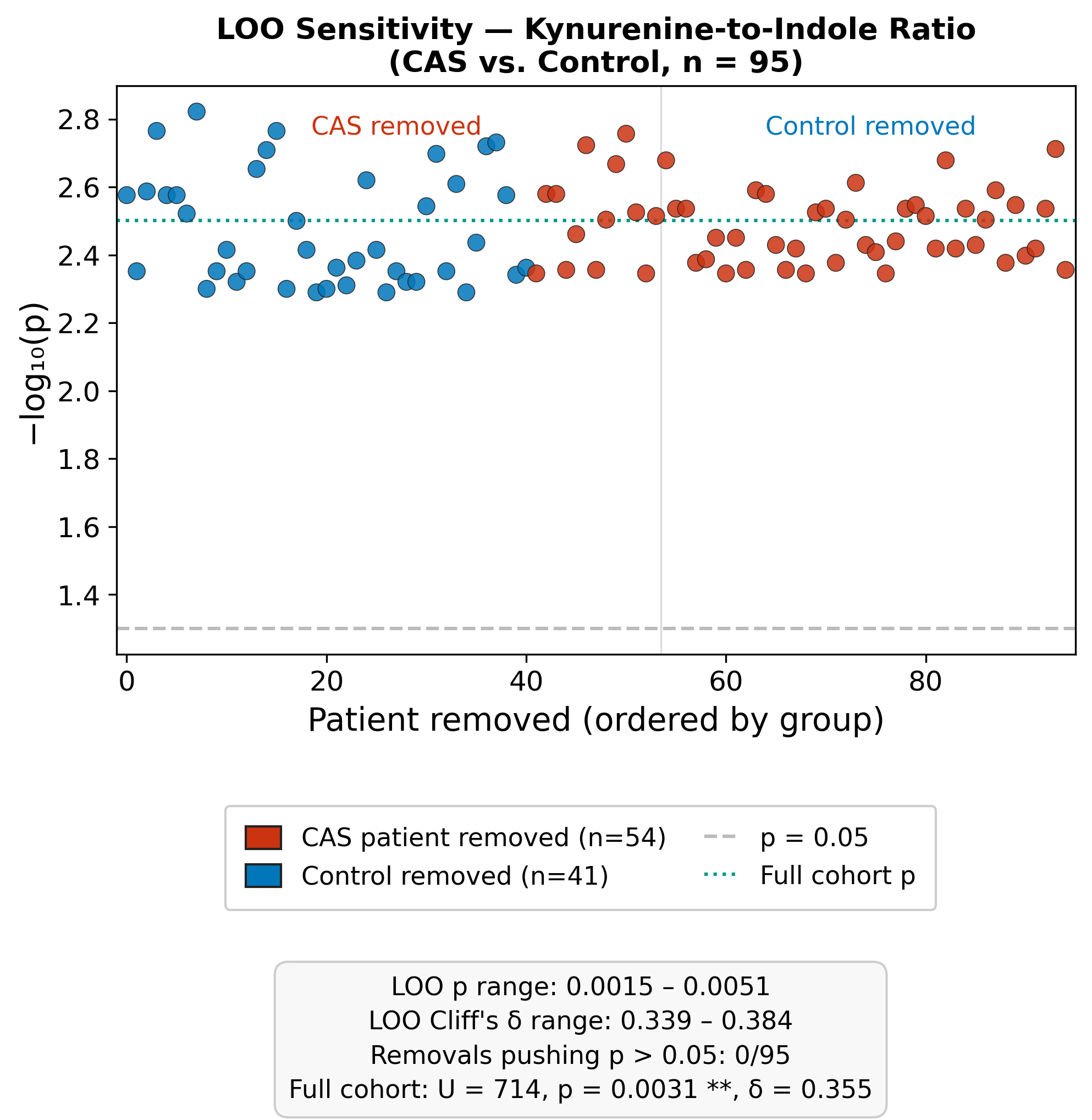

Supplementary Figure 3. Leave-one-out sensitivity analysis for the kynurenine-to-indole ratio difference between CAS and Non-CAS patients. Sequential leave-one-out analysis removing one participant at a time from the Mann-Whitney comparison of the log₂ kynurenine-to-indole-3-sulfate (KI) ratio between CAS (n = 54) and Non-CAS control (n = 41) patients (n = 95 total). Each point represents the −log₁₀(p) of the group comparison with the corresponding participant excluded; the dashed line marks p = 0.05 and the dotted line the full-cohort p-value. Significance was maintained across all iterations (leave-one-out p range 0.0015–0.0051; 0 of 95 removals raised p above 0.05; full cohort Mann-Whitney U = 714, p = 0.0031, Cliff’s δ = 0.355), confirming that no single influential observation drives the elevated KI ratio in CAS.

### Supplementary Figure 4

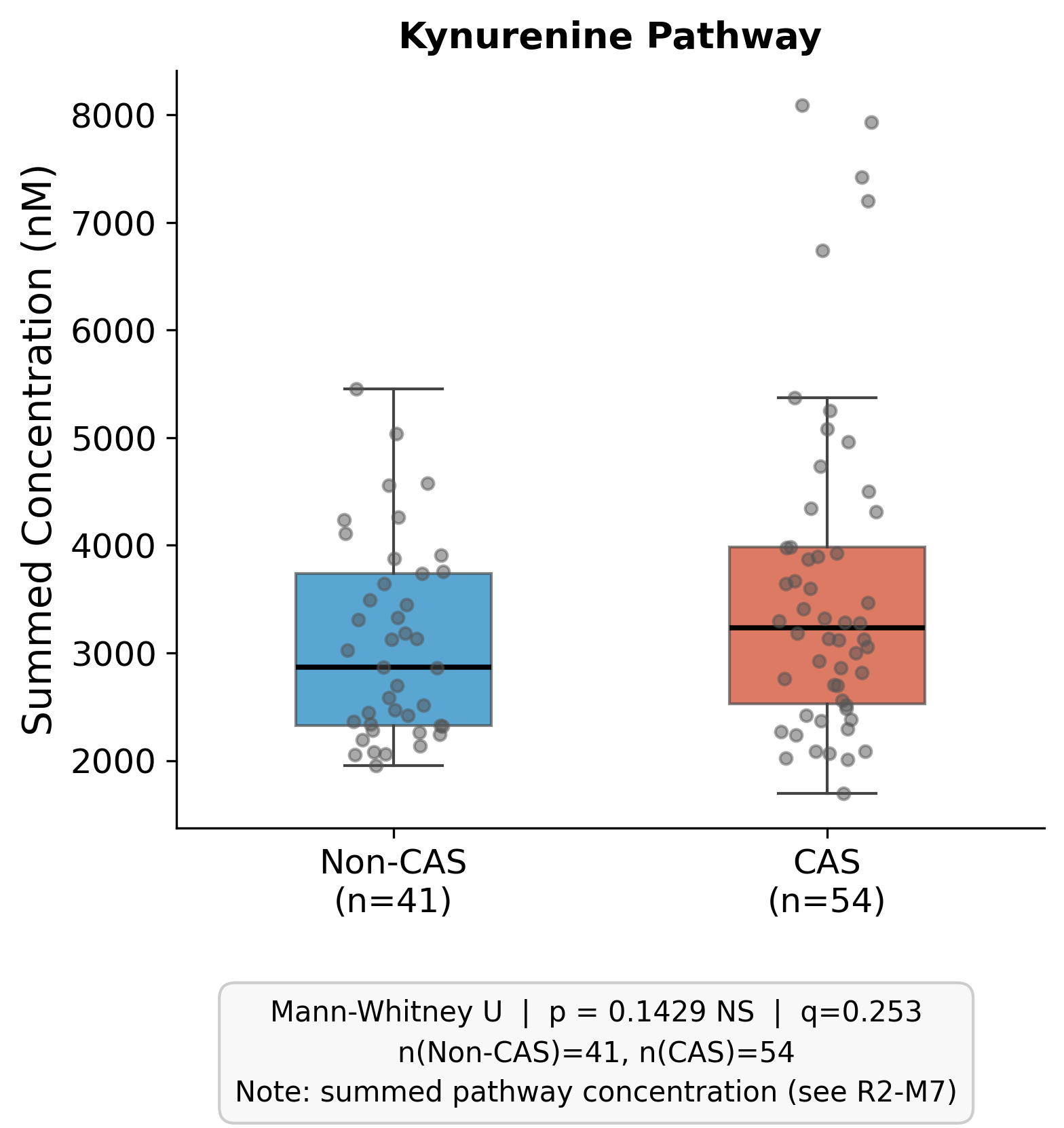

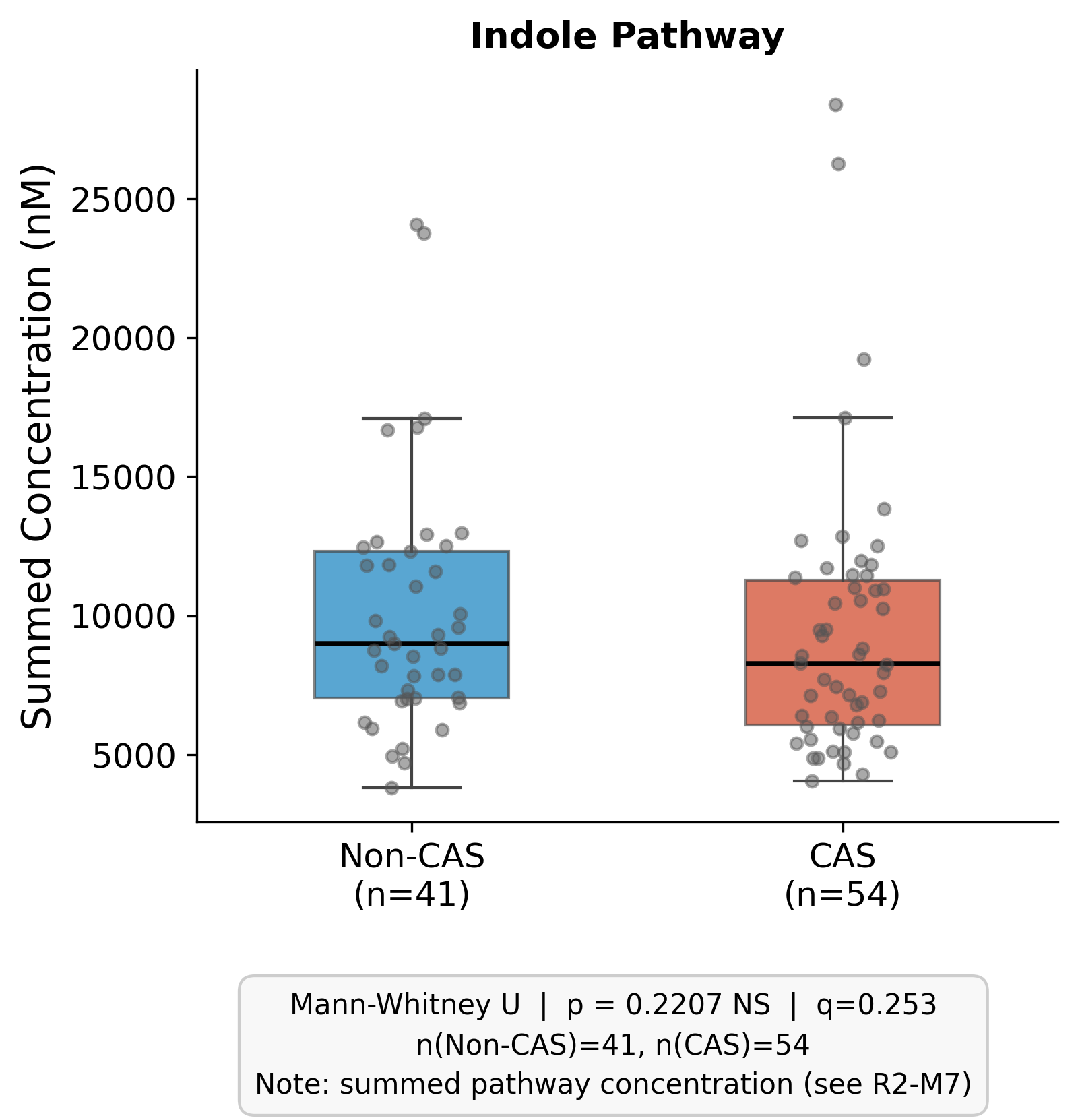

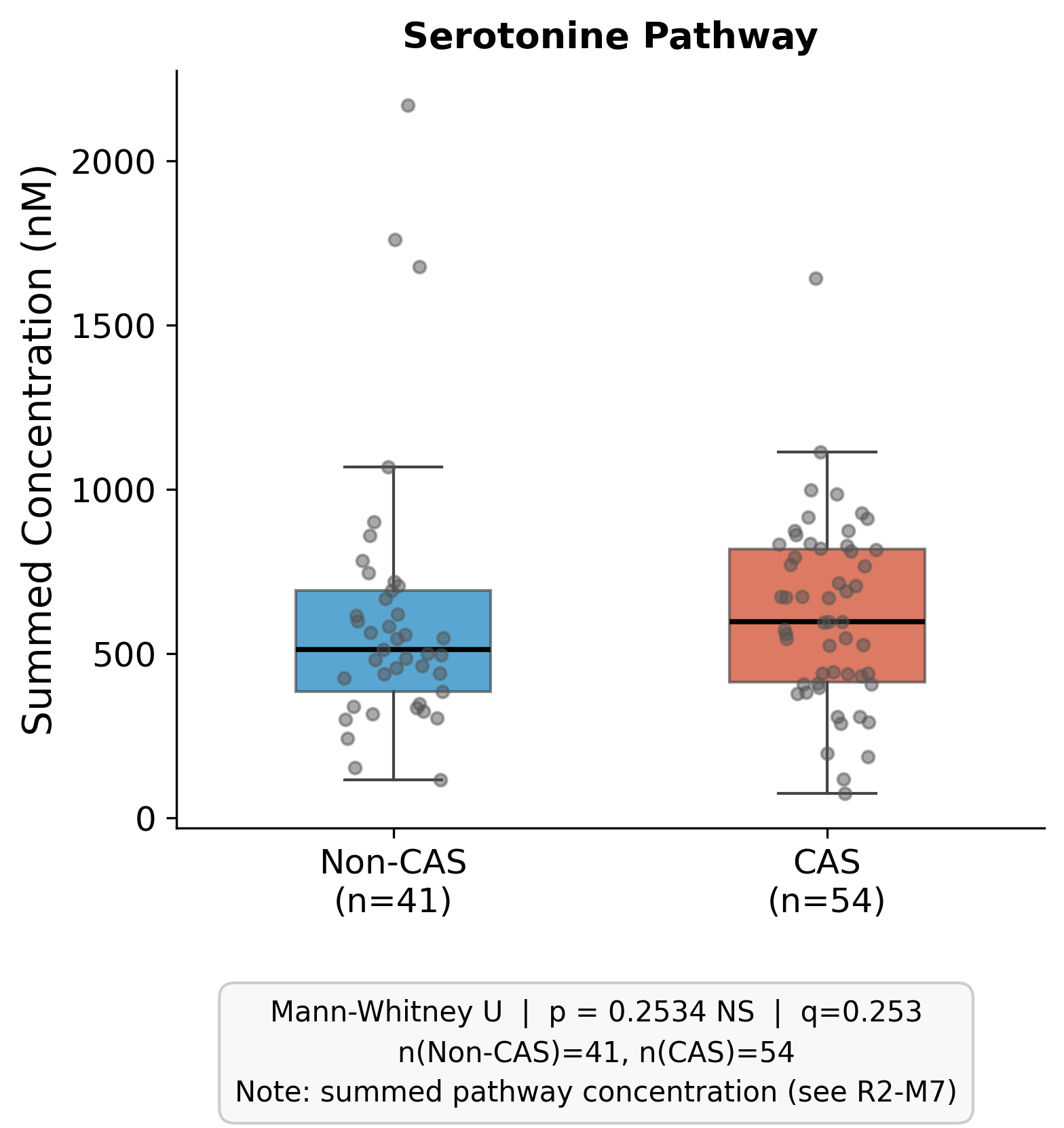

Supplementary Figure 4. Cumulative pathway scores of tryptophan metabolites in CAS and Non-CAS patients. Box plots show the summed concentrations of metabolites within the kynurenine pathway (A), indole pathway (B), and serotonin pathway (C) in matched CAS (n = 54) and Non-CAS (n = 41) patients. None of the aggregate pathway scores differed significantly between groups (kynurenine p = 0.14, indole p = 0.22, serotonin p = 0.25; all q = 0.25). The absence of a difference at the cumulative pathway level indicates that the CAS-associated tryptophan signal is driven by specific individual metabolites rather than by a uniform shift across an entire pathway. Mann-Whitney U test with BH-FDR correction.

### Supplementary Figure 5

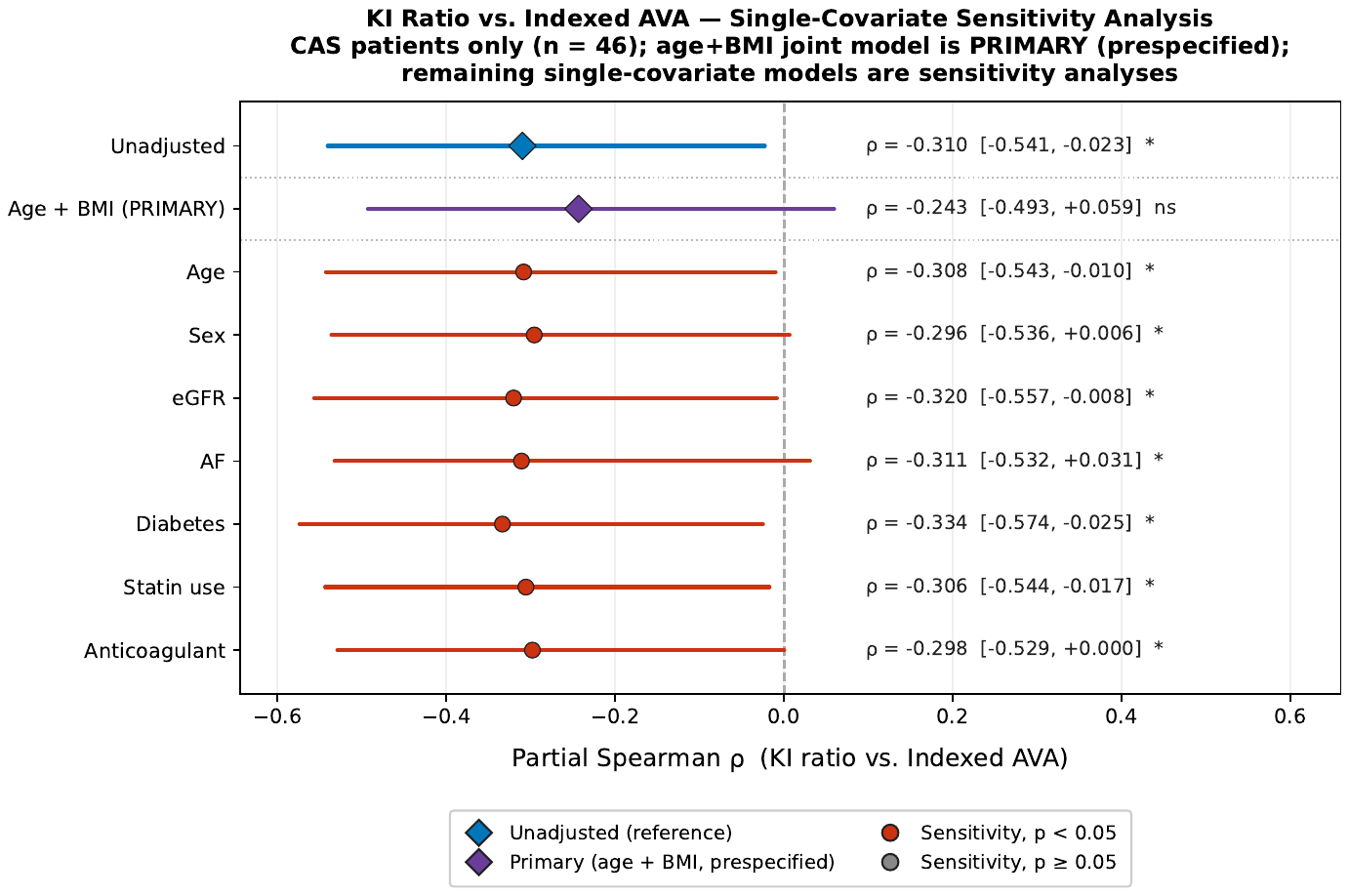

Supplementary Figure 5. Single-covariate sensitivity analysis for the kynurenine-to-indole ratio vs. indexed aortic valve area. Forest plot of partial Spearman ρ between log₂ kynurenine-to-indole-3-sulfate ratio and indexed AVA (cm²/m²), with sequential adjustment for individual clinical covariates (age, BMI, sex, renal function, medications). The inverse association remains stable across all single-covariate adjustments, supporting robustness of the KI ratio–AVA correlation except for BMI+age. n = 46 CAS patients with available echocardiographic data.

### Supplementary Figure 6

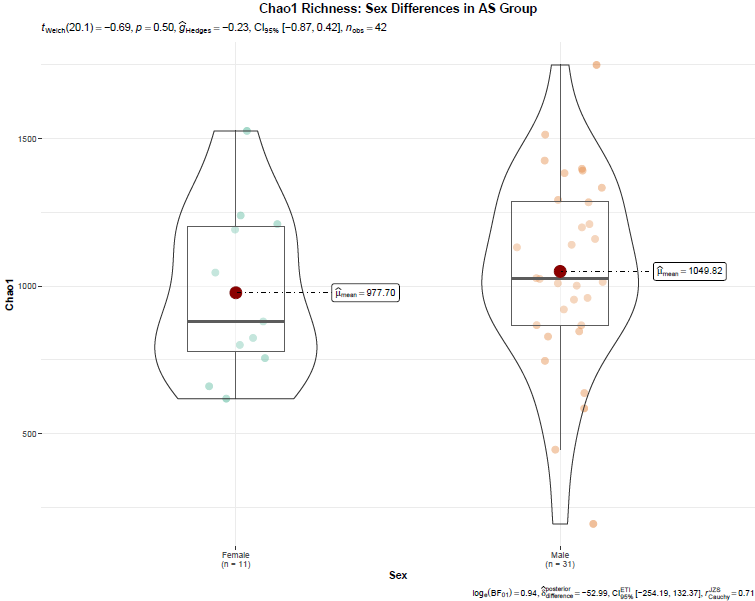

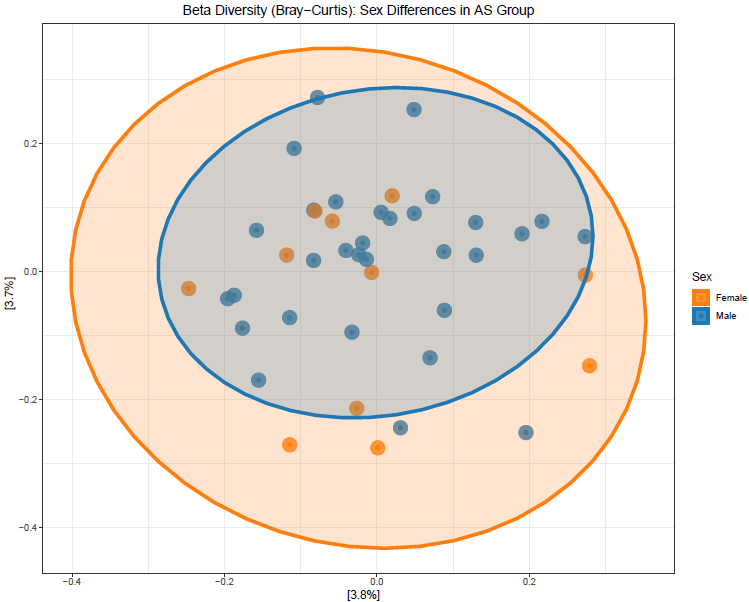

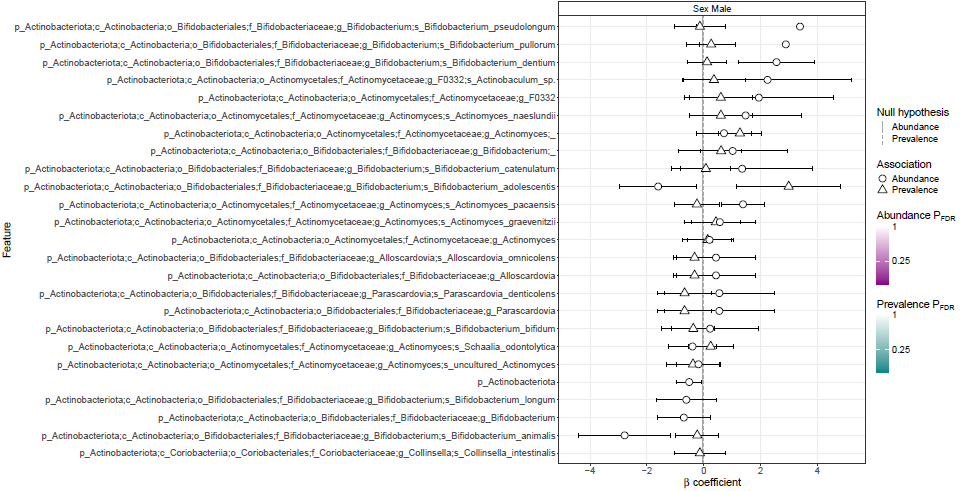

Supplementary Figure 6. Sex-stratified gut microbiome diversity and taxonomic associations in CAS. Shannon alpha diversity and Chao1 species richness did not differ significantly by sex within the CAS group (Mann-Whitney U, p > 0.05 for both), indicating comparable within-sample diversity in male and female CAS patients. Bray-Curtis beta-diversity ordination showed no marked separation by sex. Multivariable differential abundance analysis identified several genera nominally associated with sex within the CAS group, consistent with known physiological sex differences in gut microbiome composition.

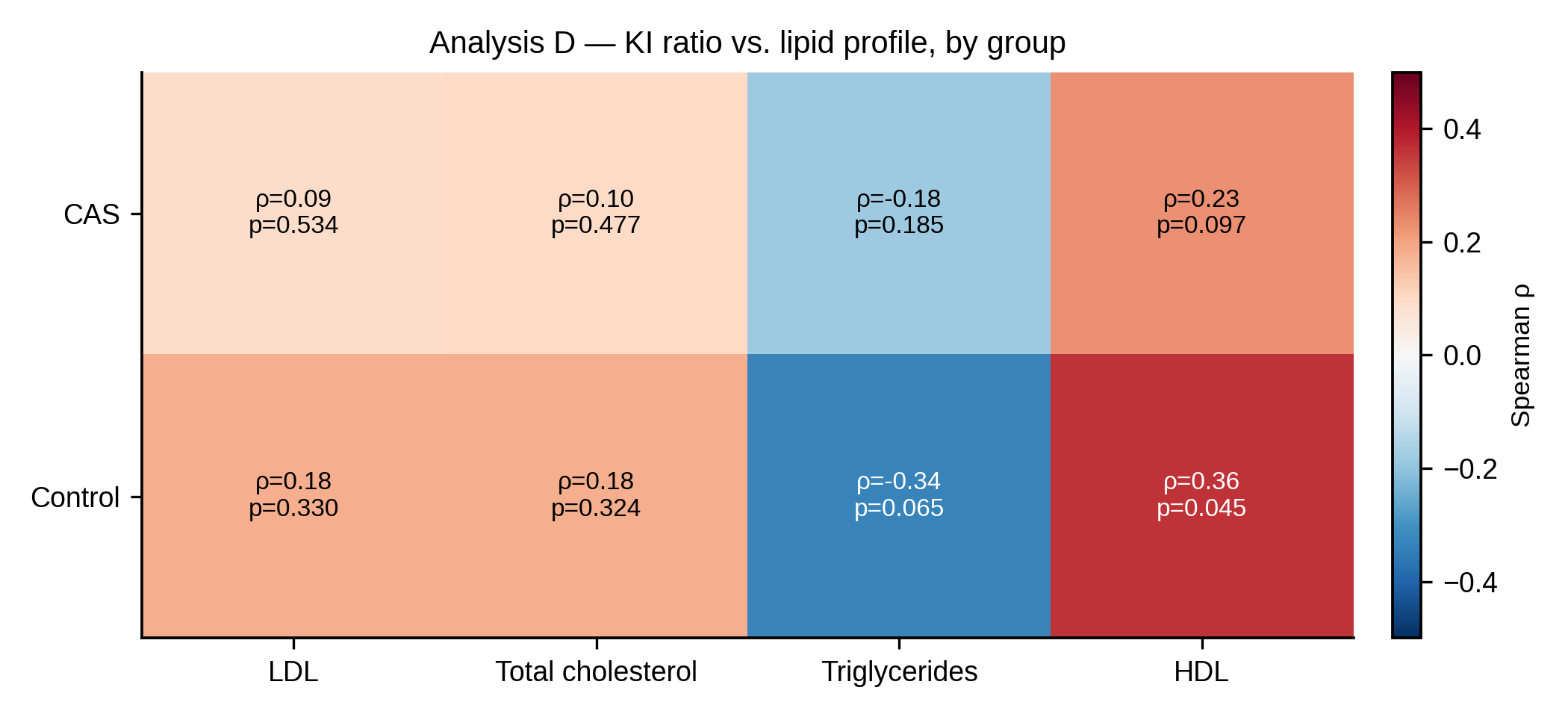

**Supplementary Figure 7. Spearman correlations between the kynurenine-to-indole-3-sulfate (KI) ratio and circulating lipid parameters, stratified by group.**

Heatmap showing Spearman rank correlation coefficients (ρ) between the KI ratio and four fasting lipid parameters (LDL cholesterol, total cholesterol, triglycerides, and HDL cholesterol) in CAS patients (upper row; n = 50–54 depending on lipid data availability) and controls (lower row; n = 31). Cell colour reflects the magnitude and direction of ρ (red = positive, blue = negative); Spearman ρ and unadjusted p-values are annotated within each cell. No significant associations were observed between the KI ratio and any lipid parameter in CAS patients (all |ρ| ≤ 0.23, all p ≥ 0.097), indicating that the KI ratio elevation in this group is not driven by dyslipidaemia. In controls, a nominal positive correlation was observed between the KI ratio and HDL cholesterol (ρ = 0.36, p = 0.045); this association did not reach significance after correction for multiple comparisons and was not present in CAS patients, and therefore does not confound the primary group-level finding. No corrections for multiple comparisons were applied within this sensitivity analysis; all p-values are unadjusted.

### Supplementary Table 1. MRM Transitions for All Quantified Metabolite Analytes

Multiple Reaction Monitoring (MRM) transitions used for targeted LC-MS/MS quantification of metabolites across all four analytical panels (serum). Columns: Name = analyte name; Trace = precursor > product ion (m/z); RT = retention time (min); ES = electrospray ionization mode (POS/NEG); CV = cone voltage (V); CE = collision energy (eV). This table lists technical LC-MS/MS acquisition parameters; it contains no P values and no statistical tests were performed.

#### TRP Derivatives (Serum)

| **Name** | **Trace** | **RT** | **ES** | **CV** | **CE** |
| --- | --- | --- | --- | --- | --- |
| **Picolinic acid** | 124.1 > 78.103 | 0.7 | POS | 20 | 16 |
| **Quinolinic acid** | 166.1 > 78.03 | 0.87 | NEG | 10 | 11 |
| **3-OH-Kynurenine** | 225.2 > 110.1010 | 0.79 | POS | 5 | 13 |
| **Serotonine** | 177.1 > 160.202 | 0.95 | POS | 10 | 12 |
| **5-OH-Tryptophane** | 221.1 > 162.104 | 1.11 | POS | 10 | 16 |
| **Kynurenine** | 209.1 > 94.104 | 1.29 | POS | 10 | 30 |
| **Tryptamine** | 161.2 > 144.104 | 2.06 | POS | 30 | 14 |
| **3-OH-Anthranilic acid** | 154.2 > 136.104 | 1.65 | POS | 10 | 15 |
| **Tryptophane** | 205.1 > 188.104 | 2.22 | POS | 5 | 20 |
| **5-OH-Indole acetic acid** | 192.2 > 146.005 | 2.65 | POS | 10 | 22 |
| **Indole-3-Sulfate** | 211.9 > 80.001 | 2.99 | NEG | 10 | 12 |
| **N-acetyl-serotonine** | 219.2 > 160.201 | 3.18 | POS | 5 | 12 |
| **Xanthurenic acid** | 206.1 > 160.104 | 3.48 | POS | 10 | 22 |
| **Indole-3-acetamide** | 175.1 > 130.103 | 3.76 | POS | 19 | 15 |
| **Kynurenic acid** | 190.1 > 144.104 | 3.8 | POS | 10 | 16 |
| **Indole-3-Lactic acid** | 204 > 186.105 | 4.6 | NEG | 5 | 14 |
| **Indole-3-Aldehyde** | 146.1 > 118.104 | 4.7 | POS | 38 | 17 |
| **Indole-3-Acetic acid** | 176.1 > 130.101 | 5.01 | POS | 10 | 30 |
| **Tryptophol** | 162.2 > 144.205 | 5.05 | POS | **0** | 16 |
| **Melatonine** | 233.2 > 174.102 | 5.18 | POS | 38 | 10 |
| **Indole-3-propionic acid** | 190.1 > 130.105 | 5.88 | POS | 20 | 30 |
| **PICO_d4** | 128.1 > 82.1 | 0.7 | POS | 20 | 16 |
| **QUIN-d3** | 169 > 81 | 0.86 | NEG | 10 | 11 |
| **3-OH-K-13C2-15N** | 228.1 > 110.1 | 0.79 | POS | 5 | 13 |
| **Serotonine-d4** | 181.1 > 164 | 0.94 | POS | 10 | 12 |
| **5-OH-TRP-d4** | 225.1 > 165.1 | 1.11 | POS | 10 | 16 |
| **KYN-d4** | 213.1 > 98 | 1.27 | POS | 10 | 30 |
| **3-OH-AA-d3** | 157.2 > 139.1 | 1.62 | POS | 10 | 15 |
| **Tryptamine-d4** | 165 > 148 | 2.04 | POS | 30 | 14 |
| **TRP-d5** | 210.1 > 193.1 | 2.18 | POS | 5 | 20 |
| **5-HIAA-d5** | 197 > 150 | 2.61 | POS | 10 | 22 |
| **I-3-S 13C6** | 218 > 80 | 3 | NEG | 10 | 12 |
| **N-ac-SER-d3** | 222.2 > 160.2 | 3.17 | POS | 5 | 12 |
| **XANA-d4** | 210 > 164.1 | 3.45 | POS | 10 | 22 |
| **I-3-acNH2-d5** | 180 > 134 | 3.69 | POS | 19 | 15 |
| **KYNA-d5** | 195 > 149 | 3.76 | POS | 10 | 16 |
| **I-3-LA-d5** | 209 > 191.1 | 4.55 | NEG | 5 | 14 |
| **I-3-ALD-13C8** | 154.1 > 126 | 4.7 | POS | 38 | 17 |
| **I-3-AA-d4** | 180 > 133 | 4.97 | POS | 10 | 30 |
| **Tryptophol-d4** | 166.2 > 148.2 | 5 | POS | **0** | 16 |
| **I-3-PA d2** | 192.1 > 130.104 | 5.86 | POS | 20 | 30 |
| **MEL-d4** | 237 > 178 | 5.16 | POS | 38 | 10 |

#### Short-Chain Fatty Acids (SCFA)

| **Name** | **Trace** | **RT** | **ES** | **CV** | **CE** |
| --- | --- | --- | --- | --- | --- |
| **Acetic acid** | 194.1 > 152. | 1.18 | NEG | 20 | 14 |
| **Propionic acid** | 208.1 > 165.1 | 1.74 | NEG | 20 | 16 |
| **Butyric acid** | 222.1 > 179.1 | 2.62 | NEG | 20 | 14 |
| **Isobutyric acid** | 222.1 > 179.1 | 2.49 | NEG | 20 | 14 |
| **Valeric acid** | 236.1 > 193.1 | 3.75 | NEG | 20 | 14 |
| **Isovaleric acid** | 236.1 > 193.1 | 3.6 | NEG | 20 | 14 |
| **C2_EI_13C2** | 196.1 > 152.001 | 1.18 | NEG | 20 | 14 |
| **C4_EI_d7** | 229.1 > 186.1 | 2.57 | NEG | 20 | 14 |
| **C5_EI_d9** | 245.1 > 202.1 | 3.7 | NEG | 20 | 14 |
| **C3_EI_d6** | 214.1 > 171.1 | 2.41 | NEG | 20 | 16 |

#### Bile Acids

| **Name** | **Trace** | **RT** | **ES** | **CV** | **CE** |
| --- | --- | --- | --- | --- | --- |
| **LCA** | 375.301 > 375.301 | 10.82 |  | 60 | 10 |
| **LCA d4** | 379.301 > 379.301 | 10.82 |  | 60 | 10 |
| **UDCA** | 391.301 > 391.301 | 7.96 |  | 60 | 10 |
| **CDCA** | 391.301 > 391.301 | 10.21 |  | 60 | 10 |
| **DCA** | 391.301 > 391.301 | 10.34 |  | 60 | 10 |
| **UDCA d4** | 395.301 > 395.301 | 7.95 |  | 60 | 10 |
| **CDCA d4** | 395.301 > 395.301 | 10.21 |  | 60 | 10 |
| **DCA d5** | 396.301 > 396.301 | 10.33 |  | 60 | 10 |
| **HCA** | 407.301 > 407.301 | 7.66 |  | 60 | 10 |
| **CA** | 407.301 > 407.301 | 8.23 |  | 60 | 10 |
| **CA d5** | 412.305 > 412.305 | 8.21 |  | 60 | 10 |
| **GLCA** | 432.3 > 74. | 10.31 |  | 60 | 40 |
| **GUDCA** | 448.3 > 74. | 5.89 |  | 60 | 40 |
| **GCDCA** | 448.3 > 74. | 8.45 |  | 60 | 40 |
| **GDCA** | 448.3 > 74. | 8.96 |  | 60 | 40 |
| **LCA-S** | 455.247 > 97 | 9.89 |  | 60 | 40 |
| **GHCA** | 464.3 > 74. | 5.55 |  | 60 | 40 |
| **GCA** | 464.3 > 74. | 6.43 |  | 60 | 40 |
| **GCA d4** | 468.326 > 74. | 6.43 |  | 60 | 40 |
| **TLCA** | 482.3 > 80. | 9.39 |  | 60 | 40 |
| **CA-S** | 487.237 > 97 | 6.04 |  | 60 | 40 |
| **CA-S d4** | 491.262 > 97 | 6.05 |  | 60 | 40 |
| **TUDCA** | 498.28 > 80. | 4.76 |  | 60 | 60 |
| **TCDCA** | 498.28 > 80. | 7.16 |  | 60 | 60 |
| **TDCA** | 498.28 > 80. | 7.64 |  | 60 | 60 |
| **THDCA d4** | 502.28 > 84. | 4.76 |  | 60 | 60 |
| **THCA** | 514.28 > 80. | 4.37 |  | 60 | 60 |
| **TCA** | 514.28 > 80. | 5.3 |  | 60 | 60 |
| **TCA d4** | 518.309 > 80. | 5.29 |  | 60 | 60 |

#### TMA/TMAO

| **Name** | **Trace** | **RT** | **ES** | **CV** | **CE** |
| --- | --- | --- | --- | --- | --- |
| **TMA** | 146.1 > 59.102 | 2.64 | POS | 10 | 25 |
| **Choline** | 104.1 > 60.03 | 4.05 | POS | 10 | 25 |
| **TMAO** | 76 > 59 | 3.51 | POS | 10 | 20 |
| **L-Carnitine** | 162.1 > 85.03 | 5.04 | POS | 10 | 30 |
| **TMA-d9** | 155.1 > 68 | 2.64 | POS | 10 | 20 |
| **Choline-d9** | 113.1 > 69 | 4.05 | POS | 10 | 25 |
| **TMAO-d9** | 85 > 68 | 3.47 | POS | 10 | 20 |
| **L-Carnitine-d9** | 171.1 > 85.03 | 5.04 | POS | 10 | 30 |

**Supplementary Table 2A. Tryptophan Derivatives Metabolomics Panel**

Absolute concentrations of tryptophan pathway metabolites quantified by stable-isotope dilution LC-MS/MS in the matched cohort. Values < LLOQ are reported as not detected (ND). Concentrations in nmol/L for serum samples. This table reports absolute measured concentrations; it contains no P values and no statistical tests were performed.

| **Metabolite** | **Non-CAS (median [IQR])** | **CAS (median [IQR])** | **p value** | **FDR q value** |
| --- | --- | --- | --- | --- |
| **Picolinic acid** | 16.41 [13.96–27.95] | 20.03 [14.13–32.87] | 0.825 | 0.868 |
| **3-OH-Kynurenine** | 47.31 [35.60–61.33] | 48.72 [38.40–61.32] | 0.342 | 0.534 |
| **Quinolinic acid** | 630.29 [455.19–877.89] | 638.73 [435.01–1070.43] | 0.666 | 0.783 |
| **Serotonin** | 438.53 [343.82–597.28] | 537.38 [358.61–729.27] | 0.182 | 0.456 |
| **5-OH-Tryptophan** | 5.69 [4.80–7.76] | 5.40 [4.63–6.73] | 0.367 | 0.534 |
| **Kynurenine** | 1986.76 [1699.15–2744.73] | 2300.85 [1911.46–2869.23] | ***0.078*** | ***0.254*** |
| **Tryptamine** | 1.27 [1.16–1.86] | 1.23 [1.13–1.45] | 0.374 | 0.534 |
| **3-OH-Anthranilic acid** | 42.58 [33.55–49.46] | 50.05 [41.50–57.61] | **0.030** | **0.227** |
| **Tryptophan** | 63785.12 [55587.52–71493.01] | 64568.20 [56866.29–76868.15] | 0.596 | 0.745 |
| **5-OH-Indole acetic acid** | 64.59 [51.70–86.54] | 62.76 [48.57–74.39] | 0.361 | 0.534 |
| **Indole-3-Sulfate** | 5652.37 [4062.48–7991.94] | 4178.70 [2772.39–6496.20] | **0.040** | **0.227** |
| **Xanthurenic acid** | 12.59 [9.25–18.28] | 17.60 [12.94–23.08] | **0.045** | **0.227** |
| **Indole-3-acetamide** | 2.18 [1.34–2.67] | 2.72 [1.92–3.80] | ***0.070*** | ***0.254*** |
| **Kynurenic acid** | 48.54 [36.48–66.72] | 49.74 [39.19–67.48] | 0.511 | 0.681 |
| **Indole-3-lactic acid** | 713.20 [632.56–957.12] | 759.67 [585.67–983.79] | 0.716 | 0.795 |
| **Indole-3-aldehyde** | 28.98 [20.22–32.95] | 29.79 [24.60–43.69] | ***0.089*** | ***0.254*** |
| **Indole-3-propionic acid** | 732.14 [587.42–1066.84] | 854.21 [486.91–1365.65] | 0.309 | 0.534 |
| **Indole-3-acetic acid** | 1681.81 [1300.38–1953.23] | 1721.93 [1239.762415.81] | 0.895 | 0,895 |

### Supplementary Table 2B. Short-Chain Fatty Acids (SCFA) Panel

Absolute concentrations of short-chain fatty acids quantified by LC-MS/MS. Values < LLOQ reported as ND. Concentrations in µmol/L. This table reports absolute measured concentrations; it contains no P values and no statistical tests were performed.

| **Metabolite** | **Non-CAS (median [IQR])** | **CAS (median [IQR])** | **p value** | **FDR q value** |
| --- | --- | --- | --- | --- |
| **Acetic acid** | 78971.14 [44712.00–123756.11] | 65249.62 [35851.20–119090.98] | 0.507 | 0.676 |
| **Propionic acid** | 1050.61 [795.53–1809.12] | 1301.83 [597.61–1858.07] | 0.903 | 0.903 |
| **Butyric acid** | 1758.74 [1423.72–2242.88] | 1888.87 [1510.90–2219.24] | 0.502 | 0.676 |
| **Isobutyric acid** | 355.52 [303.34–413.90] | 322.34 [302.68–395.07] | 0.272 | 0.676 |
| **Valeric acid** | 895.32 [838.32–1037.58] | 910.76 [881.55–970.61] | 0.772 | 0.882 |
| **Isovaleric acid** | 877.99 [715.00–1074.42] | 804.81 [620.44–1093.04] | 0.478 | 0.676 |

### Supplementary Table 2C. Bile Acids (BA) Panel

Absolute concentrations of bile acid species quantified by LC-MS/MS. Values < LLOQ reported as ND. Concentrations in nmol/L. This table reports absolute measured concentrations; it contains no P values and no statistical tests were performed.

| **Metabolite** | **Non-CAS (median [IQR])** | **CAS (median [IQR])** | **p value** | **FDR q value** |
| --- | --- | --- | --- | --- |
| **THCA** | 3.32 [1.99–7.10] | 4.45 [3.04–8.20] | 0.242 | 0.949 |
| **TCA** | 29.14 [11.98–68.16] | 25.04 [10.11–43.78] | 0.483 | 0.964 |
| **GHCA** | 12.03 [8.87–18.53] | 15.25 [8.26–22.36] | 0.516 | 0.964 |
| **GCA** | 180.95 [52.65–406.94] | 151.74 [84.11–261.15] | 0.964 | 0.964 |
| **TUDCA** | 8.84 [6.05–11.60] | 9.47 [5.45–21.10] | 0.761 | 0.964 |
| **TCDCA** | 59.54 [30.72–215.04] | 88.38 [33.01–163.85] | 0.795 | 0.964 |
| **TDCA** | 24.77 [12.24–99.06] | 23.48 [10.65–87.55] | 0.936 | 0.964 |
| **HCA** | 37.18 [13.83–51.92] | 19.37 [13.45–47.35] | 0.763 | 0.964 |
| **UDCA** | 6.91 [4.35–12.37] | 9.02 [5.71–15.06] | 0.259 | 0.949 |
| **CA** | 49.42 [22.62–140.06] | 38.93 [22.72–115.18] | 0.923 | 0.964 |
| **GUDCA** | 179.86 [98.96–335.83] | 208.72 [83.89–427.38] | 0.508 | 0.964 |
| **GCDCA** | 584.84 [284.19–1292.52] | 730.95 [313.42–1455.85] | 0.743 | 0.964 |
| **GDCA** | 197.93 [68.43–420.01] | 186.55 [79.01–386.41] | 0.909 | 0.964 |
| **TLCA** | 25.71 [12.80–36.24] | 16.51 [14.15–20.03] | 0.733 | 0.964 |
| **LCA-S** | 28.01 [15.31–67.75] | 14.33 [12.67–18.16] | 0.095 | 0.697 |
| **CDCA** | 130.44 [58.06–326.13] | 120.45 [72.75–328.57] | 0.758 | 0.964 |
| **GLCA** | 17.46 [14.21–21.98] | 13.90 [12.21–25.35] | 0.581 | 0.964 |
| **DCA** | 243.88 [126.15–389.08] | 249.90 [129.34–436.87] | 0.920 | 0.964 |
| **LCA** | 80.16 [59.37–115.72] | 62.23 [41.09–91.89] | 0.077 | 0,697 |

### Supplementary Table 2D. TMA/TMAO Panel

Absolute concentrations of TMA, TMAO, choline, and L-carnitine quantified by LC-MS/MS. Values < LLOQ reported as ND. Concentrations in µmol/L. This table reports absolute measured concentrations; it contains no P values and no statistical tests were performed.

| Metabolite | Non-CAS (median [IQR]) | CAS (median [IQR]) | p value | FDR q value |
| --- | --- | --- | --- | --- |
| TMA | 29.52 [20.80–47.06] | 26.95 [19.78–43.38] | 0.594 | 0.693 |
| Choline | 17048.78 [16258.06–19865.45] | 17578.38 [14858.92–20189.72] | 0.567 | 0.693 |
| TMAO | 3111.82 [1943.27–5098.19] | 3240.86 [1924.51–4484.40] | 0.795 | 0.795 |
| L-Carnitine | 48343.19 [41528.99–55213.63] | 52896.51 [41536.92–62868.13] | 0.211 | 0.530 |

### Supplementary Table 3. Comorbidity-Adjusted Sensitivity Analysis for Kynurenine-Pathway Metabolites

Sensitivity analysis examining robustness of kynurenine-pathway metabolite associations with CAS after adjustment for key clinical confounders (Sensitivity Analysis 6). Linear regression models with log₂-transformed metabolite concentrations as outcome. β = standardized CAS effect estimate (log-scale); p = p-value for the CAS term. P values were derived from multivariable linear regression models with log₂-transformed metabolite concentration as the outcome; the reported P value corresponds to the CAS-status coefficient.

| **Metabolite** | **Model** | **CAS beta (log)** | **CAS p** |
| --- | --- | --- | --- |
| **Kynurenine** | CAS only | 0.111 | 0.071 |
| **Kynurenine** | Adjusted for age + BMI | 0.107 | 0.077 |
| **Kynurenine** | + atrial fibrillation | 0.119 | 0.061 |
| **Kynurenine** | + anticoagulant use | 0.127 | **0.034** |
| **Kynurenine** | + diabetes | 0.113 | 0.064 |
| **3-OH-Anthranilic acid** | CAS only | 0.136 | **0.046** |
| **3-OH-Anthranilic acid** | Adjusted for age + BMI | 0.118 | 0.083 |
| **3-OH-Anthranilic acid** | + atrial fibrillation | 0.108 | 0.131 |
| **3-OH-Anthranilic acid** | + anticoagulant use | 0.132 | 0.054 |
| **3-OH-Anthranilic acid** | + diabetes | 0.115 | 0.095 |
| **Xanthurenic acid** | CAS only | 0.192 | 0.077 |
| **Xanthurenic acid** | Adjusted for age + BMI | 0.199 | 0.071 |
| **Xanthurenic acid** | + atrial fibrillation | 0.192 | 0.093 |
| **Xanthurenic acid** | + anticoagulant use | 0.218 | **0.049** |
| **Xanthurenic acid** | + diabetes | 0.195 | 0.080 |
| **Indole-3-Sulfate** | CAS only | **-0.249** | **0.024** |
| **Indole-3-Sulfate** | Adjusted for age + BMI | **-0.236** | **0.035** |
| **Indole-3-Sulfate** | + atrial fibrillation | **-0.267** | **0.023** |
| **Indole-3-Sulfate** | + anticoagulant use | **-0.242** | **0.034** |
| **Indole-3-Sulfate** | + diabetes | **-0.252** | **0.025** |

### Supplementary Table 4. Sensitivity Analyses: eGFR Adjustment (I3S) and Platelet Adjustment (Serotonin)

A. Indole-3-Sulfate (I3S) and eGFR Adjustment (Sensitivity Analysis 6): Addresses renal confounding. B. Serotonin and Platelet Count Adjustment (Sensitivity Analysis 7): Addresses serum-serotonin platelet confounding. P values were derived from multivariable linear regression adjusting for eGFR (Panel A) or platelet count (Panel B); the reported P value corresponds to the CAS-status coefficient.

#### A. Indole-3-Sulfate (I3S) and eGFR Adjustment

| **Model** | **β (CAS effect on I3S)** | **p-value** | **Interpretation** |
| --- | --- | --- | --- |
| **Base model (CAS effect, N=95)** | **-0.236** | **0.035** | Significant — I3S lower in CAS |
| **+ eGFR adjustment (N=95)** | **-0.278** | **0.013** | Signal strengthens — renal confounding refuted |
| **eGFR coefficient (in adjusted model)** | **-0.176** | **0.022** | eGFR independently predicts I3S |

#### B. Serotonin and Platelet Count Adjustment

| **Model** | **β (CAS effect on serotonin)** | **p-value** | **Interpretation** |
| --- | --- | --- | --- |
| **Base model (CAS effect)** | — | 0.807 | Serotonin null — CAS has no effect |
| **+ Platelet count adjustment** | **-0.007** | 0.96 | Remains null after platelet adjustment |
| **Platelet count coefficient** | +0.231 | **0.003** | Platelet count predicts serotonin (r=0.33) |

### Supplementary Table 5. KI Ratio vs. Indexed AVA: Single-Covariate Sensitivity Analysis

Robustness of the kynurenine-to-indole ratio vs. indexed aortic valve area association to single-covariate adjustment. Outcome: log₂(Kynurenine/Indole-3-Sulfate) vs. Indexed AVA (cm²/m²). Method: Partial Spearman ρ with successive single-covariate adjustments in CAS patients with available echocardiographic data (n = 46). P values were derived from partial Spearman rank correlations (rank-based ordinary-least-squares residualisation) between log₂(kynurenine/indole-3-sulfate) and indexed AVA, each adjusted for a single covariate.

| **Covariate** | **Type** | **n** | **Partial ρ** | **95% CI lower** | **95% CI upper** | **p-value** | **Significance** |
| --- | --- | --- | --- | --- | --- | --- | --- |
| **Unadjusted** | — | 46 | **-0.310** | **-0.540** | **-0.022** | **0.0306** | * |
| **Age** | continuous | 46 | **-0.307** | **-0.542** | **-0.013** | **0.0325** | * |
| **Sex** | binary (0=F, 1=M) | 46 | **-0.306** | **-0.533** | **0.000** | **0.0327** | * |
| **eGFR** | continuous | 46 | **-0.296** | **-0.549** | **0.006** | **0.0400** | * |
| **AF** | binary | 46 | **-0.300** | **-0.529** | **-0.013** | **0.0372** | * |
| **Diabetes** | binary | 46 | **-0.287** | **-0.549** | **-0.017** | **0.0468** | * |
| **BMI** | continuous | 46 | **-0.264** | **-0.490** | **0.045** | 0.0696 | ns |
| **Statin use** | binary | 46 | **-0.307** | **-0.532** | **-0.013** | **0.0326** | * |
| **Anticoagulant** | binary | 46 | **-0.301** | **-0.525** | **-0.003** | **0.0362** | * |

### Supplementary Table 6. BH-FDR Corrected Metabolite Comparisons by CAS Severity

BH-FDR correction applied within the severity stratum (18 QC-passing metabolites). P values were derived from Mann–Whitney U tests comparing severe vs. mild-moderate CAS; q values reflect Benjamini–Hochberg FDR correction applied within the severity stratum.

| **Metabolite class** | **Metabolite** | **Mild–Moderate AS (N = 16)** | **Severe AS (N = 39)** | **p value** | **FDR q value** |
| --- | --- | --- | --- | --- | --- |
| **Bile acids** | **THCA** | **5.78 [4.34–24.29]** | **4.55 [2.90–7.17]** | **0.230** | **0.817** |
|  | TCA | 10.87 [7.24–46.93] | 27.98 [11.37–43.80] | 0.400 | 0.832 |
|  | GHCA | 12.32 [8.58–26.98] | 17.26 [8.22–22.89] | 0.747 | 0.966 |
|  | GCA | 99.48 [58.94–211.66] | 161.32 [87.77–323.55] | 0.260 | 0.817 |
|  | TUDCA | 9.38 [5.04–30.02] | 9.56 [6.09–20.01] | 1.000 | 1.000 |
|  | TCDCA | 44.98 [30.61–161.77] | 109.96 [32.83–196.13] | 0.388 | 0.832 |
|  | TDCA | 31.85 [14.61–116.33] | 26.76 [9.68–87.55] | 0.539 | 0.966 |
|  | HCA | 16.37 [12.86–28.47] | 20.63 [13.12–43.37] | 0.684 | 0.966 |
|  | UDCA | 11.32 [7.64–19.62] | 8.98 [4.68–13.55] | 0.174 | 0.817 |
|  | CA | 34.67 [18.47–73.83] | 46.00 [22.98–358.48] | 0.228 | 0.817 |
|  | GUDCA | 243.18 [71.67–454.61] | 192.41 [86.13–438.18] | 0.991 | 1.000 |
|  | GCDCA | 380.94 [219.00–1232.12] | 928.81 [461.22–1531.22] | 0.165 | 0.817 |
|  | GDCA | 169.83 [99.66–418.05] | 178.83 [71.88–421.74] | 0.956 | 1.000 |
|  | TLCA | 18.16 [15.66–24.73] | 16.51 [13.23–18.88] | 0.648 | 0.966 |
|  | LCA-S | 15.54 [13.98–20.09] | 13.68 [12.00–17.09] | 0.366 | 0.832 |
|  | CDCA | 122.21 [72.70–146.68] | 120.21 [70.95–404.09] | 0.584 | 0.966 |
|  | GLCA | 16.46 [12.10–22.16] | 16.93 [12.42–29.05] | 0.733 | 0.966 |
|  | DCA | 269.46 [172.23–473.63] | 233.14 [102.74–389.76] | 0.197 | 0.817 |
|  | LCA | 46.34 [40.92–93.57] | 64.82 [41.09–79.45] | 1.000 | 1.000 |
| **SCFAs** | **Acetic acid** | **126371.34 [52113.32–168640.16]** | **61696.60 [29955.98–99528.56]** | **0.082** | **0.328** |
|  | **Propionic acid** | 1407.04 [992.43–2551.29] | 743.08 [552.95–1658.83] | **0.056** | 0.328 |
|  | Butyric acid | 1954.45 [1510.38–2217.95] | 1874.54 [1411.11–2164.55] | 0.670 | 0.802 |
|  | Isobutyric acid | 330.84 [307.07–360.93] | 321.41 [300.70–389.94] | 0.623 | 0.802 |
|  | Valeric acid | 921.46 [878.60–983.30] | 904.39 [882.48–942.96] | 0.477 | 0.802 |
|  | Isovaleric acid | 811.06 [596.40–994.04] | 790.07 [639.05–1042.83] | 0.702 | 0.802 |
| **TMA/TMAO & L-Carnitine** | **TMA** | **26.67 [16.89–39.01]** | **28.72 [20.73–43.43]** | **0.711** | **0.995** |
|  | Choline | 16715.99 [15596.07–20262.52] | 16640.69 [14328.61–20041.14] | 0.592 | 0.995 |
|  | TMAO | 3276.01 [2082.50–4380.75] | 2822.58 [1837.06–4469.21] | 0.607 | 0.995 |
|  | L-Carnitine | 55251.85 [42707.02–63479.26] | 53929.71 [41901.06–63457.75] | 0.887 | 1.000 |
| **Tryptophan derivatives** | **Picolinic acid** | **30.31 [16.48–47.00]** | **18.29 [13.75–24.46]** | **0.113** | **0.720** |
|  | 3-OH-Kynurenine | 43.91 [36.28–54.95] | 48.72 [39.11–60.96] | 0.225 | 0.799 |
|  | **Quinolinic acid** | 540.88 [361.62–653.90] | 714.51 [470.39–1131.67] | **0.045** | 0.454 |
|  | Serotonine | 526.88 [361.91–761.17] | 606.80 [356.86–724.19] | 0.853 | 1.000 |
|  | 5-OH-Tryptophane | 5.10 [4.57–5.49] | 5.43 [4.66–6.91] | 0.320 | 0.799 |
|  | **Kynurenine** | 1951.43 [1833.16–2364.60] | 2501.75 [2085.39–2907.90] | **0.031** | 0.454 |
|  | 3-OH-Anthranilic acid | 44.69 [38.83–53.44] | 50.05 [42.45–57.31] | 0.309 | 0.799 |
|  | Tryptophane | 62072.96 [50137.67–74595.40] | 67242.38 [58486.79–76821.70] | 0.425 | 0.849 |
|  | 5-OH-Indole acetic acid | 60.98 [40.26–68.38] | 58.45 [47.26–73.45] | 0.638 | 1.000 |
|  | Indole-3-Sulfate | 4094.52 [3378.13–5783.37] | 4043.19 [2714.17–6637.66] | 0.922 | 1.000 |
|  | N-acetyl-serotonine | 0.52 [0.52–0.52] | 0.51 [0.51–0.51] | — | — |
|  | Xanthurenic acid | 15.46 [11.44–17.66] | 17.98 [12.93–24.29] | 0.144 | 0.720 |
|  | Indole-3-acetamide | 2.95 [2.22–4.50] | 2.55 [1.97–3.52] | 0.756 | 1.000 |
|  | Kynurenic acid | 49.83 [41.12–59.60] | 47.59 [37.02–67.62] | 0.785 | 1.000 |
|  | Indole-3-Lactic acid | 740.00 [600.65–931.94] | 750.97 [564.00–984.99] | 0.735 | 1.000 |
|  | Indole-3-Aldehyde | 29.79 [24.47–42.47] | 30.29 [25.20–43.82] | 0.904 | 1.000 |
|  | Tryptophol | 1.79 [1.79–1.79] | — | — | — |
|  | Indole-3-propionic acid | 774.02 [470.25–1117.37] | 806.42 [486.35–1180.04] | 0.751 | 1.000 |

### Supplementary Table 7. Clinical Characteristics by CAS Severity

Baseline characteristics of CAS patients by severity: Severe CAS (n = 32) vs. Mild-Moderate CAS (n = 16). Continuous variables: median [IQR]; Mann-Whitney U test. Categorical variables: n (%); Fisher exact test. Severity defined as: Severe = AVA ≤ 1.0 cm² or mean gradient ≥ 40 mmHg; Mild-Moderate = AVA > 1.0 cm². P values for continuous variables were derived from the Mann–Whitney U test and for categorical variables from Fisher’s exact test.

| **Variable** | **Non-severe CAS (N = 16)** | **Severe CAS (N = 32)** | **p value** |
| --- | --- | --- | --- |
| **Demographics** |  |  |  |
| **Age, years** | 67.0 [61.5–76.3] | 64.0 [57.5–70.3] | 0.304 |
| **BMI, kg/m²** | 28.41 [25.94–31.86] | 28.57 [27.09–33.74] | 0.638 |
| **Female, n (%)** | 0 (0.0%) | 12 (37.5%) | - |
| **Clinical measurements** |  |  |  |
| **SBP, mmHg** | 138.5 [128.3–143.0] | 134.0 [125.0–139.3] | 0.185 |
| **DBP, mmHg** | 76.5 [65.8–82.5] | 76.5 [68.0–83.3] | 0.895 |
| **Heart rate, bpm** | 70.0 [66.0–77.0] | 71.0 [62.0–81.3] | 0.891 |
| **Laboratory parameters** |  |  |  |
| **Hemoglobin, g/dL** | 14.2 [13.3–15.7] | 14.1 [12.6–14.9] | 0.208 |
| **Leukocytes, ×10⁹/L** | 7.43 [6.61–8.62] | 6.34 [5.32–8.59] | 0.115 |
| **Platelets, ×10⁹/L** | 207 [176–229] | 223 [195–255] | 0.186 |
| **GFR, mL/min/1.73m²** | 90.6 [83.0–120.0] | 103.7 [85.4–120.4] | 0.818 |
| **Total cholesterol, mmol/L** | 2.99 [1.92–4.09] | 4.20 [3.57–5.22] | **0.010** |
| **LDL, mmol/L** | 1.33 [0.79–2.53] | 2.34 [1.48–3.54] | **0.021** |
| **HDL, mmol/L** | 1.02 [0.47–1.37] | 1.22 [1.01–1.50] | 0.058 |
| **BNP, pg/mL** | 215.5 [66.3–535.5] | 238.0 [149.0–538.3] | 0.459 |
| **Echocardiography** |  |  |  |
| **LVEF, %** | 59.0 [55.0–60.0] | 60.0 [60.0–63.5] | **0.021** |
| **AV max gradient, mmHg** | 33.0 [25.0–43.0] | 65.0 [49.0–80.5] | **<0.001** |
| **AV mean gradient, mmHg** | 21.5 [14.8–28.5] | 44.0 [36.5–55.0] | **<0.001** |
| **AV peak velocity, m/s** | 2.92 [2.48–3.21] | 3.91 [3.41–4.24] | **0.001** |
| **AV area, cm²** | 1.38 [1.06–1.71] | 0.83 [0.76–0.96] | **<0.001** |
| **AV area index, cm²/m²** | 0.62 [0.51–0.85] | 0.41 [0.37–0.48] | **<0.001** |
| **Comorbidities, n (%)** |  |  |  |
| **CAD** | 6 (37.5%) | 7 (21.9%) | 0.421 |
| **PCI** | 4 (25.0%) | 1 (3.1%) | 0.066 |
| **Prior MI** | 2 (12.5%) | 1 (3.1%) | 0.527 |
| **Cerebrovascular disease** | 3 (18.8%) | 1 (3.1%) | 0.196 |
| **Hypertension** | 10 (62.5%) | 17 (53.1%) | 0.758 |
| **Dyslipidemia** | 6 (40.0%) | 14 (43.8%) | 1.000 |
| **Diabetes mellitus** | 2 (12.5%) | 3 (9.4%) | 1.000 |
| **COPD** | 1 (6.2%) | 1 (3.1%) | 1.000 |
| **Statin use** | 12 (75.0%) | 17 (53.1%) | 0.251 |

### Supplementary Table 8. Clinical Characteristics by Sex within CAS Patients

Baseline characteristics of CAS patients by sex: Male CAS (n = 41) vs. Female CAS (n = 13). Continuous variables: median [IQR]; Mann-Whitney U test. Categorical variables: n (%); Fisher exact test. P values for continuous variables were derived from the Mann–Whitney U test and for categorical variables from Fisher’s exact test.

| **Clinical data** | **Men (N=41)** | **Women (N=13)** | **p value** |
| --- | --- | --- | --- |
| **Age, years** | 67.0 [60.0–76.0] | 65.0 [60.0–69.0] | 0.460 |
| **Body mass index, kg/m²** | 28.0 [26.8–30.4] | 32.0 [25.3–35.2] | 0.430 |
| **Systolic blood pressure, mmHg** | 136 [126–144] | 131 [113–138] | 0.202 |
| **Diastolic blood pressure, mmHg** | 78 [68–84] | 73 [68–83] | 0.478 |
| **Heart rate, bpm** | 70 [64.5–81.3] | 72 [62–76] | 0.893 |
| **Biological data** |  |  |  |
| **eGFR, mL/min/1.73 m²** | 101.1 [73.2–122.7] | 89.3 [80.1–107.2] | 0.492 |
| **BNP, pg/mL** | 203 [62–447] | 208 [177–598] | 0.561 |
| **Troponin, ng/L** | 15.0 [10.3–26.8] | 12.5 [7.0–21.8] | 0.425 |
| **Alkaline phosphatase, U/L** | 62 [52–79.5] | 74 [68–102] | 0.063 |
| **Total cholesterol, mmol/L** | 3.90 [2.49–5.05] | 4.51 [3.98–5.66] | **0.041** |
| **LDL cholesterol, mmol/L** | 1.79 [1.10–2.83] | 2.39 [1.74–3.61] | 0.135 |
| **Triglycerides, mmol/L** | 1.35 [0.86–1.80] | 1.38 [0.97–1.90] | 0.895 |
| **HDL cholesterol, mmol/L** | 1.14 [0.79–1.44] | 1.41 [1.22–1.59] | **0.025** |
| **Lipoprotein(a), mg/dL** | 20.0 [7.0–57.8] | 21.5 [20.0–111.5] | 0.407 |
| **Fasting glucose, g/L** | 1.02 [0.93–1.11] | 1.02 [0.95–1.06] | 0.780 |
| **HbA1c, %** | 5.6 [5.3–6.6] | 5.6 [5.5–5.7] | 0.863 |
| **Echocardiographic parameters** |  |  |  |
| **LVEF (%)** | 60.00 [55.00–62.00] | 60.00 [60.00–62.00] | 0.327 |
| **Aortic valve max gradient (mmHg)** | 40.00 [25.00–65.00] | 62.00 [59.00–72.00] | **0.049** |
| **Aortic valve mean gradient (mmHg)** | 28.00 [16.50–46.50] | 43.00 [38.00–44.00] | 0.108 |
| **Peak velocity (m/s)** | 3.20 [2.60–4.00] | 3.92 [3.90–4.12] | ***0.052*** |
| **Aortic valve area (cm²)** | 1.00 [0.82–1.38] | 0.80 [0.79–0.89] | **0.009** |
| **Aortic valve area index (cm²/m²)** | 0.48 [0.39–0.65] | 0.42 [0.39–0.48] | 0.216 |
| **CT calcium score** | 1613.00 [439.00–3336.25] | 1340.00 [1259.00–2396.00] | 0.836 |
| **Cardiovascular risk factors** |  |  |  |
| **Hypertension** | 25 (61.0%) | 7 (53.8%) | 0.90 |
| **Dyslipidemia** | 19 (47.5%) | 4 (30.8%) | 0.46 |
| **Diabetes mellitus** | 6 (14.6%) | 0 (0.0%) | 0.34 |
| **Coronary artery disease** | 13 (31.7%) | 1 (7.7%) | 0.17 |
| **Active smoking** | 3 (7.3%) | 1 (7.7%) | 1.00 |

### Supplementary Table 9. BH-FDR Corrected Tryptophan Metabolite Comparisons by Sex

BH-FDR correction applied within each sex-specific stratum (18 QC-passing metabolites per stratum). Significance key: * = q < 0.05; † = 0.05 ≤ q < 0.1 (trend). Analyses performed within the matched cohort (Male CAS n = 41, Male Ctrl n = 34, Female CAS n = 13, Female Ctrl n = 7). P values were derived from Mann–Whitney U tests within each sex stratum; q values reflect Benjamini–Hochberg FDR correction applied within each stratum.

| **Metabolite** | **Male CAS vs Ctrl p** | **Male CAS vs Ctrl q** | **Female CAS vs Ctrl p** | **Female CAS vs Ctrl q** | **Male vs Female (CAS) p** | **Male vs Female (CAS) q** | **Male Ctrl vs Female Ctrl p** | **Male Ctrl vs Female Ctrl q** |
| --- | --- | --- | --- | --- | --- | --- | --- | --- |
| **Picolinic acid** | 0.300 | 0.666 | — | 1.0 | **0.012** | **0.131** | 0.465 | 0.782 |
| **3-OH-Kynurenine** | 0.349 | 0.666 | 0.968 | 1.0 | 0.564 | 0.966 | 0.436 | 0.782 |
| **Quinolinic acid** | 0.624 | 0.771 | 0.905 | 1.0 | 0.723 | 1.0 | 0.232 | 0.782 |
| **Serotonine** | 0.106 | 0.444 | 0.905 | 1.0 | 0.976 | 1.0 | 0.499 | 0.782 |
| **5-OH-Tryptophane** | 0.392 | 0.685 | 0.905 | 1.0 | 0.800 | 1.0 | 0.522 | 0.782 |
| **Kynurenine** | **0.022** | **0.185** | 0.452 | 1.0 | 0.551 | 0.966 | 0.080 | 0.721 |
| **Tryptamine** | 0.859 | 1.0 | 0.462 | 1.0 | 0.644 | 0.966 | 0.391 | 0.782 |
| **3-OH-Anthranilic acid** | **0.009** | **0.185** | 0.721 | 1.0 | 0.307 | 0.645 | 0.456 | 0.782 |
| **Tryptophane** | 0.322 | 0.666 | 0.501 | 1.0 | 0.863 | 1.0 | **0.030** | 0.546 |
| **5-OH-Indole acetic acid** | 0.456 | 0.734 | 0.501 | 1.0 | 0.620 | 0.966 | 0.591 | 0.791 |
| **Indole-3-Sulfate** | 0.154 | 0.461 | 0.104 | 1.0 | 0.199 | 0.597 | 0.849 | 0.901 |
| **N-acetyl-serotonine** | — | 1.0 | — | 1.0 | — | 1.0 | — | — |
| **Xanthurenic acid** | **0.026** | **0.185** | 0.866 | 1.0 | 0.180 | 0.597 | 0.851 | 0.901 |
| **Indole-3-acetamide** | 0.140 | 0.461 | 0.193 | 1.0 | **0.058** | **0.242** | 0.334 | 0.782 |
| **Kynurenic acid** | 0.287 | 0.666 | 0.905 | 1.0 | **0.009** | **0.131** | 0.275 | 0.782 |
| **Indole-3-Lactic acid** | 0.489 | 0.734 | 0.606 | 1.0 | **0.025** | **0.178** | 0.377 | 0.782 |
| **Indole-3-Aldehyde** | **0.036** | 0.189 | 0.843 | 1.0 | **0.040** | **0.21** | 0.615 | 0.791 |
| **Tryptophol** | — | 1.0 | — | 1.0 | — | 1.0 | — | — |
| **Melatonine** | — | 1.0 | — | 1.0 | — | 1.0 | — | — |
| **Indole-3-propionic acid** | 0.537 | 0.752 | 0.663 | 1.0 | 0.253 | 0.645 | 0.931 | 0.931 |
| **Indole-3-Acetic acid** | 0.624 | 0.771 | 0.663 | 1.0 | 0.288 | 0.645 | 0.716 | 0.859 |

### Supplementary Table 10. Forward MR Lipoprotein(a) Wald Ratio: Confirmed Values

Confirmed values for lipoprotein(a) fallback Wald-ratio Mendelian randomization estimate. This single-SNP Wald-ratio estimate is provided as a supplementary reference. IVW multi-SNP MR results for lipoprotein(a) are reported in Supplementary Table 12 (forward MR) and are the primary evidence used in the manuscript. The P value was derived from a single-SNP Wald-ratio Mendelian randomization estimate (Wald ratio z test).

| **Field** | **Value** |
| --- | --- |
| **Method** | Wald ratio (single cis-SNP, rs10455872) |
| **SNPs** | 1 |
| **OR** | 1.116 |
| **95% CI** | [1.106–1.125] |
| **p-value** | **< 0.001** |
| **F-statistic** | 77,829 |
| **Weighted median** | N/A (single-SNP instrument) |
| **MR-Egger** | N/A (single-SNP instrument) |

Supplementary Table 11. **Weighted median Mendelian randomization analysis of circulating biomarkers on calcific aortic stenosis (CAS).**

This table presents causal estimates derived from weighted median Mendelian randomization analyses assessing the effect of circulating biomarkers on CAS. For each marker, the number of SNPs used as genetic instruments, odds ratios (OR), 95% confidence intervals (CI), and P-values are reported. P values were derived from weighted median Mendelian randomization estimates.

Markers labeled as “weak instrument” had fewer than three independent SNPs or insufficient genetic strength and were not included in the weighted median estimation.

The weighted median method provides consistent causal estimates even when up to 50% of the genetic instruments are invalid, offering robustness to horizontal pleiotropy.

† For lipoprotein(a), the multi-SNP IVW instrument could not be harmonized via rsid (CAS GWAS stores rs10455872 under a post-liftover rsid); additionally, the 9-SNP instrument exhibited extreme heterogeneity (I²=98.8%). In accordance with the pre-specified fallback strategy (Methods), the estimate was derived as a Wald ratio using rs10455872 (F=77,829) harmonized by chromosomal position. Weighted Median and MR-Egger are not computable for a single-SNP instrument.

| **Biomarker** | **SNPs (MR)** | **Beta** | **95% CI** | **P-value** | **Mean F-statistic** |
| --- | --- | --- | --- | --- | --- |
| **ALPL** | 15 | 0.273 | 0.207–0.340 | 8.88 × 10⁻¹⁶ | 158.63 |
| **IFNγ receptor 1** | 33 | 1.437 | 0.967–1.908 | 2.19 × 10⁻⁹ | 43.97 |
| **Indole acetate** | 8 | 0.263 | 0.161–0.365 | 4.66 × 10⁻⁷ | 36.36 |
| **LDL cholesterol** | 41 | 0.084 | 0.048–0.121 | 5.59 × 10⁻⁶ | 117.93 |
| **IFNγ** | 33 | 0.737 | 0.266–1.208 | 2.16 × 10⁻³ | 31.86 |
| **Kynurenine** | 8 | **-0.088** | -0.150 to -0.027 | 4.88 × 10⁻³ | 54.40 |
| **Lipoprotein(a)** | 15 | **-0.095** | -0.170 to -0.021 | 1.24 × 10⁻² | 669.24 |
| **IL18** | 39 | **-0.249** | -0.461 to -0.037 | 2.14 × 10⁻² | 101.22 |
| **IL6 receptor subunit** | 40 | **-0.221** | -0.425 to -0.016 | 3.41 × 10⁻² | 867.05 |
| **BMPR1A** | 33 | 0.443 | -0.027 to 0.914 | 6.49 × 10⁻² | 0.00 (no SNP) |
| **IL18 receptor** | 33 | **-0.422** | -0.892 to 0.049 | 7.94 × 10⁻² | 151.94 |
| **Indole propionate** | 8 | 0.091 | -0.029 to 0.211 | 1.39 × 10⁻¹ | 60.21 |
| **Osteopontin** | 34 | **-0.502** | -1.398 to 0.395 | 2.73 × 10⁻¹ | 35.72 |
| **Serotonin** | 8 | 0.056 | -0.058 to 0.170 | 3.37 × 10⁻¹ | 0.00 (no SNP) |
| **IL6** | 35 | 0.078 | -0.176 to 0.333 | 5.47 × 10⁻¹ | 70.92 |
| **IDO** | 33 | **0.026** | -0.445 to 0.496 | 9.15 × 10⁻¹ | 69.99 |
| **Tryptophan** | 8 | **-0.002** | -0.067 to 0.064 | 9.65 × 10⁻¹ | 36.21 |
| **IL1β** | 32 | **0.009** | -0.400 to 0.418 | 9.66 × 10⁻¹ | 0.00 (no SNP) |

Supplementary Table 12. Weighted median Mendelian randomization analysis of calcific aortic stenosis (CAS) on circulating biomarkers. P values were derived from weighted median Mendelian randomization estimates.

| **Biomarker** | **SNPs (n)** | **IVW OR** | **Weighted Median OR** | **MR-Egger OR** | **Egger Intercept P-value** | **Egger Slope P-value** | **Consistency** |
| --- | --- | --- | --- | --- | --- | --- | --- |
| **IL6 receptor subunit** | 7 | 0.977 | 0.970 | 1.890 | 0.192 | 0.257 | Consistent |
| **LDL cholesterol** | 639 | 1.009 | 1.003 | 0.932 | 0.675 | 0.632 | Consistent |
| **ALPL** | 155 | 0.955 | 0.957 | 0.844 | 0.327 | **<0.001** | Consistent |
| **Lipoprotein(a) †** | 1 | 1,116 | N/A | N/A | single-SNP instrument | - | - |
| **IL18 receptor** | 4 | 0.849 | 0.653 | 1.329 | 0.394 | 0.634 | Divergent |
| **IL18** | 6 | 0.979 | 0.930 | **0.005** | 0.109 | 0.136 | Consistent |

Supplementary Table 13. **Sensitivity and robustness analyses for direct Mendelian randomization of circulating biomarkers on calcific aortic stenosis (CAS).**

This table presents causal estimates obtained using three complementary Mendelian randomization methods: inverse-variance weighted (IVW), weighted median, and MR-Egger regression. Odds ratios (OR) derived from each method are reported alongside MR-Egger intercept and slope P-values. P values were derived from inverse-variance weighted (IVW), weighted median, and MR-Egger regression estimates; the MR-Egger intercept P value tests for directional horizontal pleiotropy.

The MR-Egger intercept test was used to assess directional horizontal pleiotropy, while the MR-Egger slope provides a pleiotropy-adjusted causal estimate. Consistency across methods was evaluated qualitatively, with “consistent” indicating concordant direction and magnitude of effect across IVW, weighted median, and MR-Egger estimates, and “divergent” indicating discrepancies between methods.

The use of multiple MR approaches allows assessment of robustness under different assumptions regarding instrument validity.

† For lipoprotein(a), the multi-SNP IVW instrument could not be harmonized via rsid (CAS GWAS stores rs10455872 under a post-liftover rsid); additionally, the 9-SNP instrument exhibited extreme heterogeneity (I²=98.8%). In accordance with the pre-specified fallback strategy (Methods), the estimate was derived as a Wald ratio using rs10455872 (F=77,829) harmonized by chromosomal position. Weighted Median and MR-Egger are not computable for a single-SNP instrument.

| **Biomarker** | **SNPs (n)** | **IVW OR** | **Weighted Median OR** | **MR-Egger OR** | **Egger Intercept P-value** | **Egger Slope P-value** | **Consistency** |
| --- | --- | --- | --- | --- | --- | --- | --- |
| **IDO** | 33 | 1.021 | 1.026 | 0.905 | 0.552 | 0.669 | Consistent |
| **IL1β** | 32 | 0.907 | 1.009 | 0.562 | 0.471 | 0.314 | Consistent |
| **IL6** | 35 | 0.809 | 1.081 | 0.711 | 0.878 | 0.297 | Divergent |
| **IL6 receptor subunit** | 40 | 0.707 | 0.802 | 0.399 | 0.287 | 0.075 | Consistent |
| **Indole propionate** | 8 | 1.102 | 1.095 | 0.980 | 0.696 | 0.918 | Consistent |
| **Indole acetate** | 8 | 1.210 | 1.300 | 1.747 | **0.005** | **<0.001** | Consistent |
| **Kynurenine** | 8 | 0.946 | 0.916 | 0.804 | **0.005** | **<0.001** | Consistent |
| **LDL cholesterol** | 41 | 1.047 | 1.088 | 1.025 | 0.794 | 0.772 | Consistent |
| **Lipoprotein(a)** | 15 | 0.991 | 0.909 | 1.937 | 0.552 | 0.741 | Consistent |
| **Tryptophan** | 8 | 1.021 | 0.999 | 0.950 | 0.064 | 0.110 | Consistent |
| **Serotonin** | 8 | 1.026 | 1.058 | 1.170 | 0.107 | **0.036** | Consistent |
| **IFNγ receptor 1** | 33 | 2.271 | 4.210 | 0.884 | 0.366 | 0.876 | Divergent |
| **IFNγ** | 33 | 1.606 | 2.090 | 2.440 | 0.172 | **0.002** | Divergent |
| **IL18 receptor** | 33 | 0.694 | 0.656 | 0.813 | 0.267 | 0.449 | Consistent |
| **IL18** | 39 | 0.780 | 0.779 | 0.421 | **0.029** | **0.008** | Consistent |
| **ALPL** | 15 | 1.199 | 1.314 | 0.121 | 0.152 | 0.231 | Consistent |
| **BMPR1A** | 33 | 1.048 | 1.558 | 1.433 | 0.400 | 0.421 | Divergent |
| **Osteopontin** | 34 | 0.771 | 0.606 | 0.069 | 0.799 | 0.522 | Divergent |

Supplementary Table 14. **Sensitivity and robustness analyses for reverse Mendelian randomization of calcific aortic stenosis (CAS) on circulating biomarkers.** P values were derived from inverse-variance weighted (IVW), weighted median, and MR-Egger regression estimates; the MR-Egger intercept P value tests for directional horizontal pleiotropy.

| **Biomarker** | **SNPs (n)** | **Q Statistic** | **P-value (Q)** | **Heterogeneity** |
| --- | --- | --- | --- | --- |
| **LDL cholesterol** | 639 | 1383.20 | **< 1.0 × 10⁻¹⁶** | High |
| **Lipoprotein(a)** | N/A | single-SNP instrument; heterogeneity not computable | | |
| **ALPL** | 155 | 270.39 | 2.06 × 10⁻⁸ | High |
| **Indole propionate** | 2 | 5.58 | 1.82 × 10⁻² | High |
| **IL6** | 2 | 2.30 | 1.29 × 10⁻¹ | Low |
| **IL18** | 6 | 7.68 | 1.75 × 10⁻¹ | Low |
| **IL6 receptor subunit** | 7 | 8.18 | 2.25 × 10⁻¹ | Low |
| **IL18 receptor** | 4 | 3.53 | 3.16 × 10⁻¹ | Low |

Supplementary Table 15. **Cochran’s Q test for heterogeneity in Mendelian randomization analyses of circulating biomarkers on calcific aortic stenosis (CAS).**

This table presents Cochran’s Q statistics assessing heterogeneity among instrumental variables in direct Mendelian randomization analyses, where circulating biomarkers were considered as exposures and CAS as the outcome. P values were derived from Cochran’s Q heterogeneity test across instrumental variables.

The Q statistic evaluates variability in causal estimates across SNPs. A statistically significant Q test (P < 0.05) indicates the presence of heterogeneity

| **Biomarker** | **SNPs (n)** | **Q Statistic** | **P-value (Q)** | **Heterogeneity** |
| --- | --- | --- | --- | --- |
| **Lipoprotein(a)** | 15 | 88.72 | 6.62 × 10⁻¹³ | High |
| **ALPL** | 15 | 37.40 | 6.43 × 10⁻⁴ | High |
| **IFNγ receptor 1** | 33 | 55.89 | 5.58 × 10⁻³ | High |
| **Indole acetate** | 8 | 10.10 | 1.83 × 10⁻¹ | Low |
| **IL1β** | 32 | 37.69 | 1.90 × 10⁻¹ | Low |
| **IL6 receptor subunit** | 40 | 41.44 | 3.65 × 10⁻¹ | Low |
| **Kynurenine** | 8 | 5.79 | 5.64 × 10⁻¹ | Low |
| **LDL cholesterol** | 41 | 31.95 | 8.14 × 10⁻¹ | Low |
| **BMPR1A** | 33 | 24.37 | 8.31 × 10⁻¹ | Low |
| **Serotonin** | 8 | 3.49 | 8.36 × 10⁻¹ | Low |
| **IL6** | 35 | 23.60 | 9.09 × 10⁻¹ | Low |
| **Osteopontin** | 34 | 20.69 | 9.53 × 10⁻¹ | Low |
| **Indole propionate** | 8 | 2.07 | 9.56 × 10⁻¹ | Low |
| **Tryptophan** | 8 | 1.33 | 9.88 × 10⁻¹ | Low |
| **IFNγ** | 33 | 14.93 | 9.96 × 10⁻¹ | Low |
| **IL18** | 39 | 13.51 | 9.99 × 10⁻¹ | Low |
| **IL18 receptor** | 33 | 8.51 | 1.00 | Low |
| **IDO** | 33 | 4.54 | 1.00 | Low |

Supplementary Table 16. **Cochran’s Q test for heterogeneity in reverse Mendelian randomization analyses of calcific aortic stenosis (CAS) on circulating biomarkers.** P values were derived from Cochran’s Q heterogeneity test across instrumental variables.

| **Biomarker** | **FE Beta** | **P-value (FE)** | **RE Beta** | **P-value (RE)** | **Heterogeneity Factor (Φ)** | **Robust to Heterogeneity** |
| --- | --- | --- | --- | --- | --- | --- |
| **IL6 receptor subunit** | **-0.023** | 4.39 × 10⁻⁷ | **-0.023** | 1.50 × 10⁻⁵ | 1.17 | Yes |
| **IL6** | **-0.161** | 2.74 × 10⁻⁶ | **-0.161** | 2.00 × 10⁻³ | 1.52 | Yes |
| **ALPL** | **-0.046** | 6.91 × 10⁻³ | **-0.046** | 4.15 × 10⁻² | 1.33 | Yes |
| **IL18 receptor** | **-0.164** | 1.17 × 10⁻¹ | **-0.164** | 1.49 × 10⁻¹ | 1.09 | No |
| **Indole propionate** | **-0.271** | 9.20 × 10⁻² | **-0.271** | 4.75 × 10⁻¹ | 2.36 | No |
| **LDL cholesterol** | **0.009** | 3.15 × 10⁻¹ | **0.009** | 4.95 × 10⁻¹ | 1.47 | No |
| **IL18** | **-0.021** | 5.98 × 10⁻¹ | **-0.021** | 6.70 × 10⁻¹ | 1.24 | No |
| **Lipoprotein(a)** | **0.000** | Wald ratio; FE = RE for single SNP | | | | |

Supplementary Table 17. **Comparison of fixed-effects and random-effects inverse-variance weighted estimates in Mendelian randomization analyses of circulating biomarkers on calcific aortic stenosis (CAS).**

This table compares causal effect estimates obtained using fixed-effects (FE) and multiplicative random-effects (RE) inverse-variance weighted models in direct Mendelian randomization analyses. P values were derived from inverse-variance weighted Mendelian randomization under fixed-effects and multiplicative random-effects models.

The heterogeneity factor (Φ) reflects overdispersion due to variability across SNP-specific estimates. Random-effects models account for heterogeneity by inflating standard errors.

Associations were considered robust when effect estimates remained consistent and statistically significant under both FE and RE models.

| **Biomarker** | **FE Beta** | **P-value (FE)** | **RE Beta** | **P-value (RE)** | **Heterogeneity Factor (Φ)** | **Robust to Heterogeneity** |
| --- | --- | --- | --- | --- | --- | --- |
| **ALPL** | 0.182 | 1.31 × 10⁻¹⁰ | 0.182 | 8.40 × 10⁻⁵ | 1.63 | Yes |
| **IL6 receptor subunit** | **-0.347** | 6.41 × 10⁻⁵ | **-0.347** | 1.06 × 10⁻⁴ | 1.03 | Yes |
| **Indole acetate** | 0.191 | 1.11 × 10⁻⁵ | 0.191 | 2.55 × 10⁻⁴ | 1.20 | Yes |
| **IFNγ receptor 1** | 0.820 | 4.21 × 10⁻⁵ | 0.820 | 1.94 × 10⁻³ | 1.32 | Yes |
| **LDL cholesterol** | **0.046** | 3.22 × 10⁻³ | **0.046** | 3.22 × 10⁻³ | 1.00 | Yes |
| **IL18** | **-0.249** | 5.87 × 10⁻³ | **-0.249** | 5.87 × 10⁻³ | 1.00 | Yes |
| **IFNγ** | 0.474 | 1.79 × 10⁻² | 0.474 | 1.79 × 10⁻² | 1.00 | Yes |
| **Kynurenine** | **-0.055** | 3.51 × 10⁻² | **-0.055** | 3.51 × 10⁻² | 1.00 | Yes |
| **IL6** | **-0.212** | 5.07 × 10⁻² | **-0.212** | 5.07 × 10⁻² | 1.00 | No |
| **Indole propionate** | 0.097 | 5.65 × 10⁻² | 0.097 | 5.65 × 10⁻² | 1.00 | No |
| **IL18 receptor** | **-0.365** | 6.81 × 10⁻² | **-0.365** | 6.81 × 10⁻² | 1.00 | No |
| **Tryptophan** | **0.020** | 4.63 × 10⁻¹ | **0.020** | 4.63 × 10⁻¹ | 1.00 | No |
| **Osteopontin** | **-0.260** | 4.95 × 10⁻¹ | **-0.260** | 4.95 × 10⁻¹ | 1.00 | No |
| **Serotonin** | **0.025** | 6.04 × 10⁻¹ | **0.025** | 6.04 × 10⁻¹ | 1.00 | No |
| **IL1β** | **-0.097** | 5.77 × 10⁻¹ | **-0.097** | 6.13 × 10⁻¹ | 1.10 | No |
| **BMPR1A** | **0.047** | 8.14 × 10⁻¹ | **0.047** | 8.14 × 10⁻¹ | 1.00 | No |
| **Lipoprotein(a)** | **-0.009** | 7.83 × 10⁻¹ | **-0.009** | 9.13 × 10⁻¹ | 2.52 | No |
| **IDO** | **0.021** | 9.16 × 10⁻¹ | **0.021** | 9.16 × 10⁻¹ | 1.00 | No |

Supplementary Table 18. **Comparison of fixed-effects and random-effects inverse-variance weighted estimates in reverse Mendelian randomization analyses of calcific aortic stenosis (CAS) on circulating biomarkers.**

This table compares causal estimates derived from fixed-effects (FE) and multiplicative random-effects (RE) IVW models in reverse Mendelian randomization analyses. P values were derived from inverse-variance weighted Mendelian randomization under fixed-effects and multiplicative random-effects models.

Random-effects models account for between-SNP heterogeneity by incorporating an overdispersion parameter (Φ). Consistency between FE and RE estimates supports the robustness of causal inference despite potential heterogeneity.

| **Biomarker** | **Max P-value** | **Min P-value** | **Stability** | **Notes** |
| --- | --- | --- | --- | --- |
| **ALPL** | 1.43 × 10⁻² | 3.13 × 10⁻³ | Stable | P ranged from 0.0031 to 0.0143 |
| **IL6 receptor subunit** | 2.10 × 10⁻¹ | 1.73 × 10⁻⁷ | Fragile | P ranged from 0.0000 to 0.2099 |
| **IL18 receptor** | 3.18 × 10⁻¹ | 8.30 × 10⁻² | Fragile | P ranged from 0.0830 to 0.3183 |
| **LDL cholesterol** | 6.09 × 10⁻¹ | 1.05 × 10⁻¹ | Fragile | P ranged from 0.1053 to 0.6086 |
| **IL18** | 9.83 × 10⁻¹ | 4.38 × 10⁻¹ | Fragile | P ranged from 0.4378 to 0.9832 |

Supplementary Table 19. **Leave-one-out sensitivity analysis for Mendelian randomization of circulating biomarkers on calcific aortic stenosis (CAS).**

This table presents leave-one-out (LOO) analyses performed to evaluate the influence of individual SNPs on causal estimates in direct Mendelian randomization analyses. The reported P-value range was obtained from inverse-variance weighted Mendelian randomization repeated with each SNP removed in turn (leave-one-out analysis).

Each SNP was sequentially removed, and the IVW analysis was repeated to assess the stability of the association. The range of P-values obtained across all iterations is reported.

Associations were considered stable when results remained statistically consistent across all iterations. Large variability in P-values suggests that the association may be driven by one or a few influential SNPs.

| **Biomarker** | **Max P-value** | **Min P-value** | **Stability** | **Notes** |
| --- | --- | --- | --- | --- |
| **ALPL** | 1.00 × 10⁻⁶ | 1.31 × 10⁻¹¹ | Stable | P ranged from ~0.0000 to 0.0000 |
| **Indole acetate** | 1.64 × 10⁻⁴ | 5.50 × 10⁻⁷ | Stable | P ranged from 0.0000 to 0.0002 |
| **IFNγ receptor 1** | 1.99 × 10⁻⁴ | 9.07 × 10⁻⁶ | Stable | P ranged from 0.0000 to 0.0002 |
| **IL6 receptor subunit** | 3.86 × 10⁻⁴ | 2.91 × 10⁻⁵ | Stable | P ranged from 0.0000 to 0.0004 |
| **LDL cholesterol** | 6.77 × 10⁻³ | 1.20 × 10⁻³ | Stable | P ranged from 0.0012 to 0.0068 |
| **IL18** | 1.25 × 10⁻² | 3.22 × 10⁻³ | Stable | P ranged from 0.0032 to 0.0125 |
| **IFNγ** | 3.58 × 10⁻² | 6.67 × 10⁻³ | Stable | P ranged from 0.0067 to 0.0358 |
| **IL18 receptor** | 8.96 × 10⁻² | 4.44 × 10⁻² | Fragile | P ranged from 0.0444 to 0.0896 |
| **Kynurenine** | 9.65 × 10⁻² | 5.96 × 10⁻³ | Fragile | P ranged from 0.0060 to 0.0965 |
| **Indole propionate** | 1.28 × 10⁻¹ | 3.73 × 10⁻² | Fragile | P ranged from 0.0373 to 0.1275 |
| **IL6** | 1.37 × 10⁻¹ | 3.70 × 10⁻² | Fragile | P ranged from 0.0370 to 0.1374 |
| **Osteopontin** | 6.60 × 10⁻¹ | 2.67 × 10⁻¹ | Fragile | — |
| **Tryptophan** | 7.33 × 10⁻¹ | 4.11 × 10⁻¹ | Fragile | — |
| **Serotonin** | 8.01 × 10⁻¹ | 2.41 × 10⁻¹ | Fragile | — |
| **Lipoprotein(a)** | 8.25 × 10⁻¹ | 4.87 × 10⁻¹ | Fragile | — |
| **IL1β** | 8.51 × 10⁻¹ | 4.18 × 10⁻¹ | Fragile | — |
| **BMPR1A** | 9.85 × 10⁻¹ | 4.88 × 10⁻¹ | Fragile | — |
| **IDO** | 9.87 × 10⁻¹ | 8.27 × 10⁻¹ | Fragile | — |

Supplementary Table 20. **Leave-one-out sensitivity analysis for reverse Mendelian randomization of calcific aortic stenosis (CAS) on circulating biomarkers.** The reported P-value range was obtained from inverse-variance weighted Mendelian randomization repeated with each SNP removed in turn (leave-one-out analysis).

| **Biomarker** | **SNPs (n)** | **Steiger Pass (n)** | **Avg R² Exposure** | **Avg R² Outcome** | **Pass Rate (%)** | **Direction** |
| --- | --- | --- | --- | --- | --- | --- |
| **ALPL** | 155 | 155 | **0.003762** | 9.03 × 10⁻⁷ | 100 | Forward |
| **IL18 receptor** | 4 | 4 | **0.007505** | 7.54 × 10⁻⁷ | 100 | Forward |
| **IL18** | 6 | 6 | **0.002237** | 6.69 × 10⁻⁷ | 100 | Forward |
| **IL6** | 2 | 2 | **0.001635** | 6.12 × 10⁻⁶ | 100 | Forward |
| **IL6 receptor subunit** | 7 | 7 | **0.042504** | 2.43 × 10⁻⁶ | 100 | Forward |
| **Indole propionate** | 2 | 2 | **0.001467** | 2.12 × 10⁻⁶ | 100 | Forward |
| **LDL cholesterol** | 639 | 639 | **0.003217** | 1.09 × 10⁻⁶ | 100 | Forward |
| **Lipoprotein(a)** | 10 | 10 | **0.022719** | 7.45 × 10⁻⁶ | 100 | Forward |

Supplementary Table 21. **Steiger directionality test for direct Mendelian randomization analyses.**

This table summarizes the results of the Steiger test assessing the directionality of genetic associations in Mendelian randomization. For each marker, the total number of SNPs used, the number passing the Steiger test, the average proportion of variance explained in the exposure and outcome (R²), and the pass rate (%) are reported. P values were derived from the Steiger directionality test comparing variance explained (R²) in the exposure vs. the outcome.

Markers with a high pass rate indicate that the genetic instruments explain more variance in the exposure than in the outcome, supporting the hypothesized causal direction (**Forward**). Markers with low pass rates or labeled as **Reverse** suggest potential reverse causality, indicating caution in interpretation.

| **Biomarker** | **SNPs (n)** | **Steiger Pass (n)** | **Avg R² Exposure** | **Avg R² Outcome** | **Pass Rate (%)** | **Direction** |
| --- | --- | --- | --- | --- | --- | --- |
| **ALPL** | 15 | 8 | 2.10 × 10⁻⁵ | 1.75 × 10⁻⁴ | 53.3 | Forward |
| **IDO** | 33 | 32 | 1.90 × 10⁻⁵ | 5.00 × 10⁻⁶ | 97.0 | Forward |
| **IFN** | 33 | 17 | 1.90 × 10⁻⁵ | 2.10 × 10⁻⁵ | 51.5 | Forward |
| **IFNγ receptor 1** | 33 | 7 | 1.90 × 10⁻⁵ | 7.30 × 10⁻⁵ | 21.2 | Reverse |
| **IL18 receptor** | 33 | 31 | 1.90 × 10⁻⁵ | 1.20 × 10⁻⁵ | 93.9 | Forward |
| **IL18** | 39 | 27 | 1.90 × 10⁻⁵ | 1.80 × 10⁻⁵ | 69.2 | Forward |
| **BMPR1A** | 33 | 23 | 1.90 × 10⁻⁵ | 2.50 × 10⁻⁵ | 69.7 | Forward |
| **IL1β** | 32 | 14 | 1.90 × 10⁻⁵ | 4.00 × 10⁻⁵ | 43.8 | Reverse |
| **IL6** | 35 | 22 | 1.90 × 10⁻⁵ | 2.60 × 10⁻⁵ | 62.9 | Forward |
| **IL6 receptor subunit** | 40 | 23 | 1.90 × 10⁻⁵ | 4.80 × 10⁻⁵ | 57.5 | Forward |
| **Indole propionate** | 8 | 4 | 2.10 × 10⁻⁵ | 2.40 × 10⁻⁵ | 50.0 | Forward |
| **Indole acetate** | 8 | 2 | 2.10 × 10⁻⁵ | 1.23 × 10⁻⁴ | 25.0 | Reverse |
| **Kynurenine** | 8 | 2 | 2.10 × 10⁻⁵ | 4.30 × 10⁻⁵ | 25.0 | Reverse |
| **LDL cholesterol** | 41 | 20 | 2.00 × 10⁻⁵ | 3.30 × 10⁻⁵ | 48.8 | Reverse |
| **Lipoprotein(a)** | 15 | 5 | 2.10 × 10⁻⁵ | 1.97 × 10⁻⁴ | 33.3 | Reverse |
| **Osteopontin** | 34 | 25 | 2.10 × 10⁻⁵ | 2.10 × 10⁻⁵ | 73.5 | Forward |
| **Serotonin** | 8 | 7 | 2.10 × 10⁻⁵ | 1.60 × 10⁻⁵ | 87.5 | Forward |

Supplementary Table 22. Steiger directionality test for indirect Mendelian randomization analyses.
 P values were derived from the Steiger directionality test comparing variance explained (R²) in the exposure vs. the outcome.

| **Biomarker** | **SNPs (n)** | **Status** | **OR** | **95% CI** | **P-value** |
| --- | --- | --- | --- | --- | --- |
| **IL6 receptor subunit** | 7 | Success | 0.971 | 0.960–0.981 | 7.01 × 10⁻⁸ |
| **IL18 receptor** | 4 | Success | 0.653 | 0.511–0.835 | 6.85 × 10⁻⁴ |
| **Lipoprotein(a) †** | 1 | Success | 1.116 | 1.106–1.125 | **< 0.001** |
| **ALPL** | 155 | Success | 0.957 | 0.920–0.996 | 3.25 × 10⁻² |
| **IL18** | 6 | Success | 0.930 | 0.846–1.023 | 1.36 × 10⁻¹ |
| **LDL cholesterol** | 639 | Success | 1.003 | 0.983–1.023 | 7.83 × 10⁻¹ |
| **IDO** | **0** | Weak instrument | — | — | — |
| **IL1β** | **0** | Weak instrument | — | — | — |
| **IL6** | 2 | Weak instrument | — | — | — |
| **Indole propionate** | 2 | Weak instrument | — | — | — |
| **Indole acetate** | **0** | Weak instrument | — | — | — |
| **Kynurenine** | **0** | Weak instrument | — | — | — |
| **Tryptophan** | **0** | Weak instrument | — | — | — |
| **Serotonin** | **0** | Weak instrument | — | — | — |
| **IFNγ receptor 1** | **0** | Weak instrument | — | — | — |
| **IFNγ** | **0** | Weak instrument | — | — | — |
| **BMPR1A** | **0** | Weak instrument | — | — | — |
| **Osteopontin** | **0** | Weak instrument | — | — | — |
