## Supplementary material for "Altered kynurenine and indole tryptophan pathways in calcific aortic stenosis: cross-sectional evidence for a gut-host metabolic imbalance (GUT-CAS)": Main tables

**Table of Contents**

Table 1. Baseline characteristics of patients without calcified aortic stenosis (Non-CAS) and with calcified aortic stenosis (CAS).

Table 2. Description of GWAS consortia used for each phenotype.

**Table 1. Baseline characteristics of patients without calcified aortic stenosis (Non-CAS) and with calcified aortic stenosis (CAS).**

| **Variable** | **Non-CAS (n = 41)** | **CAS (n = 54)** | **p value** |
| --- | --- | --- | --- |
| **Clinical parameters** | | | |
| **Age, years** | 69.00 [60.00–76.00] | 66.00 [60.00–75.50] | 0.482 |
| **Body mass index, kg/m²** | 27.76 [25.93–31.06] | 28.38 [26.58–32.73] | 0.266 |
| **Systolic blood pressure, mmHg** | 123.00 [114.00–141.00] | 135.50 [125.25–141.75] | 0.116 |
| **Diastolic blood pressure, mmHg** | 71.00 [65.00–78.00] | 76.50 [68.00–83.75] | 0.071 |
| **Heart rate, bpm** | 67.00 [60.75–76.50] | 70.00 [63.00–81.00] | 0.145 |
| **Laboratory parameters** | | | |
| **Hemoglobin, g/dL** | 14.45 [13.53–14.97] | 13.90 [12.62–14.97] | 0.097 |
| **Leukocytes, ×10⁹/L** | 6.78 [5.83–7.80] | 6.61 [5.44–8.51] | 0.877 |
| **Platelets, ×10⁹/L** | 208.50 [182.25–260.00] | 217.00 [190.75–244.50] | 0.575 |
| **eGFR, mL/min/1.73m²** | 94.97 [76.16–103.86] | 91.26 [76.46–119.69] | 0.463 |
| **Sodium, mmol/L** | 140.00 [139.00–141.25] | 140.00 [138.25–141.00] | 0.593 |
| **Potassium, mmol/L** | 4.00 [3.90–4.30] | 4.10 [3.90–4.30] | 0.204 |
| **BNP, pg/mL** | 165.00 [81.25–406.75] | 206.00 [63.91–466.25] | 0.738 |
| **Troponin** | 12.00 [7.75–15.50] | 14.50 [9.75–26.25] | 0.080 |
| **CK-MB** | 3.00 [2.20–4.95] | 3.15 [2.20–4.62] | 0.908 |
| **CK** | 117.00 [88.50–159.50] | 135.00 [97.00–195.00] | 0.242 |
| **Albumin, g/L** | 38.00 [36.00–40.50] | 37.00 [36.00–40.00] | 0.600 |
| **CRP, mg/L** | 2.50 [1.00–3.00] | 2.00 [1.00–4.50] | 0.383 |
| **ASAT** | 26.50 [23.00–32.25] | 24.00 [21.00–31.75] | 0.626 |
| **ALAT** | 23.00 [17.00–28.50] | 27.00 [19.00–34.00] | 0.171 |
| **GGT** | 22.00 [20.50–23.50] | 13.00 [2.00–28.00] | 0.723 |
| **Alkaline phosphatase** | 67.50 [55.75–74.00] | 66.50 [52.75–82.25] | 0.850 |
| **Total bilirubin** | 9.75 [8.07–16.05] | 10.00 [8.00–13.50] | 0.821 |
| **Direct bilirubin** | 3.40 [3.00–6.40] | 4.00 [3.25–5.50] | 0.859 |
| **LDH** | 202.00 [187.00–221.50] | 220.00 [168.75–250.75] | 0.638 |
| **Lipid and metabolic profile** | | | |
| **Total cholesterol, mmol/L** | 4.12 [3.57–4.96] | 4.00 [3.05–5.19] | 0.611 |
| **LDL cholesterol, mmol/L** | 1.97 [1.65–3.12] | 2.10 [1.25–3.11] | 0.586 |
| **Triglycerides, mmol/L** | 1.38 [0.98–1.80] | 1.35 [0.88–1.83] | 0.920 |
| **HDL cholesterol, mmol/L** | 1.42 [1.06–1.58] | 1.19 [0.86–1.50] | 0.114 |
| **Lipoprotein(a)** | 21.00 [20.00–100.00] | 20.00 [11.50–85.25] | 0.197 |
| **Glucose, g/L** | 1.00 [0.96–1.12] | 1.02 [0.93–1.08] | 0.944 |
| **HbA1c, %** | 5.70 [5.55–5.80] | 5.60 [5.40–6.20] | 0.776 |
| **Echocardiographic parameters** | | | |
| **LVEF, %** | 60.00 [56.00–65.00] | 60.00 [55.00–62.00] | 0.744 |
| **Aortic max gradient, mmHg** | 7.00 [5.00–10.00] | 48.00 [27.75–68.00] | <0.001 |
| **Aortic mean gradient, mmHg** | 3.50 [3.00–5.00] | 34.50 [20.25–44.00] | <0.001 |
| **Peak velocity, m/s** | 1.23 [1.12–1.60] | 3.50 [2.77–4.02] | <0.001 |
| **Aortic valve area, cm²** | 3.09 [2.88–3.63] | 0.98 [0.80–1.13] | <0.001 |
| **Aortic valve area index** | 1.52 [1.29–1.69] | 0.47 [0.39–0.56] | <0.001 |
| **sPAP, mmHg** | 25.00 [25.00–25.00] | 25.00 [25.00–28.00] | 0.553 |
| **CT and surgical risk** | | | |
| **Calcium score** | 0 | 1451.00 [1047.00–2905.00] | NA |
| **EuroSCORE II** | 0.94 [0.60–1.32] | 0.82 [0.64–1.58] | 0.558 |
| **STS score** | 5.62 [3.70–7.34] | 4.67 [3.60–5.80] | 0.171 |
| **Cardiovascular risk factors and history** | | | |
| **Female sex** | 7 (17.1%) | 13 (24.1%) | 0.456 |
| **Hypertension** | 27 (65.9%) | 32 (59.3%) | 0.531 |
| **Dyslipidemia** | 21 (51.2%) | 23 (43.4%) | 0.533 |
| **Diabetes mellitus** | 2 (4.9%) | 6 (11.1%) | 0.459 |
| **Current smoker** | 7 (17.1%) | 4 (7.4%) | 0.198 |
| **Previous smoker** | 8 (19.5%) | 15 (27.8%) | 0.469 |
| **Coronary artery disease** | 7 (17.5%) | 14 (25.9%) | 0.454 |
| **Previous PCI** | 7 (17.5%) | 5 (9.3%) | 0.349 |

*Continuous variables are presented as median [interquartile range], and categorical variables as n (%). P values for continuous variables were derived from Student’s t test (normally distributed variables) or the Mann–Whitney U test (non-normally distributed variables), with normality assessed by the Shapiro–Wilk test; P values for categorical variables were derived from the chi-squared test or Fisher’s exact test.*

**Table 2. Description of GWAS consortia used for each phenotype.**

| **Variable** | **GWAS ID (raw)** | **First author (year)** | **Consortium / Source** | **Sample size** | **Population** | **Sex** |
| --- | --- | --- | --- | --- | --- | --- |
| **Aortic stenosis** | GCST90651074 | Kany (2025) | GBMI | 1,984,229 | European | Mixed |
| **Indole-3-propionate** | met-a-475 | Shin (2014) | Non available | 7,803 | European | Mixed |
| **Indole-3-acetate** | met-a-446 | Shin (2014) | Non available | 7618 | European | Mixed |
| **Kynurenine** | met-a-375 | Shin (2014) | Non available | 7816 | European | Mixed |
| **Tryptophan** | met-a-584 | Shin (2014) | Non available | 7804 | European | Mixed |
| **Serotonin** | met-a-358 | Shin (2014) | Non available | 6139 | European | Mixed |
| **Interleukin-1β** | ebi-a-GCST004448 | Ahola-Olli (2016) | Non available | 3,309 | European | Mixed |
| **Interleukin-6** | ebi-a-GCST90012005 | Folkersen (2020) | Non available | 21,758 | European | Mixed |
| **Interleukin-6 receptor subunit a** | ebi-a-GCST90012025 | Folkersen (2020) | Non available | 21,758 | European | Mixed |
| **Interleukin-18** | ebi-a-GCST90012024 | Folkersen (2020) | Non available | 21,758 | European | Mixed |
| **Interleukin-18 receptor** | prot-a-1491 | Sun BB (2020) | Non available | 3301 | European | Mixed |
| **Interferon-γ** | prot-a-1428 | Sun BB (2018) | Non available | 3,301 | European | Mixed |
| **Interferon-γ receptor 1** | prot-a-1430 | Sun BB (2018) | Non available | 3,301 | European | Mixed |
| **Indoleamine 2,3-dioxygenase (IDO1)** | prot-a-1410 | Sun BB (2018) | Non available | 3,301 | European | Mixed |
| **Bone morphogenetic protein receptor type 1A** | prot-a-259 | Sun BB (2018) | Non available | 3,301 | European | Mixed |
| **Low-density lipoprotein cholesterol** | ebi-a-GCST90002412 | Klimentidis YC (2020) | Non available | 431,167 | European | Non available |
| **Lipoprotein(a)** | ebi-a-GCST90025993 | Barton AR (2021) | Non available | 348,806 | European | Non available |
| **Alkaline phosphatase** | ebi-a-GCST90025947 | Barton AR (2021) | Non available | 437,896 | European | Non available |
| **Osteopontin** | ebi-a-GCST90010244 | Gilly (2016) | Non available | 1,322 | European | Non available |

*GWAS sources are shown as provided in the source table. Population and sex fields are reported according to available source metadata. This table is descriptive; it summarizes the source GWAS datasets and contains no P values; no statistical tests were performed.*
