## Supplementary methods for "Altered kynurenine and indole tryptophan pathways in calcific aortic stenosis: cross-sectional evidence for a gut-host metabolic imbalance (GUT-CAS)"

**Metabolomics detailed methods**

**Bile acids^1^**

Serum samples were thawed at room temperature and vortexed before processing. For absolute quantification of bile acids, 50 µL of serum, calibration standards, and quality controls prepared in methanol/water (1:1) were mixed with 400 µL of internal standard solution in isopropanol in a 96-deepwell plate. After incubation at −20 °C for 30 min, samples were centrifuged at 4 °C (3000 rpm, 30 min). A volume of 350 µL of supernatant was transferred and evaporated under nitrogen at 40 °C, then reconstituted in 100 µL methanol/water (1:1), centrifuged again, and 80 µL transferred to a 96-well plate for LC-MS/MS injection (5 µL). For highly concentrated samples, an additional 1:10 dilution step was performed prior to extraction. Chromatographic separation was achieved on a UPLC I-Class system using an Acquity BEH C8 column, and detection was performed on a Xevo TQ-XS mass spectrometer in MRM mode with ESI ±. Quantification was based on metabolite-to–internal standard peak area ratios using external calibration curves, validated by quality controls and expressed within the established LLOQ–ULOQ ranges.

**Short-chain fatty acids^2^**

SCFAs were quantified in serum using a derivatization-based LC-MS/MS method adapted from Monteiro et al. Briefly, 50 µL of serum was mixed with 150 µL methanol, shaken, incubated at −20 °C, and centrifuged. An aliquot of supernatant (or a 1:10 diluted extract for highly concentrated samples) was subjected to derivatization with internal standards, pyridine, EDC, and 3-nitrophenylhydrazine, followed by incubation for 1 h at 4 °C. The reaction was quenched with formic acid–containing water and the plate was shaken prior to LC-MS/MS analysis (15 µL injection). Separation was performed on a C18 column using a water/acetonitrile gradient with formic acid, and detection was carried out on a Xevo TQ-XS in negative ESI MRM mode. Concentrations were calculated from calibration curves built on metabolite-to–internal standard response ratios and expressed in nM, with batch validity ensured by quality control samples within predefined acceptance limits.

**TMA, TMAO, choline, and L-carnitine**^3^

Serum levels of TMA, TMAO, choline, and L-carnitine were measured after ethyl bromoacetate derivatization. Fifty microliters of serum, calibration standards, and quality controls were spiked with internal standards, derivatized with ethyl bromoacetate, and incubated at room temperature under agitation. The reaction was quenched with acetonitrile/water/formic acid, followed by centrifugation, and 2 µL of supernatant were injected into the LC-MS/MS system. Chromatographic separation was performed on a Cortecs HILIC column using ammonium formate/formic acid buffers, and detection was achieved on a Xevo TQ-XS in positive ESI MRM mode. Absolute concentrations were obtained from external calibration curves using metabolite-to–internal standard response ratios, with quality controls used to validate analytical performance.

**Tryptophan metabolites**^4^

Samples were allowed to return to room temperature and vortexed prior to processing. Calibration standards and quality control samples were prepared in PBS containing 4% (w/v) BSA. For extraction, 200 µL of internal standard solution in methanol were added to 50 µL of sample, calibration standard, or quality control in a 96-deepwell plate. Plates were sealed and centrifuged (4 °C, 3000 rpm, 30 min). A total of 175 µL of supernatant was transferred to a new plate and evaporated under a nitrogen stream at 40 °C for 30 min. The dried residue was reconstituted in 100 µL of methanol/water (1:9, v/v), followed by centrifugation (4 °C, 3000 rpm, 10 min). Finally, 80 µL of each sample was transferred to a 96-well plate, and 5 µL was injected into the LC-MS/MS system.

Tryptophan and its downstream metabolites, including kynurenine pathway and indole derivatives, were quantified using a targeted LC-MS/MS method following methanol extraction and addition of isotope-labeled internal standards. Serum samples, calibration standards, and quality controls were spiked with internal standards, derivatized with ethyl bromoacetate, quenched, centrifuged, and analyzed by UPLC-MS/MS on a HILIC column with positive ESI in MRM mode. Quantification relied on metabolite-to–internal standard peak area ratios and external calibration curves covering the validated LLOQ–ULOQ ranges. Batch accuracy and precision were monitored using quality control samples, and concentrations were reported as absolute values after correction for recovery and instrumental variability.

**Metabolomics detailed methods**

**DNA extraction and sample preparation**

Total DNA was extracted using the ZymoBIOMICS DNA MagBead Kit (Zymo Research, D4311) on a KingFisher Apex automated platform (Thermo Fisher Scientific), following the manufacturer’s instructions.

For tissue samples, approximately 15 mg of material was transferred into ZR BashingBead Lysis Tubes (2.0 mm) containing 750 µL DNA/RNA Shield. For fecal samples, 100 mg of material was processed using ZR BashingBead Lysis Tubes (0.1 and 0.5 mm) with 750 µL DNA/RNA Shield.

Samples were homogenized using a FastPrep-96 instrument (MP Biomedicals) with five bead-beating cycles (1 min at 6.5 m/s, followed by 5 min rest).

**DNA extraction and quality control**

After homogenization, 200 µL of lysate was transferred to a 96-deepwell plate for automated extraction. DNA quantity and quality were assessed using a Qubit 4.0 fluorometer (dsDNA HS assay), FEMTO Pulse system (Agilent), and Denovix DS-11 spectrophotometer.

**Library preparation and sequencing**

Full-length 16S rRNA gene libraries (V1–V9 regions) were prepared using the PacBio Kinnex workflow (PacBio, Rev05, Jan 2025). Amplification was performed using universal primers targeting the 27F and 1492R regions with sample-specific barcodes.

Library quality and fragment size (~18 kb) were assessed using Qubit and Fragment Analyzer systems. SMRTbell libraries were prepared according to PacBio protocols and sequenced on the PacBio Revio platform using 30-hour movie acquisition and adaptive loading at 160 pM.

**Controls**

Positive controls included commercially available microbial community standards (ZymoBIOMICS DNA standards and ATCC MSA-3001). Negative controls included extraction blanks and PCR no-template controls to monitor contamination.

Bibliographie

*(1)	Sarafian, M. H.; Lewis, M. R.; Pechlivanis, A.; Ralphs, S.; McPhail, M. J. W.; Patel, V. C.; Dumas, M.-E.; Holmes, E.; Nicholson, J. K. Bile Acid Profiling and Quantification in Biofluids Using Ultra-Performance Liquid Chromatography Tandem Mass Spectrometry. Anal. Chem.* ***2015****, 87 (19), 9662–9670. https://doi.org/10.1021/acs.analchem.5b01556.*

*(2)	Monteiro, J.; Lefèvre, A.; Dufour-Rainfray, D.; Oury, A.; Chicheri, G.; Galineau, L.; Blasco, H.; Nadal-Desbarats, L.; Emond, P. Multi-Compartment SCFA Quantification in Human. Am. J. Anal. Chem.* ***2024****, 15 (06), 177–200. https://doi.org/10.4236/ajac.2024.156012.*

*(3)	Andrikopoulos, P.; Aron-Wisnewsky, J.; Chakaroun, R.; Myridakis, A.; Forslund, S. K.; Nielsen, T.; Adriouch, S.; Holmes, B.; Chilloux, J.; Vieira-Silva, S.; Falony, G.; Salem, J.-E.; Andreelli, F.; Belda, E.; Kieswich, J.; Chechi, K.; Puig-Castellvi, F.; Chevalier, M.; Le Chatelier, E.; Olanipekun, M. T.; Hoyles, L.; Alves, R.; Helft, G.; Isnard, R.; Køber, L.; Coelho, L. P.; Rouault, C.; Gauguier, D.; Gøtze, J. P.; Prifti, E.; Froguel, P.; MetaCardis Consortium; Zucker, J.-D.; Bäckhed, F.; Vestergaard, H.; Hansen, T.; Oppert, J.-M.; Blüher, M.; Nielsen, J.; Raes, J.; Bork, P.; Yaqoob, M. M.; Stumvoll, M.; Pedersen, O.; Ehrlich, S. D.; Clément, K.; Dumas, M.-E. Evidence of a Causal and Modifiable Relationship between Kidney Function and Circulating Trimethylamine N-Oxide. Nat. Commun.* ***2023****, 14 (1), 5843. https://doi.org/10.1038/s41467-023-39824-4.*

*(4)	Alarcan, H.; Chaumond, R.; Emond, P.; Benz-De Bretagne, I.; Lefèvre, A.; Bakkouche, S.-E.; Veyrat-Durebex, C.; Vourc’h, P.; Andres, C.; Corcia, P.; Blasco, H. Some CSF Kynurenine Pathway Intermediates Associated with Disease Evolution in Amyotrophic Lateral Sclerosis. Biomolecules* ***2021****, 11 (5), 691. https://doi.org/10.3390/biom11050691.*
