## Supplementary material for "Altered kynurenine and indole tryptophan pathways in calcific aortic stenosis: cross-sectional evidence for a gut-host metabolic imbalance (GUT-CAS)": STROBE

STROBE Statement—checklist of items that should be included in reports of observational studies

|  | Item No. | Recommendation | Page  No. | Relevant text from manuscript |
| --- | --- | --- | --- | --- |
| **Title and abstract** | 1 | (*a*) Indicate the study’s design with a commonly used term in the title or the abstract | 1, 3 | Title: "Tryptophan-Kynurenine shunt and depletion of indole-producing Firmicutes: A new Gut-Heart axis in Calcific Aortic Stenosis (GUT-CAS)". Abstract: "Methods: In a prospective cohort of 54 patients with CAS and 41 age, sex, BMI-balanced non-CAS controls..." |
|  |  | (*b*) Provide in the abstract an informative and balanced summary of what was done and what was found | 3 | Introduction: "CAS is a progressive valvular disease characterized by lipid accumulation, inflammation, and osteogenic remodeling." Methods: "...we performed integrated microbiome and metabolomic profiling." Results: "CAS patients exhibited a distinct tryptophan metabolic profile..." Conclusion: "CAS is characterized by a focused gut-host metabolic reprogramming..." |
| Introduction | | | |  |
| Background/rationale | 2 | Explain the scientific background and rationale for the investigation being reported | 6 | "Calcific aortic stenosis (CAS) is the leading cause of death from valvular heart disease in high-income countries, with prevalence increasing sharply with age." "In parallel, growing evidence implicates the gut microbiota as a central regulator of host metabolic and inflammatory homeostasis." "Despite these advances, the specific contribution of the gut microbiota-metabolite axis to CAS remains poorly defined." |
| Objectives | 3 | State specific objectives, including any prespecified hypotheses | 6 | "In this study, we performed an integrated analysis of gut microbiota composition and circulating microbial metabolites in patients with and without CAS. By combining microbiome profiling with targeted metabolomics and Mendelian randomization, we aimed to characterize the gut-valve metabolic axis and assess its potential causal contribution to CAS." |
| Methods | | | |  |
| Study design | 4 | Present key elements of study design early in the paper | 7 | "The GUT-CAS study is a prospective observational cohort study conducted at Bern University Hospital in Switzerland." ClinicalTrials.gov registration: NCT06021535. |
| Setting | 5 | Describe the setting, locations, and relevant dates, including periods of recruitment, exposure, follow-up, and data collection | 7 | "Bern University Hospital, Switzerland. Adult patients (≥18 years) with CAS and control participants without CAS were recruited." "Stool samples were collected at baseline in CAS patients and controls. Stool sampling was repeated in CAS patients annually." "Patients with mild, moderate, or asymptomatic severe CAS who did not undergo aortic valve replacement were followed annually for 5 years." ClinicalTrials.gov: NCT06021535. |
| Participants | 6 | (*a*) *Cohort study*—Give the eligibility criteria, and the sources and methods of selection of participants. Describe methods of follow-up  *Case-control study*—Give the eligibility criteria, and the sources and methods of case ascertainment and control selection. Give the rationale for the choice of cases and controls  *Cross-sectional study*—Give the eligibility criteria, and the sources and methods of selection of participants | 7 | "Adult patients (≥18 years) with CAS and control participants without CAS were recruited. CAS severity was defined according to current echocardiographic guidelines. Controls had no evidence of aortic valve disease on echocardiography." "All participants underwent standardized clinical evaluation, laboratory testing (including cardiac biomarkers, lipid profile, glycemic markers, renal and hepatic function), electrocardiography, and echocardiography." Follow-up: annually for 5 years in non-surgical patients. |
|  |  | (*b*) *Cohort study*—For matched studies, give matching criteria and number of exposed and unexposed  *Case-control study*—For matched studies, give matching criteria and the number of controls per case | 10 | "Controls were selected to achieve comparable distributions of age, sex, and BMI relative to the CAS group." "A caliper-based selection approach was applied in which control subjects with a BMI within ±2.5 kg/m² of a given CAS patient were considered eligible. Selection was performed without replacement... When multiple eligible controls were available for a given CAS patient, one control was randomly selected." Final cohort: 54 CAS patients, 41 matched controls. |
| Variables | 7 | Clearly define all outcomes, exposures, predictors, potential confounders, and effect modifiers. Give diagnostic criteria, if applicable | 7–9 | Outcome: CAS status (defined by echocardiography per current guidelines). Exposures: gut microbiota composition (16S rRNA full-length sequencing, V1-V9), circulating metabolites (tryptophan derivatives, bile acids, SCFAs, TMAO/choline pathway). Confounders: age, sex, BMI, statin use, diabetes. "Clearly define all outcomes, exposures, predictors, potential confounders, and effect modifiers." Clinical variables: cardiac biomarkers, lipid profile, glycemic markers, renal and hepatic function. |
| Data sources/ measurement | 8* | For each variable of interest, give sources of data and details of methods of assessment (measurement). Describe comparability of assessment methods if there is more than one group | 8–9 | Microbiota: "Full-length 16S rRNA gene libraries (V1-V9 regions) were generated using the PacBio Kinnex platform and sequenced on the PacBio Revio system. Taxonomic classification was performed against Silva reference databases using QIIME2-compatible classifiers." Metabolomics: "Serum and fecal metabolites were quantified using high-performance liquid chromatography coupled to mass spectrometry (HPLC-MS)." Classes analyzed: bile acids, tryptophan metabolites (kynurenine derivatives, indole derivatives), SCFAs, choline/TMAO pathway. "Quality control samples ensured analytical accuracy and reproducibility." |
| Bias | 9 | Describe any efforts to address potential sources of bias | 10 | "Controls were selected to achieve comparable distributions of age, sex, and BMI... BMI balancing was performed algorithmically because adiposity is a major determinant of gut microbiota composition." "Sensitivity analyses for BMI, age, sex, and diabetes did not materially modify the results." "Because statins influence both cholesterol metabolism and gut microbial composition, we performed stratified analyses to assess whether lipid-lowering therapy modified these associations." FDR correction applied using the Benjamini-Hochberg procedure. |
| Study size | 10 | Explain how the study size was arrived at | 9 | "We estimated that enrolling 50 participants per group (CAS and controls) would provide sufficient power to detect differences in microbiota and metabolite. Based on previously published studies with comparable designs and methodologies, a sample size of [50/group] was considered adequate for this exploratory study." Enrolled: 54 CAS patients and 41 controls. |

Continued on next page

| Quantitative variables | 11 | Explain how quantitative variables were handled in the analyses. If applicable, describe which groupings were chosen and why | 10 | "Continuous variables are expressed as mean ± standard deviation (SD) or median and interquartile range (IQR); categorical variables as frequencies and proportions." "For circulating and fecal metabolites, including bile acids, SCFAs, tryptophan derivatives, and choline/TMAO pathway compounds, concentrations were log-transformed when necessary to approximate normality." BMI categories: <25 vs. ≥25 kg/m². CAS severity: severe vs. non-severe per echocardiographic guidelines. |
| --- | --- | --- | --- | --- |
| Statistical methods | 12 | (*a*) Describe all statistical methods, including those used to control for confounding | 10 | "All analyses were performed using R or python (R v4.5.1 for maaslin3 analysis and R v4.2.1/python 3.10.4 for other analysis) and GraphPad Prism (v10)." "Group comparisons were performed using t tests or Mann-Whitney U tests. Correlations assessed by Spearman or Pearson." "FDR correction for multiple comparisons was applied using the Benjamini-Hochberg procedure." "Differential abundance analysis was performed using MaAsLin3, applying multivariable linear models to identify taxa associated with clinical variables, including CAS status, BMI, and sex." MR primary estimate: Inverse Variance Weighted (IVW) method. |
|  |  | (*b*) Describe any methods used to examine subgroups and interactions | 11 | "Predefined exploratory subgroup analyses were conducted according to: (i) disease severity (severe vs. non-severe CAS), (ii) sex (female vs. male)." "These analyses were performed to investigate potential effect modifications of gut-derived and tryptophan-related metabolites by biological sex, metabolic status, and hemodynamic severity of aortic stenosis." "Additional stratified analyses were performed by BMI category (normal weight vs. overweight/obese)." "Stratified analyses by statin use were performed to assess lipid-lowering therapy effects." |
|  |  | (*c*) Explain how missing data were addressed | 10 | No formal imputation was performed. Complete case analyses were used; participants with available data at each time point were included in respective analyses. Missing data were not formally described; participants with missing variables were excluded from individual analyses. |
|  |  | (*d*) *Cohort study*—If applicable, explain how loss to follow-up was addressed  *Case-control study*—If applicable, explain how matching of cases and controls was addressed  *Cross-sectional study*—If applicable, describe analytical methods taking account of sampling strategy | 9 | "Patients with mild, moderate, or asymptomatic severe CAS who did not undergo aortic valve replacement were followed annually for 5 years, with clinical assessment, echocardiography, and repeated stool/blood sampling." "Patients who underwent valve replacement were followed longitudinally post-intervention." Loss to follow-up not separately detailed; analyses reported at cross-sectional baseline unless stated. |
|  |  | (*e*) Describe any sensitivity analyses | 12, 16 | "Sensitivity analyses for BMI, age, sex, and diabetes did not materially modify the results." "Because statins influence both cholesterol metabolism and gut microbial composition, we performed stratified analyses." MR sensitivity: "IVW, weighted median, MR-Egger, Leave-one-out analyses, for both direct and reverse MR (Supplementary Tables 12-19)." "Steiger filtering (Supplementary Tables 20-21) further supported correct directionality in forward MR." "Associations were not driven by outlier instruments." |
| Results | | | | |
| Participants | 13* | (a) Report numbers of individuals at each stage of study—eg numbers potentially eligible, examined for eligibility, confirmed eligible, included in the study, completing follow-up, and analysed | 13 | "We prospectively enrolled 54 patients with CAS (median age 66.0 years, IQR 60.0-75.5) and 41 control subjects without CAS (median age 69.0 years, IQR 60.25-76.0) (Figure 1)." Flow diagram in Figure 1 presents numbers at each stage of recruitment, eligibility confirmation, and analysis inclusion. |
|  |  | (b) Give reasons for non-participation at each stage | 13 | Reasons for non-participation and exclusion at each stage are presented in the participant flow diagram (Figure 1). Controls were algorithmically selected from those with BMI within ±2.5 kg/m² of eligible CAS patients; unmatched controls were excluded. |
|  |  | (c) Consider use of a flow diagram | 13 | Flow diagram presented in Figure 1, depicting the number of participants screened, enrolled, and included in analyses (CAS: n=54; controls: n=41). |
| Descriptive data | 14* | (a) Give characteristics of study participants (eg demographic, clinical, social) and information on exposures and potential confounders | 13 | "Baseline demographic, cardiovascular, inflammatory, and metabolic characteristics were summarized in Table 1." "We prospectively enrolled 54 patients with CAS (median age 66.0 years, IQR 60.0-75.5) and 41 control subjects without CAS (median age 69.0 years)." "Baseline characteristics were comparable between groups." Characteristics include age, sex, BMI, cardiovascular risk factors, medication use, cardiac biomarkers, lipid profile, glycemic and renal markers. |
|  |  | (b) Indicate number of participants with missing data for each variable of interest | 10 | Missing data per variable not formally reported per item. Complete case analyses used throughout. Participants with unavailable samples or data at a given time point were excluded from those specific analyses. No imputation performed. |
|  |  | (c) *Cohort study*—Summarise follow-up time (eg, average and total amount) | 9 | "Patients with mild, moderate, or asymptomatic severe CAS who did not undergo aortic valve replacement were followed annually for 5 years." "Patients who underwent valve replacement were followed longitudinally post-intervention." Present analyses focus on baseline cross-sectional metabolomic and microbiome comparisons. |
| Outcome data | 15* | *Cohort study*—Report numbers of outcome events or summary measures over time | 13 | "We prospectively enrolled 54 patients with CAS... and 41 control subjects without CAS." "Baseline characteristics were comparable between groups." Primary outcomes: metabolomic and microbiome profiles at baseline. Key findings: elevated kynurenine-pathway metabolites; reduced indole-3-sulfate; depletion of Eubacterium coprostanoligenes in CAS vs. controls. |
|  |  | *Case-control study—*Report numbers in each exposure category, or summary measures of exposure | N/A | Not applicable – cohort study design. |
|  |  | *Cross-sectional study—*Report numbers of outcome events or summary measures | N/A | Not applicable – cohort study design. |
| Main results | 16 | (*a*) Give unadjusted estimates and, if applicable, confounder-adjusted estimates and their precision (eg, 95% confidence interval). Make clear which confounders were adjusted for and why they were included | 13–16 | "CAS patients exhibited a distinct tryptophan metabolic profile, characterized by a tendency toward increased kynurenine-pathway metabolites and lower indole-3-sulfate." "Effect-size ranking using Cliff's δ also supported modest but directionally consistent differences." Largest positive effects: indole-3-acetamide (δ=0.282), 3-hydroxyanthranilic acid (δ=0.242), xanthurenic acid (δ=0.229). "Alpha diversity indices did not differ between CAS and non-CAS patients (all p>0.05)." "Eubacterium coprostanoligenes depleted in statin-naive CAS patients." MR: "significant associations observed for lipoprotein(a), IL6 receptor α subunit, IL18 receptor, and ALPL" (forward direction). "Kynurenine and indole derivatives significantly altered in reverse MR." Confounders adjusted for: BMI, sex, age, statin use. Results presented with p-values and effect sizes (Cliff's δ, log2FC). |
|  |  | (*b*) Report category boundaries when continuous variables were categorized | 10, 15 | CAS severity: severe defined as aortic valve area ≤1.0 cm² per echocardiographic guidelines (severe N=32, non-severe N=16). BMI categories: <25 kg/m² (normal weight) vs. ≥25 kg/m² (overweight/obese). "CAS patients with severe CAS (N=32) compared to those with mild-moderate disease (N=16)." |
|  |  | (*c*) If relevant, consider translating estimates of relative risk into absolute risk for a meaningful time period | N/A | Not applicable – study reports continuous metabolite concentrations and microbiota relative abundance; no relative risk estimates translated to absolute risk for a time period. |

Continued on next page

| Other analyses | 17 | Report other analyses done—eg analyses of subgroups and interactions, and sensitivity analyses | 15–16 | Subgroup analyses by CAS severity (severe vs. mild-moderate, Figure 3; N=32 vs. N=16) and by sex (male N=41 vs. female N=13). "Sensitivity analyses for BMI, age, sex, and diabetes did not materially modify the results." "Stratified analyses by statin use." MR sensitivity analyses: "IVW, weighted median, MR-Egger, Leave-one-out analyses (Supplementary Tables 12-19)." "Steiger filtering (Supplementary Tables 20-21) further supported correct directionality." |
| --- | --- | --- | --- | --- |
| Discussion | | | | |
| Key results | 18 | Summarise key results with reference to study objectives | 17 | "In this integrated metabolomic and microbiome analysis of well-matched patients with CAS, we observed a selective gut-host metabolic signature characterized by preferential activation of the kynurenine branch of tryptophan metabolism alongside subtle depletion of specific Firmicutes taxa." "A second key observation is the relative depletion in CAS patients of cholesterol-reducing gut bacteria, most notably Eubacterium coprostanoligenes." "Bidirectional Mendelian randomization establishes a causal hierarchy in CAS — positioning LDL and Lp(a) as upstream drivers and kynurenine pathway metabolites as downstream disease sequelae." |
| Limitations | 19 | Discuss limitations of the study, taking into account sources of potential bias or imprecision. Discuss both direction and magnitude of any potential bias | 21 | "This study has several limitations that should be acknowledged. First, the sample size is relatively small, particularly for subgroup analyses, which may have limited statistical power to detect subtle differences, such as early-stage kynurenine pathway alterations or sex-specific effects." "[Cross-sectional metabolomic data precludes causal inference at the individual patient level]." "The study was conducted at a single centre (Bern University Hospital), which may limit generalisability." "Residual BMI imbalance within the BMI-balanced cohort (BMI SMD=0.27) was explored in sensitivity analyses." |
| Interpretation | 20 | Give a cautious overall interpretation of results considering objectives, limitations, multiplicity of analyses, results from similar studies, and other relevant evidence | 22 | "CAS emerges from this analysis as a microbiota-influenced immunometabolic disease, shaped by targeted alterations in gut-derived metabolic pathways rather than by global dysbiosis." "These findings support a model in which microbiota-derived inflammatory signaling and defective microbial cholesterol handling actively drive the initiation and progression of CAS." "By identifying the kynurenine pathway and microbial cholesterol transformation as central mediators, these data open new therapeutic avenues for the prevention and treatment of this prevalent and currently untreatable disease." |
| Generalisability | 21 | Discuss the generalisability (external validity) of the study results | 21 | "The study was conducted at a single centre (Bern University Hospital, Switzerland), predominantly in elderly patients. Findings require validation in larger, multicentre, and ethnically diverse cohorts." "Given the limited sample size, subgroup analyses were considered hypothesis-generating and should be interpreted with caution." "Mendelian randomization analyses used publicly available GWAS summary statistics from large European-ancestry cohorts, which may limit generalisability to other populations." |
| Other information | |  | | |
| Funding | 22 | Give the source of funding and the role of the funders for the present study and, if applicable, for the original study on which the present article is based | 23 | "The authors gratefully acknowledge financial support from the Peter Bockhoff Foundation, the Sana Foundation, and the Swiss Life Foundation for the metabolomics and metagenomics analyses." Conflicts of interest are declared individually per author in the Declarations section. Thomas Pilgrim: grants from Swiss National Science Foundation, Swiss Heart Foundation. Caroline Chong-Nguyen: research grants from French Society of Cardiology, MIDHAS group, Swiss Life Foundation, Sana Foundation, Peter Bockhoff Foundation. No role of funders in study design, data collection, or interpretation is stated. |

*Give information separately for cases and controls in case-control studies and, if applicable, for exposed and unexposed groups in cohort and cross-sectional studies.

**Note:** An Explanation and Elaboration article discusses each checklist item and gives methodological background and published examples of transparent reporting. The STROBE checklist is best used in conjunction with this article (freely available on the Web sites of PLoS Medicine at http://www.plosmedicine.org/, Annals of Internal Medicine at http://www.annals.org/, and Epidemiology at http://www.epidem.com/). Information on the STROBE Initiative is available at www.strobe-statement.org.
